# Metagenomics reveals novel microbial signatures of farm exposures in house dust

**DOI:** 10.1101/2023.04.07.23288301

**Authors:** Ziyue Wang, Kathryn R. Dalton, Mikyeong Lee, Christine G. Parks, Laura E. Beane Freeman, Qiyun Zhu, Antonio González, Rob Knight, Shanshan Zhao, Alison A Motsinger-Reif, Stephanie J. London

**Author notes:** **Correspondence:** Stephanie J. London. Equal contribution co-first authors (Ziyue Wang, Kathryn R. Dalton). Co-senior authors (Alison A Motsinger-Reif, Stephanie J. London).

## Abstract

Indoor home dust microbial communities, important contributors to human health outcomes, are shaped by environmental factors, including farm-related exposures. Detection and characterization of microbiota are influenced by sequencing methodology; however, it is unknown if advanced metagenomic whole genome shotgun sequencing (WGS) can detect novel associations between environmental exposures and the indoor built-environment dust microbiome, compared to conventional 16S rRNA amplicon sequencing (16S). This study aimed to better depict indoor dust microbial communities using WGS to investigate novel associations with environmental risk factors from the homes of 781 farmers and farm spouses enrolled in the Agricultural Lung Health Study. We examined various farm-related exposures, including living on a farm, crop versus animal production, and type of animal production, as well as non-farm exposures, including home cleanliness and indoor pets. We assessed the association of the exposures on within-sample alpha diversity and between-sample beta diversity, and the differential abundance of specific microbes by exposure. Results were compared to previous findings using 16S. We found most farm exposures were significantly positively associated with both alpha and beta diversity. Many microbes exhibited differential abundance related to farm exposures, mainly in the phyla *Actinobacteria, Bacteroidetes, Firmicutes*, and *Proteobacteria*. The identification of novel differential taxa associated with farming at the genera level, including *Rhodococcus, Bifidobacterium, Corynebacterium*, and *Pseudomonas*, was a benefit of WGS compared to 16S. Our findings indicate that characterization of dust microbiota, an important component of the indoor environment relevant to human health, is heavily influenced by sequencing techniques. WGS is a powerful tool to survey the microbial community that provides novel insights on the impact of environmental exposures on indoor dust microbiota, and should be an important consideration in designing future studies in environmental health.

## 1 Introduction

Humans spend 90% of their lives indoors (1), with much of this time spent in the home, where they both contribute to and are exposed to environmental microbiota. Home dust microbiota are commonly captured by vacuuming living spaces, including bedrooms. Exposure to bacterial and fungal communities inside the home has been associated with allergic, atopic, and respiratory conditions in children and adults (2-5). These associations could reflect the direct impacts of environmental microbial exposure on inhabitants’ health, as well as through indirect effects of dust microbiota on the human gut, skin, oral, and respiratory microbiomes (6-8). Housing characteristics and other environmental exposures have been shown to influence indoor microbial communities, including farm-related exposures (8-11). Living in or near a farm environment entails unique microbial exposures and subsequent health concerns. Farm exposures have been associated with altered microbial composition in home dust, which in turn have been associated with allergic outcomes in adults and children (4, 12-14). Identifying environmental factors that influence home dust microbiota is a critical first step in determining exposure pathways relevant to health outcomes.

The emergence and optimization of high-throughput sequencing have enabled new approaches to assessing the composition of bacterial communities present in home dust samples, which have a complex matrix and low microbial biomass compared to host-associated microbiome samples such as stool. 16S rRNA amplicon sequencing (16S) is a traditional next-generation technique in which all amplified products are sequenced from a single gene (i.e., the 16S rRNA gene). The technique is limited, however, because annotation is based on putative associations of the 16S rRNA gene with bacterial taxa defined computationally as operational taxonomic units (OTUs). Thus, specific bacterial entities are not directly sequenced, but rather predicted based on OTUs, and consequently have more uncertainty at the lower taxonomy ranks of genus and species (15-18). Metagenomic whole genome shotgun sequencing (WGS), in which random fragments of the genome are sequenced, is an alternative approach and offers a major advantage in that taxa can be more accurately defined at the genus/species level (16, 19). However, WGS is more expensive and requires more extensive data processing and analysis (15, 20). Most of the published data on associations of home dust microbiota with environmental exposures or health outcomes have relied on the older 16S methodology.

Higher taxonomic classification resolution with WGS provides a more comprehensive description of the microbial community, and may improve the ability to detect novel associations with environmental risk factors, which is important when considering environmental health pathways. In human microbial communities, especially the gut microbiome, WGS generally identifies a larger number of unique phyla and higher overall microbial diversity within samples compared to 16S (16, 19-26). However, results are mixed for environmental samples in water and soil (27, 28). At present, no research has evaluated sequencing methodology on microbial community characterization in indoor home dust samples, and how this will impact the upstream associations with farm and non-farm environmental exposures.

In the present study, we analyzed samples from 781 participant homes in the Agricultural Lung Health Study (ALHS), a study of farmers and their spouses in North Carolina and Iowa, using advanced WGS methods, and evaluated associations with farm and nonfarm exposures found to be important in previous work based on 16S, in this cohort and others (4, 8, 29). We considered both microbial community diversity levels and specific bacterial taxa, in order to determine whether WGS can provide novel insights into farming environmental exposure pathways, the results of which are relevant to the design of future research integrating environmental health and microbiology.

## 2 Materials and methods

### 2.1 Study population and design

ALHS is a case-control study of adult asthma study nested within the Agricultural Health Study (AHS), a prospective cohort of licensed pesticide applicators, mostly farmers and their spouses, enrolled between 1993 and 1997 (30). ALHS participants were selected from among AHS participants who were either farmers or farm spouses in North Carolina (NC) and Iowa (IA) and completed an AHS telephone follow-up conducted from 2005-2010. ALHS enrolled individuals with asthma diagnosis and current asthma symptoms or medication use along with individuals with symptoms and medication use suggesting likely asthma (n = 1,223). The comparison group was a random sample of AHS participants without these criteria (n = 2,078). The Supplemental Methods further details study population selection and inclusion criteria. The Institutional Review Board at the National Institute of Environmental Health Sciences approved the study. Written informed consent was obtained from all participants.

### 2.2 Dust sample and environmental exposure data collection

Of the 3,301 ALHS participants, 2,871 received a home visit and had adequate levels of collected dust from the bedroom (Figure 1), as described in Carnes et al. (31). A trained field technician vacuumed two 1-yd^2^ (0.84-m^2^) areas—one on participants’ sleeping surface and one on the floor next to the bed— for 2 min each with a DUSTREAM Collector (Indoor Biotechnologies Inc.). The samples were divided into aliquots of 50 mg and stored at −20°C until DNA processing.

**Figure 1.**
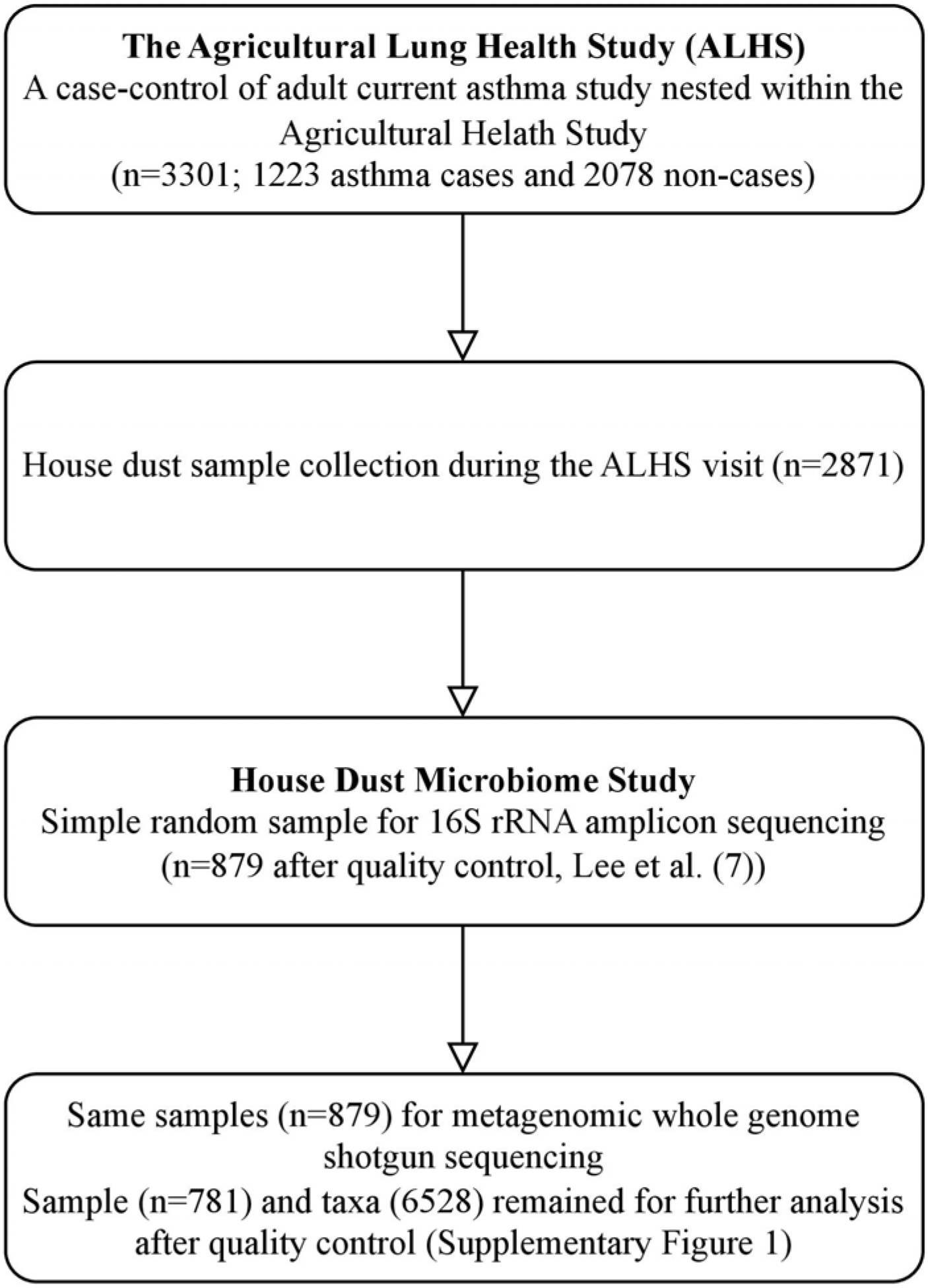
Workflow of house dust microbiome study in WGS. This workflow includes a summary of sample selection from the Agricultural Lung Health Study (ALHS) (n=3,301) to the house dust microbiome study with 16S (n=879) and WGS sequencing (n=781).

During the home visit, information was obtained on environmental factors, including current (past 12 months) farming activities (living on a farm, working with crops, and working with animals), type of animals raised on the farm (beef or dairy cattle, swine, or poultry) and the presence of indoor pets (cats and dogs). Field technicians noted the presence of carpeting in the bedroom and ranked overall home cleanliness on a standardized five-point scale (32). For our analysis, we created a binary variable comprising poor/lower (score of 1 or 2) or good/higher (score of 3–5) home condition. We categorized season of dust collection based on the date of the home visit: March 21–June 20 for spring, June 21–September 20 for summer, September 21–December 20 for fall, and December 21– March 20 for winter.

### 2.3 DNA extraction

A random selection (n=879, including 333 asthma cases) of dust samples were sent for WGS analysis (Figure 1). DNA extraction in described elsewhere (4). Briefly, DNA was isolated using a MoBio 96 well plate PowerSoil DNA extraction kit (QIAGEN Inc.), as recommended by the manufacturer, with the modification of loading 0.3-0.5g per dust sample into each well and incubated in PowerSoil bead solution and C1 buffer at 70°C for 20 min before the beating step to aid in lysis of spores. We quantified using the NanoDrop (A260) (Thermo Fisher Scientific Inc.) and normalized to 5 ng/L DNA.

### 2.4 Metagenomic whole genome shotgun sequencing and preprocessing

The University of California San Diego IGM Genomics Center performed library preparation, multiplexing, and whole genome shotgun sequencing using standard techniques (33). Extracted DNA was quantified via QubitTM dsDNA HS Assay (ThermoFisher Scientific). The library size was selected for fragments between 300 and 700 bp using the Sage Science PippinHT and sequenced as a paired-end 150-cycle run using an Illumina HiSeq2500 v2 in Rapid Run mode.

We performed several quality control steps, which are summarized in Supplementary Figure S1. We first trimmed low-quality reads, duplicates, and adapters based on FastQC results (v0.11.5) (34). We then identified and removed reads not from microbial genomes, as potential contaminant host genomic sources (human, PhiX, cow, pig, chicken, turkey, horse, goat, sheep, dog, cat, and dust mite genomes) (Supplementary Table S1) using Bowtie2 (35) and KneadData (v0.7.10) (36). We further assessed the taxonomic classification of sequences using Kraken2 (v2.1.1) (37) and obtained accurate estimations of abundance using Bracken (v2.5.0) (38) with pre-compiled data comprising RefSeq genomes for bacteria, archaea, eukaryotes, fungi, viruses, and plasmids and NCBI taxonomy information. Supplementary Tables S2 and S3 summarize the overall read sequence statistics and proportion of host genome contaminants across samples. Additionally, we accounted for the potential introduction of contaminant DNA sequences during sample collection or laboratory processing by incorporating negative ‘blank’ sequencing controls of sterile water, with contaminants identified and removed with the decontam R package (v1.10.0) (39). A total of 168 taxa were filtered out (Supplementary Table S4). Because dust samples have low microbial biomass (fewer microbes), we performed two sequencing runs, each with separate quality control processes, and then performed abundance pooling across the two runs. At the sample level, we excluded low-quality samples defined by sequencing depths less than 1000 (Supplementary Figure S2). Rare taxa were filtered out if they did not appear in at least 10 samples (Supplementary Figure S2). This quality control pipeline left 781 samples and 6,528 taxa for downstream analysis. A taxonomy chart was created that assigned all taxa to a taxonomic classification across the seven phylogenetic levels - kingdom, phylum, class, order, family, genus, and species. The Supplemental Methods provides details of the bioinformatic procedures.

### 2.5 Statistical analysis

We performed all statistical analyses and visualization in R (v4.0.3) (40). We rarefied data to the minimum library size (1,003) across all samples before calculating alpha and beta diversities using the phyloseq R package (v1.34.0) (41). We considered both non-farming exposures, including state of residence, sex, presence of indoor pets, home condition, and season of dust collection, and farming exposures in the past 12 months, including living on a farm, crop farming, and animal farming. All exposures were treated as binary variables. For season of dust collection, we compared one season to all other seasons combined. We included asthma as a covariate in all models due to the nested case-control design.

To evaluate intra-group alpha diversity and its association with farming and non-farming exposures we used the Shannon index, exponentially transformed for normality, as the outcome in linear models. We first fitted a baseline univariable regression model for each exposure to identify exposures associated with alpha diversity. We also considered whether associations differed by state of residence (IA or NC) by using product terms. Our final multivariable model included any exposure with significant association to alpha diversity from the baseline univariable model, along with any significant product terms for the individual interactions of each exposure with state of residence. Detailed analytical formula were described in Supplemental methods (SM3). We set p<0.05 as the statistical significance threshold for all analyses.

To explore beta diversity, we calculated unweighted and weighted UniFrac distance metrics. We conducted permutational multivariate analysis of variance (PERMANOVA) analysis to test the differences in microbial community structure across exposure levels using the *adonis* method in the R vegan package (v2.5.7) (42, 43). We used the R^2^ value to quantify the percentage of variance explained. We did similar analysis as alpha diversity to evaluate differences in associations by state. We conducted non-metric multidimensional scaling (NMDS) analysis to visualize the separation between samples by exposure levels in a two-dimensional space using the phyloseq (v1.34.0) (41) and R ggplot2 (v3.3.6) (44) packages.

To identify differentially abundant taxa for each exposure, we used analysis of composition of microbiomes with bias correction (ANCOM-BC, v1.0.5) models (45), which is based on a linear regression framework on the log transformed taxa counts, with exposures as dependent variables and sampling fraction as an offset term. To account for variation in sequencing depth, we performed normalization by estimating the sampling fraction using the ANCOM-BC built-in algorithm. We tested taxa at the OTU level and summarized the results by genus and phylum rank. We also calculated the log2 fold-difference which is the ratio of the mean abundance after normalized by ANCOM-BC across exposure levels. We controlled the false discovery rate (FDR) at 0.05 with the Benjamini-Hochberg (BH) method (46). We determined a taxon to be significantly differentially abundant if it had both p<0.05 after FDR correction and had log2 fold-difference larger than 1 or smaller than -1. We performed sensitivity analyses to evaluate differences in associations by state of residence.

Lee et al. (47) analyzed samples for the same population with 16S rRNA amplicon sequencing. To examine differences of house dust microbial profile between these two methods, we compared the taxonomic chart from our WGS data to the previous 16S data to determine the number of unique and overlapping microbial organisms, at the phyla rank, detected by each sequencing method. We note how common or rare the uniquely identified phyla were based on the frequency of assigned taxa and the relative abundance across samples. In addition, we evaluated the differences between alpha diversities (richness and Shannon index) generated by the two sequencing methods by calculating the Spearman’s correlation coefficient.

## 3 Results

### 3.1 Summary statistics for the study population and metagenomics characteristics

Table 1 summarizes the demographic characteristics and environmental exposures of the study population. Iowa residents accounted for 68% of samples; North Carolina for 32% (247). Sixty percent of participants were male. Indoor pets (dogs or cats) were present in 43% of homes. Most homes (78%) were in good/higher cleanness, and nearly all had carpeted floors (93%). Overall, 83% of participants lived on a farm, 56% farmed crops in the past 12 months, and 51% worked with farm animals in the past 12 months. Of the 401 (51%) participants who reported animal farming, 281 worked with beef cattle, 48 worked with dairy cattle, 120 worked with hogs, and 90 worked with poultry. Overall, 31% of dust samples were obtained in summer, 25% in spring, 20% in fall, and 23% in winter. Current asthma was present in 296 (37.9%) participants and the overall mean age of participants was 62 years (standard deviation 11).

**Table 1.**
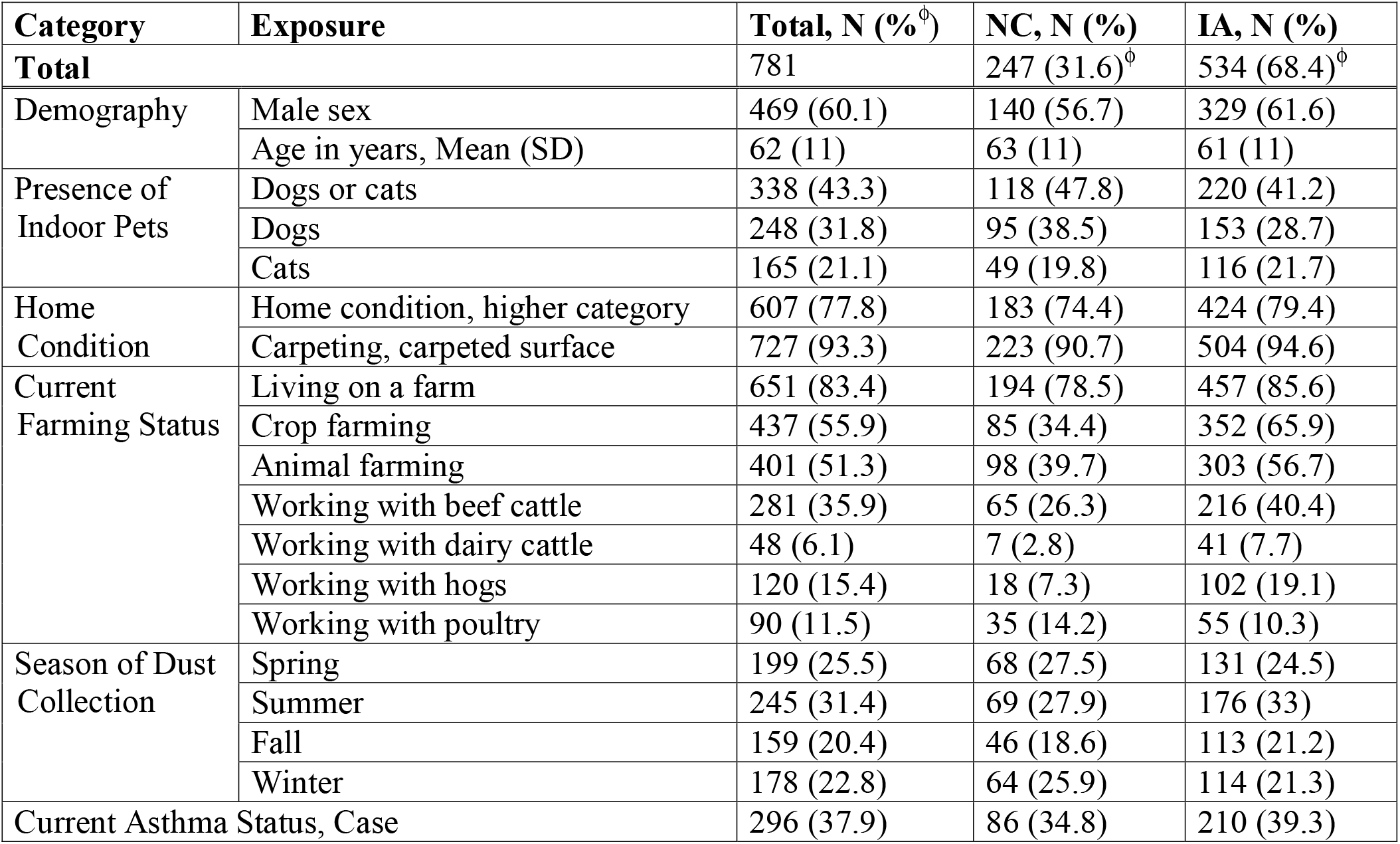
Characteristics of Study Population. ^ϕ^ percentage based on full cohort versus within each state. Exposures that were different by state of residence using Pearson’s chi-squared test (p<0.05): Dogs, Living on a Farm, Crop Farming, Animal Farming, Working with Beef Cattle, Working with Dairy Cattle, Working with Hogs.

After filtering out samples with low sequencing depth and filtering out rare taxa, 781 samples and 6,528 taxa remained for downstream analysis with 183,025,561 reads across all samples. At the Kingdom phylogenetic level, 5,661 taxa were assigned to Bacteria, 156 to Archaea, 96 to Eukaryota, and 615 to viruses, with an average of 2,247 (±1,226) taxa per sample (n=781). Figure 2 outlines the phylum composition across all samples. Among the 59 phyla identified from WGS, 16 had relative abundance greater than 1% in at least one sample (Figure 2, Supplementary Table S5). Phyla *Firmicutes, Proteobacteria, Actinobacteria*, and *Bacteroidetes* were the most prominent among home dust microbial communities. At lower taxonomy rank, 1789 unique genera were identified, where 36 had relative abundance greater than 10% in at least one sample. The five most abundant genera were *Mycobacterium, Serratia, Toxoplasma, Lactobacillus*, and *Alcaligenes* (Supplementary Table S6).

**Figure 2.**
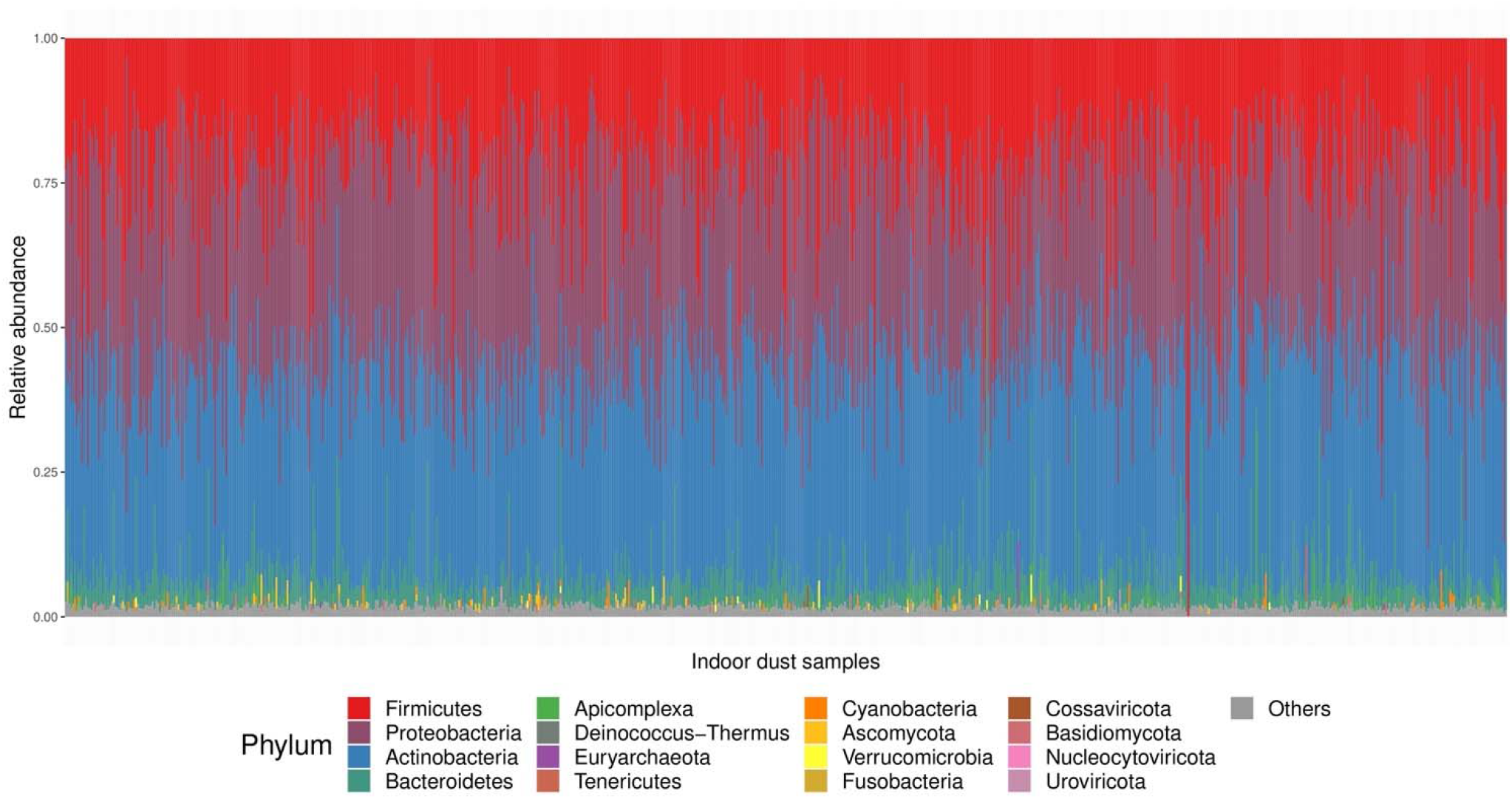
Relative abundance at the phylum level across all home dust samples. The 16 phyla with relative abundance greater than 1% in at least one sample are color-coded according to the legend. All other phyla are represented in grey.

### 3.2 House dust microbial community diversity analysis

Figure 3 shows the association between alpha diversity and each exposure. The presence of indoor pets and farming status (living on a farm, crop farming, animal farming with beef cattle, hogs, and poultry) were positively associated with alpha diversity, while good/higher home cleanliness was negatively associated with alpha diversity (p<0.050). State of residence had a suggestive significant association with alpha diversity with p=0.057. In our multivariable primary model including all statistically significant exposures and all significant interaction terms with state of residence, living on a farm and animal farming remained significantly positively related to alpha diversity (Supplementary Table S7).

**Figure 3.**
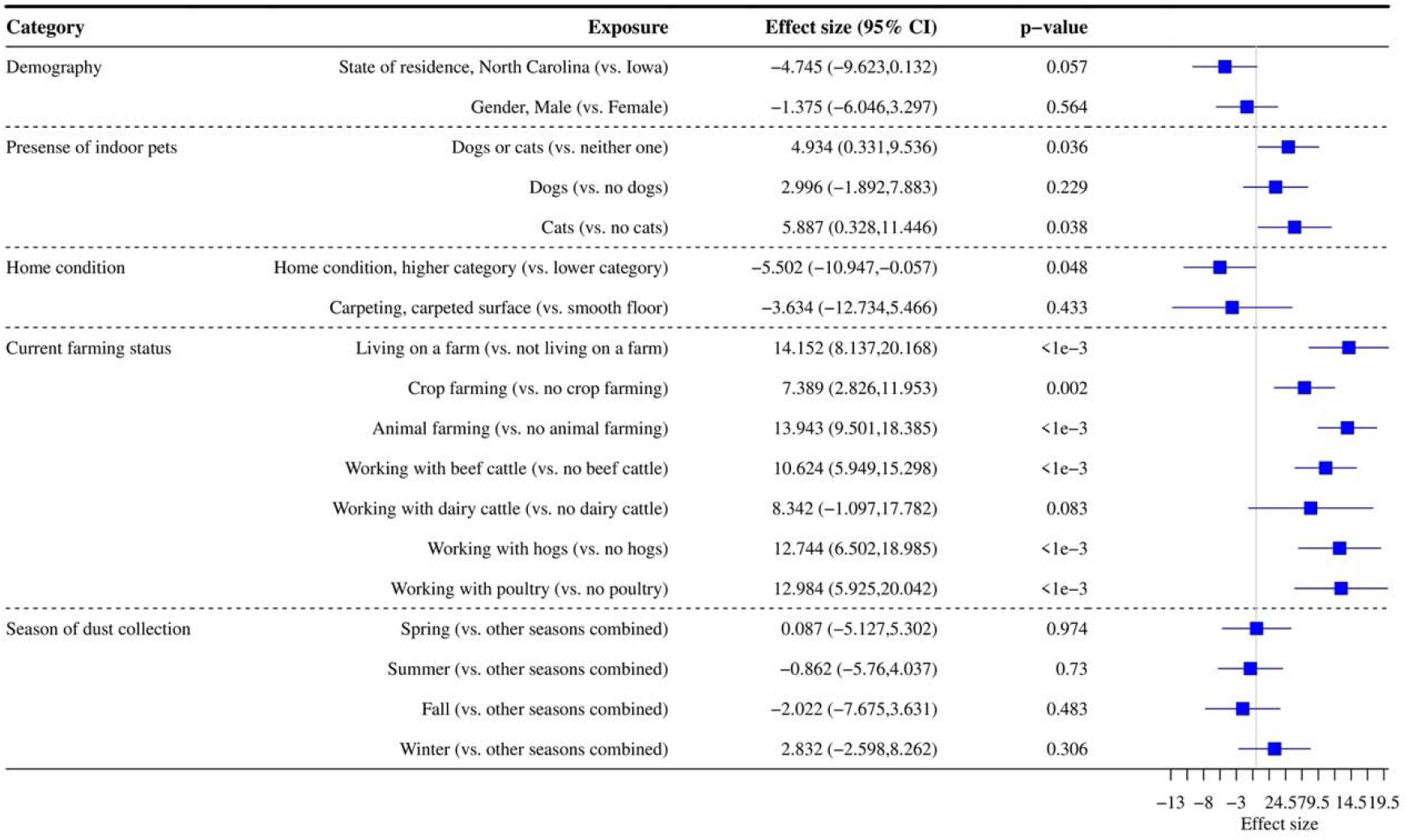
Association between exposures and alpha diversity (Shannon index with exponential transformation). Data were rarefied to the minimum library size (1,003) across all samples. Effect size refers to the coefficient from the regression model (difference in alpha diversity for yes versus no for each exposure). The 95% confidence interval (CI) and p-value for each exposure from the regression model are reported.

For beta-diversity, PERMANOVA analysis revealed significant differences in beta diversity for all demographic characteristics and exposure levels based on unweighted UniFrac distance although the percent variance explained by the exposure groups (R^2^ values) were small (Supplementary Figure S3). Current farming accounted for relatively greater explained microbial diversity variance (0.5% for crop farming and 0.7% for animal farming) compared to other farm and nonfarm exposures (Figure 4a, 4b). The differences in the microbial composition of home dust samples by state of residence explained around 1% of the variance of bacterial communities (p=0.001) (Figure 4c). The results with weighted UniFrac distance were similar to unweighted metric (Supplementary Figure S4).

**Figure 4.**
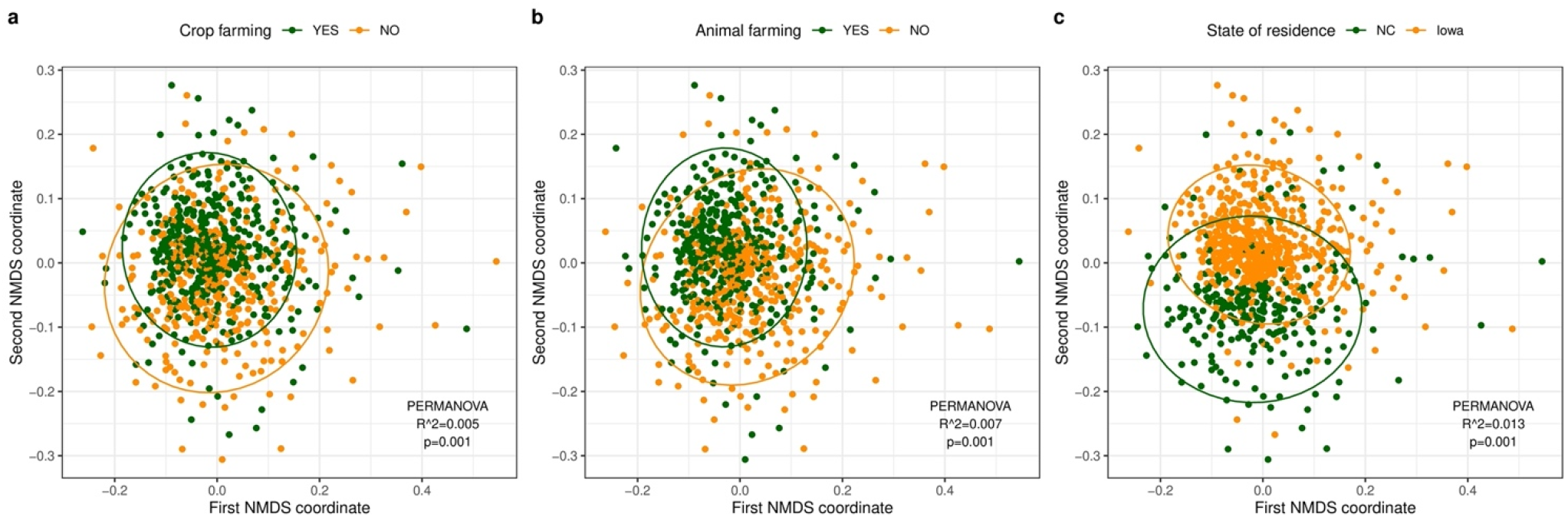
Non-metric multidimensional scaling (NMDS) analysis based on unweighted UniFrac distances for samples with different exposure levels. **(a)** Crop farming (green: with crop farming, yellow: without crop farming). **(b)** Animal farming (green: with animal farming, yellow: without animal farming). **(c)** State of residence (green: North Carolina (NC), yellow: Iowa). The dust microbial community of each sample is represented by a single dot. The ellipse represents the 95% confidence interval for the centroids of each exposure level. R^2^ values (percentage of variance explained by an exposure) and p-values from the PERMANOVA analysis are reported.

### 3.3 Differential abundance analysis of individual taxa

There were 372 unique taxa belonging to 175 genera within 16 unique phyla, that were differentially abundant in relation to at least one exposure (Supplementary Table S8, Supplementary Table S9, Supplementary Figure S5). Animal farming and living on a farm were associated with more differentially abundant taxa than non-farming exposures. Figure 5 includes volcano plots of differentially abundant taxa related to the presence of indoor pets, living on a farm, crop farming, and animal farming in the past 12 months, color coded by phylum. The top 10 taxa based on FDR values are labeled by their genus rank. Working with hogs was identified with the greatest number of differentially abundant taxa compared with other types of farming animals (Figure 5a, Supplementary Figure S5).

**Figure 5.**
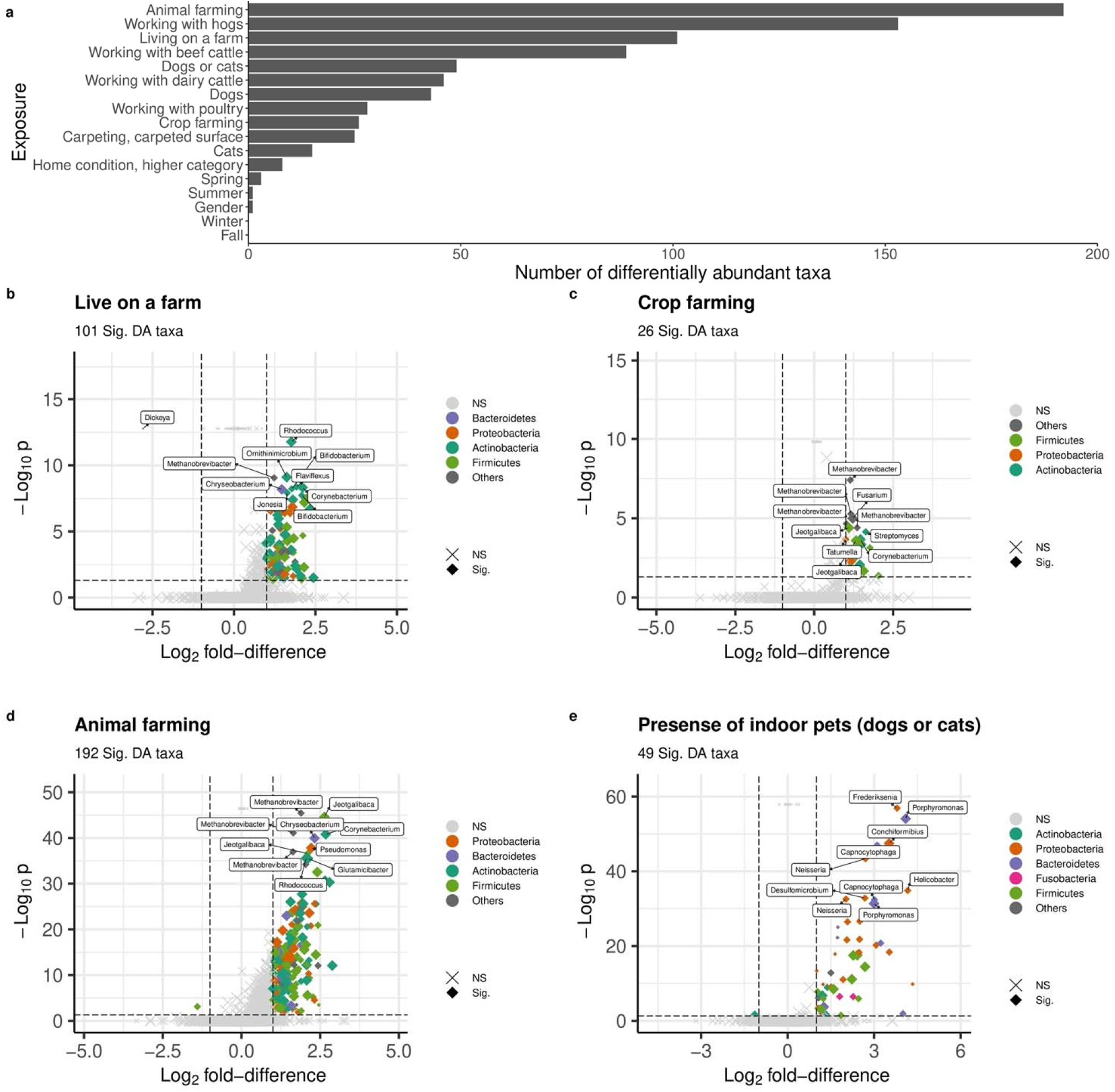
Differentially abundant (DA) taxa related to individual exposure (FDR<0.05). **(a) Number of DA taxa. (b)-(e) Volcano plot for (b)** Presence of indoor pets, **(c)** Living on a farm, **(d)** Crop farming, and **(e)** Animal farming. DA taxa are colored by phylum. The top 10 DA taxa with the smallest adjusted p-values are labeled by genus. Dot size indicates the medium abundance level for each taxon. a Benjamini-Hochberg method is used for FDR correction. lfd: log2 fold-difference. Vertical and horizontal dash lines indicate the threshold of p value after FDR correction and lfd for filtering DA taxa. Sig: DA taxa with p<0.05 after FDR correction (i.e., log10 p<0.5) and lfd>1 (or ldf<-1); NS: non-DA taxa.

Living on a farm was associated with differential abundance of 101 taxa (increased abundance for 100 taxa and decreased abundance for one taxon in genus *Dickeya*), which were mainly in phylum *Actinobacteria, Bacteroidetes, Firmicutes*, and *Proteobacteria* (Figure 5b). Among the top 10 taxa, two were in genus *Bifidobacterium*. The 26 differentially abundant taxa all had increased abundance related to crop farming were mainly in phyla *Actinobacteria, Firmicutes*, and *Proteobacteria* (Figure 5c). The most significant taxa were genus *Methanobrevibacter* and *Jeotgalibaca*. Animal farming was associated with increased abundance for 191 taxa and decreased abundance for one taxon in phylum *Firmicutes* (Figure 5d). Genera *Methanobrevibacter, Jeotgalibaca, Corynebacterium, Chryseobacterium, Glutamicibacter, Pseudomonas*, and *Rhodococcus* were among the top 10 taxa. Forty-nine taxa were differentially abundant for the presence of indoor pets, mostly in phylum *Actinobacteria, Bacteroidetes, Firmicutes, Fusobacteria*, and *Proteobacteria* (Figure 5e). The taxa with the smallest FDR value were genus *Frederiksenia* and *Poerphyromonas*. Only a few differentially abundant taxa belonging to phylum *Proteobacteria* were related to the season of dust collection (Supplementary Table S8, Supplementary Figure S5).

Many differentially abundant taxa were shared among exposures, but there were some taxa uniquely related to individual farming exposures (Figure 6, Supplementary Table S9). In particular, there were 103 taxa assigned to 67 genera within 7 phyla (*Proteobacteria, Actinobacteria, Bacteroidetes, Euryarchaeota, Firmicutes, Tenericutes, Chloroflexi*) specific to animal farming. For crop farming, 2 taxa were unique – *Tatumella citrea* in phylum *Proteobacteria* and *Fusarium graminearum* in phylum *Ascomycota* (Supplementary Table S9). There were only 4 taxa (*Bacillus* [*Firmicute* phyla], *Campylobacter* [*Proteobacteria*], *Streptomyces* [*Actinobacteria*], and *Acholeplasma* [*Tenericutes*]) that were identified to be associated with both animal farming and crop farming (Supplementary Table S9). In terms of specific type of farm animals, 89 taxa were unique to hogs, including *Clostridium, Campylobacter, Pseudomonas*, and *Streptococcus suis*, 14 unique to poultry, including *Enterococcus, Brucella*, and *Escherichia* genera, 5 unique to dairy cattle, including *Mycoplasma* and *Acinetobacter*, and 26 unique to beef cattle, including *Corynebacterium* and *Bacillus* (Supplementary Table S9). Several taxa were identified in multiple types of farming animals: 15 taxa were shared for hogs, beef cattle and dairy cattle, only one taxon (*Carnobacterium sp*.*_CP1*) were common among hogs, poultry, and beef cattle, and 24 taxa including *Methanobrevibacterium* was related to either cattle type (Figure 6, Supplementary Table S9).

**Figure 6.**
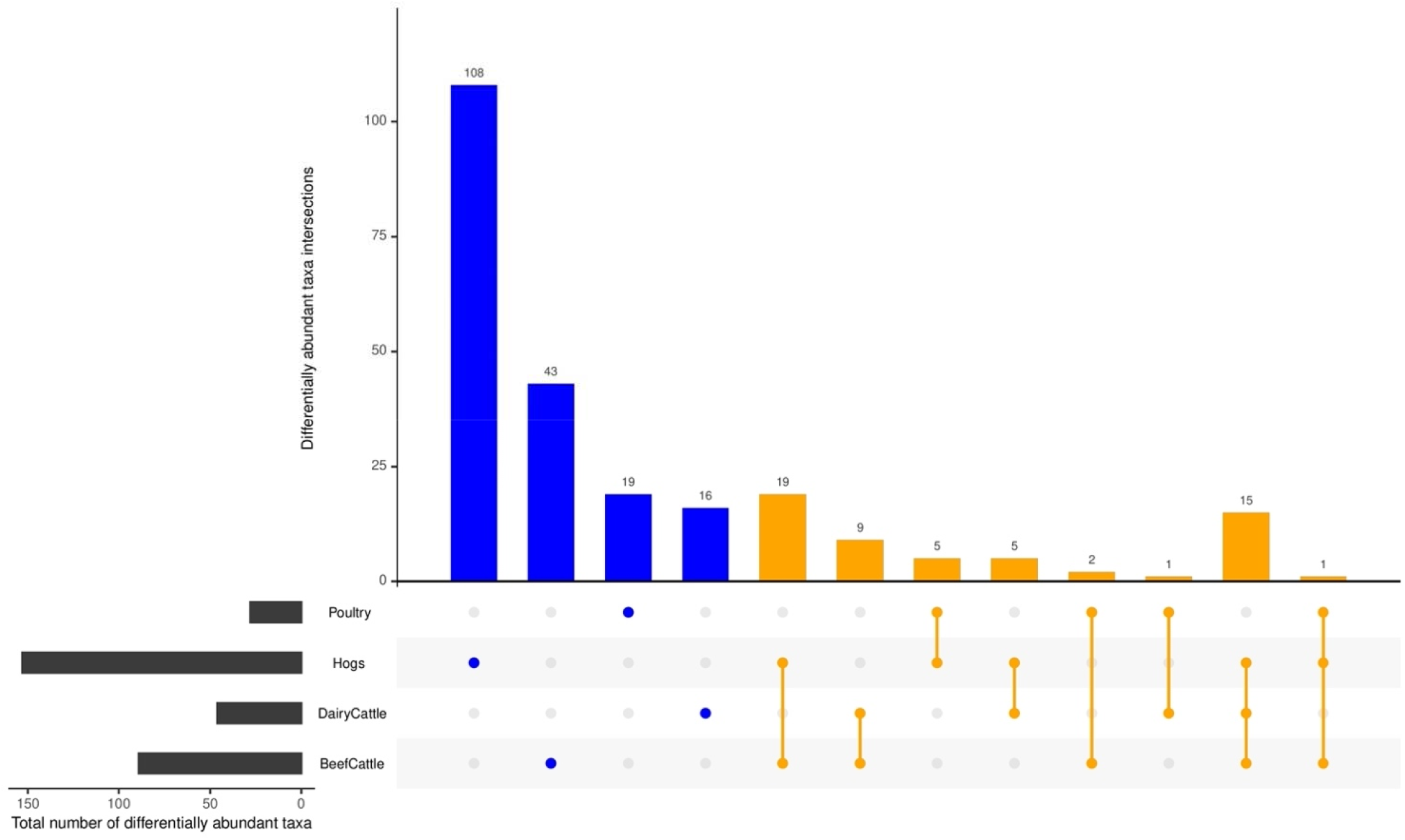
Differentially abundant taxa related to various types of farming animal (FDR<0.05). Commonly identified differentially abundant taxa shared by farming animal types were aligned by lines (orange), while differential taxa unique to farm animal type is identified by a single dot (blue).

As for non-farming exposures, 44 taxa were uniquely differentially abundant for presence of indoor pets, including animal-related *Staphylococcus* species *pseudintermedius* and *felis*. Additionally, 4 taxa were unique to home condition, 16 unique to carpeting, and 3 unique to spring dust collection (Supplementary Table S9).

### 3.4 Sensitivity Analysis by State of Residence

For interaction effects by state of residence with either alpha or beta diversity, only sex, home condition, crop farming, general animal farming, beef cattle farming, and spring dust collection had significant interactions, but most effect sizes were minimal (Supplementary Table S10, Supplementary Table S11, Supplementary Table S12). Therefore, we did not carry interaction products into the differential abundance analysis. When stratifying by state of residence, several exposures, including the presence of indoor pets, living on a farm, and general animal farming, were significantly associated with either alpha or beta diversity in Iowa, where about 2/3 of participants resided but not in North Carolina which has a much smaller sample size (Supplementary Table S13, Supplementary Table S14). Fourteen phyla were consistent for both states with differentially abundant taxa by at least one exposure (Supplementary Table S8, Supplementary Figure S6, Supplementary Figure S7).

### 3.5 Additional findings with WGS from 16S r RNA sequencing results

WGS data identified many more taxa and phyla than 16S rRNA. The 6,526 taxa identified by WGS data were assigned to 59 phyla, compared to 1,346 taxa from 18 phyla for 16S. The three phyla with the largest proportion of taxa assignment (most frequent) for WGS results (*Proteobacteria, Actinobacteria, Firmicutes*) were identical for 16S results. Among the 18 phyla identified from 16S sequencing, 17 were present in the WGS results (Figure 7, Supplementary Table S5). 47 phyla were uniquely identified by WGS, of which the most frequent phyla were *Uroviricota* with 518 (7.9%) taxa assigned, *Ascomycota* with 51 (0.8%) taxa assigned, *Spirochaetes* with 38 (0.6%) taxa assigned, *Cossaviricota* with 35 (0.5%) taxa assigned, and *Apicomplexa* with 25 (0.4%) taxa assigned (Supplementary Table S5). Additionally, many of the unique phyla in WGS were not rare, including *Apicomplexa* with average relative abundance across all samples at 3%, and *Ascomycota, Cossaviricota, Basidiomycota, Nucleocytoviricota*, and *Uroviricota* at 2% each (Supplementary Table S5). When examining differences in the alpha diversity of results from WGS and 16S sequencing, Spearman’s correlation coefficient for richness (rho=0.413, p< 2.2e-16) and the Shannon index (rho=0.355, p< 2.2e-16) were moderate.

**Figure 7.**
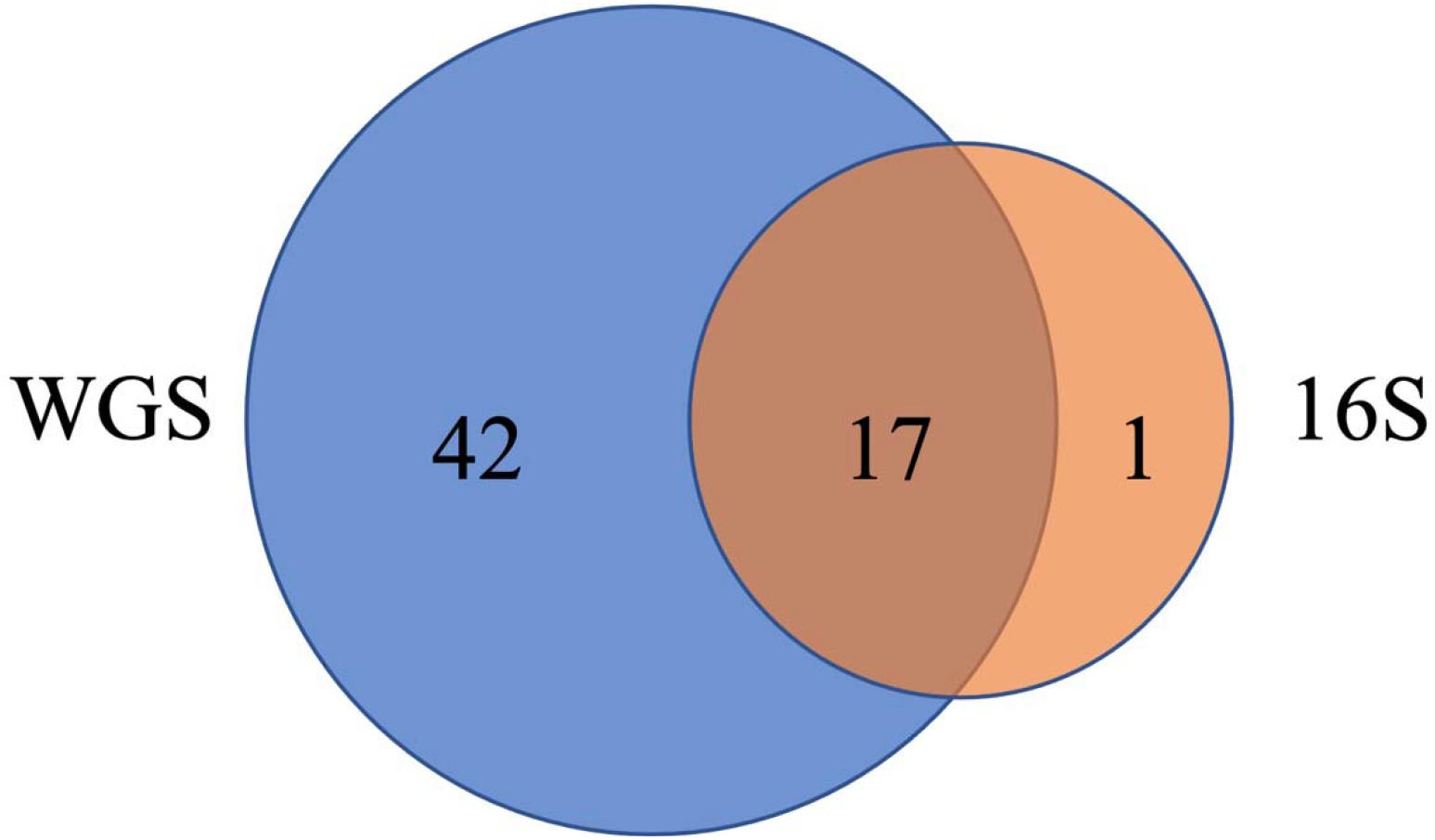
Venn diagram of the number of phyla identified in WGS (blue) and 16S (orange). 17 phyla were identified by both methods (Supplementary Table S14).

Because more microbial organisms were detected by WGS, we observed additional associations with farming exposures compared to 16S data presented by Lee et al. (39). Notably, a unique phylum (*Ascomycota)* detected only by WGS was significantly associated with crop farming. One of phyla identified by both WGS and 16S (*Tenericutes)* had differentially abundant taxa based on animal farming using WGS not with 16S (Supplementary Table S5, Supplementary Table S8). In addition, WGS provided the ability to assign taxa to genus taxonomic levels, including the 175 genera with differential abundance taxa related to at least one exposure (Supplementary Table S8), compared to 16S results at the phyla and family level. Of 175 genera, 16 had relative abundance greater than 10% in at least one sample including *Lactobacillus, Staphylococcus*, and *Bacillus* (Supplementary Table S6, Supplementary Table S8).

## 4 Discussion

In this study, we evaluated the associations between farming exposures and house dust microbiota using the whole genome shotgun sequencing method in a US agricultural population. Our results indicate that both indoor microbial diversity and composition in homes differ in relation to current farming exposures; living on a farm, and crop and animal farming were associated with increased within-sample microbial diversity levels and altered microbial composition. Expanding on our previous findings performed with 16S rRNA gene amplicon sequencing, we identified four times more unique microbial taxa. The improved detection of unique taxa with WGS enabled us to detect novel associations between farm exposures and increased abundance of specific microbes including *Rhodococcus, Bifidobacterium, Corynebacterium*, and *Pseudomonas*. Enhanced identification of factors that impact the indoor microbiome can improve understanding of environmental exposure pathways relevant to human health.

A unique aspect of this study was the use of the whole genome shotgun sequencing technique, compared to many previous home dust microbiome studies that use the 16S rRNA amplicon sequencing technique (4, 12). This work is the first reported to use WGS to evaluate farm exposures in home dust microbiota. WGS has the advantage of sequencing the entire microbial genome, versus just a single gene, which can more accurately assign taxonomic classifications (48). In this study, the use of WGS identified more unique microbial phyla – 42 phyla were found only using WGS, including both common and rare taxa, versus only one phylum using the 16S technique. Detection of a greater number of unique phyla from WGS compared to 16S enables better characterization of the mixed, complex microbial composition of indoor dust in homes. Consequently, we observed novel environmental exposure associations with the newly detected microbial outcomes from this more comprehensive WGS method. Expanded taxonomic detection and depiction, as well as the development of updated, robust bioinformatic and statistical tools for metagenomic data (49), will then have downstream effects on the interpretation of association to environmental exposures.

Consistent with findings using 16S, our data with WGS found that numerous bacteria were associated with environmental exposures across various phyla. At the phyla level, *Actinobacteria, Bacteroidetes, Firmicutes*, and *Proteobacteria* were positively associated with farm exposures, including living on a farm and crop and animal farming. These trends are similar to our findings using 16S, which found *Firmicutes* and *Proteobacteria* to be associated with farm exposures. In previous research, these phyla have been associated with various health conditions, such as asthma, atopy, and cardiometabolic outcomes (50-52). However, our 16S findings found that crop farming was associated with significant decreased abundance of taxa in 16 of the 19 phyla (4), compared to using WGS, where all 26 of our significantly associated taxa had an increased abundance with crop farming. Complementary studies evaluating home dust in Germany and Finland (12) and classroom dust in China (53) have found positive associations between nearby farm exposure and increased abundance of *Proteobacteria* (also known as *Alphaproteobacteria*) and *Actinobacteria*.

WGS enables improved classification of microbial taxa at lower taxonomic levels, including the identification of genera that are differentially abundant by environmental exposures. Using WGS, we ascertained genera that were associated with our farming exposures, including *Rhodococcus, Bifidobacterium, Corynebacterium*, and *Pseudomonas. Rhodococcus* and *Corynebacterium*, gram-positive bacteria, and *Pseudomonas*, a gram-negative bacterium, are found commonly in environmental sources (54-56). Certain strains of each can be pathogenic in immunocompromised individuals (54-56), and their abundance has been shown to be elevated in dust from children with asthma and atopy (57). *Pseudomonas* was also found to be increased using WGS in classroom dust samples in rural regions near farms compared to suburban areas in China (53). Interestingly, *Rhodococcus, Pseudomonas*, and *Methylobacterium* (another microbe positively associated with farm exposures in our data) have been previously identified in agricultural settings, where they can be bioremediation agents and degrade certain pesticides (58). *Bifidobacterium* is ubiquitous in the human and animal gastrointestinal tract and is associated with positive gut homeostasis, inhibition of pathogen colonization, and modulation of the local and systemic immune system (59, 60). We observed that *Methanobrevibacter* and *Jeotgalibaca*, both previously associated with cattle rumen and manure fermentation (61, 62), were increased with crop and animal farming, and unique to dairy and beef cattle farming, which is consistent with previous studies evaluating farm exposures in human microbial communities (12, 63, 64). Two taxa unique to crop farming, *Tatumella citrea* and *Fusarium graminearum*, are pathogens associated with grain production (65, 66). Reassuringly, we noted an increased abundance of microbes specific to farm and companion animals associated with concurrent exposure to those animals, such as *Streptococcus suis* with hog farming exposure (67) and *Staphylococcus pseudintermedius* and *felis* with dog and cat exposure (68, 69).

Our findings suggest that the home dust microbial diversity levels differ between participants exposed to farming activities, as well as pets, both for alpha and beta diversity levels. Overall, the findings from this study were generally similar to those preformed previously using 16S (4). For microbial composition beta diversity, we found distinct microbial community structure based on farm and non-farm exposures, which was significant for all explored variables, similar to results from 16S. The coefficient-of-determination R-squared (R2) statistic was greater using 16S, which supports the hypothesis that WGS resulted in more diverse microbial community identification with greater heterogeneity, so the same exposure would account for less of the variability. Both WGS and 16S findings had low R2 explained variance, consistent with previous research (70). Both analyses showed positive associations between alpha diversity and crop and animal farming. Living on a farm was a significant factor using WGS but not 16S. In addition, there were differences based on the type of animal production, with hog production having a positive association using WGS but not 16S, and dairy cattle production having a positive association using 16S but not WGS (although there was a positive trend).

The differences in associations between exposures and Shannon alpha diversity in the WGS compared to our previous 16S data are to be expected given differences between the methods and batch effects when comparing two different methods run three years apart in different laboratories. Alpha diversity was slightly higher in WGS than 16S samples with moderate correlation (Spearman’s rho=0.36); unsurprisingly, as a greater number of unique microbes were identified with WGS and is similar to previous research on environmental samples (19). The discrepancies in measurements and effect sizes between WGS and 16S can lead to altered interpretations regarding risk factors for dysbiosis in home dust microbial composition and highlights the importance of how the processing of microbiome samples can impact downstream analyzes.

The positive associations with farm exposures and alpha diversity reinforce trends observed in other literature (3, 10, 12, 14, 53), in addition to our prior 16S analyses (4). In a study of 203 homes in Finland and Germany, homes located on farms had significantly higher indoor microbial richness and diversity compared to rural non-farm home indoor dust, which was associated with decreased asthma risk in child inhabitants (12). Amin et.al. reported that airborne bacterial diversity was more abundant in farmer’s indoor environment than in suburban homes (10). Using WGS, a study in Shanxi Providence, China, found higher microbial diversity in schools in rural area near farms compared to urban non-farm schools (53).

A limitation of this work is that we only have a single dust sample per household, collected in the bedroom. Thus, we assume the sample reflects the normal home condition. To the extent that microbial composition differs across the household (11), this may not be true. However, people spend about a third of their time in the bedroom, making this a logical single sampling location. This limitation would be expected to lead to nondifferential misclassification of exposure and a bias toward the null. Our work benefits from an advanced next-generation technique, whole genome shotgun sequencing, to explore the impact of detailed farm exposures on the indoor microbiome in a large sample size compared to previous studies. The improved detection from WGS across novel phyla at the genus level adds insights on factors influencing the built environment microbiota, which plays a key component on host microbiome composition and subsequent health outcomes. Future investigations on the functional capabilities of the dust microbiota, such as presence of antibiotic resistance genes, can help better understand human health and disease etiology caused by environmental exposures.

## 5 Conclusions

We evaluated a comprehensive set of factors related to farming to determine their influence on home dust microbiome assessed using state of the art whole genome shotgun sequencing. The increased identification by WGS of microbial entities led to detection of associations missed using older 16S technology. Identifying significant predictors of indoor built environmental microbiota is an important element in understanding environmental exposure health pathways. The use of advanced whole genome shotgun sequencing techniques produced novel insights into these health pathways and may be considered an optimal metagenomic method for future environmental health studies.

## Supporting information

Supplemental contents

## Data Availability

The raw sequencing data presented in this study are stored in online platform Qiita. Contact the corresponding author for permission for the raw sequence data and metadata.

## 6 Conflict of Interest

The authors declare that the research was conducted in the absence of any commercial or financial relationships that could be construed as a potential conflict of interest.

## 7 Author Contributions

SL and ML were responsible for study design and data acquisition. CP and LF initiated ALHS study and were responsible for the sample collection. ZW designed and performed all bioinformatics and statistical analysis, with SZ, AM, KD, SL and ML providing analytical input. QZ, AG and RK planned shotgun metagenomics sequencing and prepared raw sequences data. ZW and KD formulated the research ideas and drafted the manuscript. All authors contributed to the interpretation of results and editing of the manuscript.

## 8 Funding

This work was supported by the Intramural Research Program of the National Institutes of Health (NIH), the National Institute of Environmental Health Sciences (NIEHS) (Z01-ES049030 and Z01-ES102385), the National Cancer Institute (Z01-CP010119B), and by American Recovery and Reinvestment Act funds.

## 9 Ethics

The Institutional Review Board at the National Institute of Environmental Health Sciences approved the study. Written informed consent was obtained from all participants.

## 10 Acknowledgments

This work was supported by the Intramural Research Program of the National Institutes of Health (NIH), the National Institute of Environmental Health Sciences (NIEHS) (Z01-ES049030 and Z01-ES102385), the National Cancer Institute (Z01-CP010119B), and by American Recovery and Reinvestment Act funds. The Center for Microbiome Innovation at the University of California San Diego provided support by generating sequencing data. We appreciate all the study participants for their contribution to this research. We thank Drs. F. Day of NIEHS for expert computational assistance and J. Hoppin (North Carolina State University, Raleigh, NC) for her important contribution to the Agricultural Lung Health Study during her tenure at NIEHS. We thank Dr. Gail Ackermann of UCSD for assistance with metadata curation, and Dr. Greg Humphrey for laboratory processing.

## 11 Supplementary Material

See Supplemental Materials Table of Contents document for a list of the supplementary methods, tables, and figures referenced.

## Notes

### Competing Interest Statement

The authors have declared no competing interest.

### Author Declarations

The Institutional Review Board at the National Institute of Environmental Health Sciences gave ethical approval for this work. Written informed consent was obtained from all participants.

## References

1. U.S. Environmental Protection Agency. Report to Congress on indoor air quality. 1989. Contract No.: EPA/400/1-89/001C.

2. Ege MJ, Mayer M, Normand AC, Genuneit J, Cookson WO, Braun-Fahrländer C, et al. Exposure to environmental microorganisms and childhood asthma. N Engl J Med. 2011;364(8):701–9.

3. Stein MM, Hrusch CL, Gozdz J, Igartua C, Pivniouk V, Murray SE, et al. Innate Immunity and Asthma Risk in Amish and Hutterite Farm Children. New England Journal of Medicine. 2016;375(5):411–21.

4. Lee MK, Carnes MU, Butz N, Azcarate-Peril MA, Richards M, Umbach DM, et al. Exposures Related to House Dust Microbiota in a U.S. Farming Population. Environ Health Perspect. 2018;126(6).

5. Dannemiller KC, Gent JF, Leaderer BP, Peccia J. Indoor microbial communities: Influence on asthma severity in atopic and nonatopic children. Journal of Allergy and Clinical Immunology. 2016;138(1):76-83.e1.

6. Gupta S, Hjelmsø MH, Lehtimäki J, Li X, Mortensen MS, Russel J, et al. Environmental shaping of the bacterial and fungal community in infant bed dust and correlations with the airway microbiota. Microbiome. 2020;8(1):115.

7. Lax S, Smith DP, Hampton-Marcell J, Owens SM, Handley KM, Scott NM, et al. Longitudinal analysis of microbial interaction between humans and the indoor environment. Science. 2014;345(6200):1048–52.

8. Dannemiller KC, Gent JF, Leaderer BP, Peccia J. Influence of housing characteristics on bacterial and fungal communities in homes of asthmatic children. Indoor Air. 2016;26(2):179–92.

9. Panthee B, Gyawali S, Panthee P, Techato K. Environmental and Human Microbiome for Health. Life. 2022;12(3):456.

10. Amin H, Santl-Temkiv T, Cramer C, Vestergaard DV, Holst GJ, Elholm G, et al. Cow Farmers’ Homes Host More Diverse Airborne Bacterial Communities Than Pig Farmers’ Homes and Suburban Homes. Front Microbiol. 2022;13:883991.

11. Zhou JC, Wang YF, Zhu D, Zhu YG. Deciphering the distribution of microbial communities and potential pathogens in the household dust. Sci Total Environ. 2023:162250.

12. Kirjavainen PV, Karvonen AM, Adams RI, Täubel M, Roponen M, Tuoresmäki P, et al. Farm-like indoor microbiota in non-farm homes protects children from asthma development. Nature Medicine. 2019;25(7):1089–95.

13. Lee MK, Wyss AB, Carnes MU, Richards M, Parks CG, Beane Freeman LE, et al. House dust microbiota in relation to adult asthma and atopy in a US farming population. Journal of Allergy and Clinical Immunology. 2021;147(3):910–20.

14. Birzele LT, Depner M, Ege MJ, Engel M, Kublik S, Bernau C, et al. Environmental and mucosal microbiota and their role in childhood asthma. Allergy. 2017;72(1):109–19.

15. Breitwieser FP, Lu J, Salzberg SL. A review of methods and databases for metagenomic classification and assembly. Briefings in Bioinformatics. 2019;20(4):1125–36.

16. Laudadio I, Fulci V, Palone F, Stronati L, Cucchiara S, Carissimi C. Quantitative Assessment of Shotgun Metagenomics and 16S rDNA Amplicon Sequencing in the Study of Human Gut Microbiome. OMICS. 2018;22(4):248–54.

17. Campanaro S, Treu L, Kougias PG, Zhu X, Angelidaki I. Taxonomy of anaerobic digestion microbiome reveals biases associated with the applied high throughput sequencing strategies. Scientific Reports. 2018;8(1).

18. Fouhy F, Clooney AG, Stanton C, Claesson MJ, Cotter PD. 16S rRNA gene sequencing of mock microbial populations-impact of DNA extraction method, primer choice and sequencing platform. BMC Microbiology. 2016;16(1).

19. Tessler M, Neumann JS, Afshinnekoo E, Pineda M, Hersch R, Velho LFM, et al. Large-scale differences in microbial biodiversity discovery between 16S amplicon and shotgun sequencing. Sci Rep. 2017;7(1):6589.

20. Durazzi F, Sala C, Castellani G, Manfreda G, Remondini D, De Cesare A. Comparison between 16S rRNA and shotgun sequencing data for the taxonomic characterization of the gut microbiota. Sci Rep. 2021;11(1):3030.

21. Ranjan R, Rani A, Metwally A, McGee HS, Perkins DL. Analysis of the microbiome: Advantages of whole genome shotgun versus 16S amplicon sequencing. Biochem Biophys Res Commun. 2016;469(4):967–77.

22. Clooney AG, Fouhy F, Sleator RD, O’ Driscoll A, Stanton C, Cotter PD, et al. Comparing Apples and Oranges?: Next Generation Sequencing and Its Impact on Microbiome Analysis. PLOS ONE. 2016;11(2):e0148028.

23. Tedersoo L, Anslan S, Bahram M, Põlme S, Riit T, Liiv I, et al. Shotgun metagenomes and multiple primer pair-barcode combinations of amplicons reveal biases in metabarcoding analyses of fungi. MycoKeys. 2015;10:1–43.

24. Chan TF, Ji KM, Yim AK, Liu XY, Zhou JW, Li RQ, et al. The draft genome, transcriptome, and microbiome of Dermatophagoides farinae reveal a broad spectrum of dust mite allergens. J Allergy Clin Immunol. 2015;135(2):539–48.

25. Logares R, Sunagawa S, Salazar G, Cornejo-Castillo FM, Ferrera I, Sarmento H, et al. Metagenomic 16S rDNA Illumina tags are a powerful alternative to amplicon sequencing to explore diversity and structure of microbial communities. Environmental Microbiology. 2014;16(9):2659–71.

26. Guo J, Cole JR, Zhang Q, Brown CT, Tiedje JM. Microbial Community Analysis with Ribosomal Gene Fragments from Shotgun Metagenomes. Appl Environ Microbiol. 2016;82(1):157–66.

27. Poretsky R, Rodriguez RL, Luo C, Tsementzi D, Konstantinidis KT. Strengths and limitations of 16S rRNA gene amplicon sequencing in revealing temporal microbial community dynamics. PLoS One. 2014;9(4):e93827.

28. Fierer N, Leff JW, Adams BJ, Nielsen UN, Bates ST, Lauber CL, et al. Cross-biome metagenomic analyses of soil microbial communities and their functional attributes. Proc Natl Acad Sci U S A. 2012;109(52):21390–5.

29. Sitarik AR, Havstad S, Levin AM, Lynch SV, Fujimura KE, Ownby DR, et al. Dog introduction alters the home dust microbiota. Indoor Air. 2018;28(4):539–47.

30. Alavanja MC, Sandler DP, McMaster SB, Zahm SH, McDonnell CJ, Lynch CF, et al. The Agricultural Health Study. Environ Health Perspect. 1996;104(4):362–9.

31. Carnes MU, Hoppin JA, Metwali N, Wyss AB, Hankinson JL, O’Connell EL, et al. House Dust Endotoxin Levels Are Associated with Adult Asthma in a U.S. Farming Population. Ann Am Thorac Soc. 2017;14(3):324–31.

32. Arbes SJ, Jr., Cohn RD, Yin M, Muilenberg ML, Burge HA, Friedman W, et al. House dust mite allergen in US beds: results from the First National Survey of Lead and Allergens in Housing. J Allergy Clin Immunol. 2003;111(2):408–14.

33. Sanders JG, Nurk S, Salido RA, Minich J, Xu ZZ, Zhu Q, et al. Optimizing sequencing protocols for leaderboard metagenomics by combining long and short reads. Genome Biol. 2019;20(1):226.

34. Babraham Institute. FastQC. 2010.

35. Langmead B, Salzberg SL. Fast gapped-read alignment with Bowtie 2. Nat Methods. 2012;9(4):357–9.

36. Beghini F, McIver LJ, Blanco-Míguez A, Dubois L, Asnicar F, Maharjan S, et al. Integrating taxonomic, functional, and strain-level profiling of diverse microbial communities with bioBakery 3. Elife. 2021;10.

37. Wood DE, Lu J, Langmead B. Improved metagenomic analysis with Kraken 2. Genome Biol. 2019;20(1):257.

38. Lu J, Breitwieser FP, Thielen P, Salzberg SL. Bracken: estimating species abundance in metagenomics data. PeerJ Computer Science. 2017;3:e104.

39. Davis NM, Proctor DM, Holmes SP, Relman DA, Callahan BJ. Simple statistical identification and removal of contaminant sequences in marker-gene and metagenomics data. Microbiome. 2018;6(1):226.

40. R Core Team. R: a language and environment for statistical computing. Vienna, Austria: R Foundation for Statistical Computing; 2020.

41. McMurdie PJ, Holmes S. phyloseq: an R package for reproducible interactive analysis and graphics of microbiome census data. PLoS One. 2013;8(4):e61217.

42. Anderson MJ. Permutational multivariate analysis of variance (PERMANOVA). Wiley statsref: statistics reference online. 2014:1–15.

43. Oksanen J, Simpson GL, Blanche FG, Kindt R, Legendre P, Minchin PR, et al. Community ecology package. R package version. 2013;2(0):321–6.

44. Wickham H. ggplot2: Elegant Graphics for Data Analysis: Springer; 2016.

45. Lin H, Peddada SD. Analysis of compositions of microbiomes with bias correction. Nat Commun. 2020;11(1):3514.

46. Benjamini Y, Hochberg Y. Controlling the false discovery rate: a practical and powerful approach to multiple testing. 1995. p. 289–300.

47. Lee MK, Carnes MU, Butz N, Azcarate-Peril MA, Richards M, Umbach DM, et al. Exposures Related to House Dust Microbiota in a U.S. Farming Population. Environ Health Perspect. 2018;126(6):067001.

48. Rausch P, Rühlemann M, Hermes BM, Doms S, Dagan T, Dierking K, et al. Comparative analysis of amplicon and metagenomic sequencing methods reveals key features in the evolution of animal metaorganisms. Microbiome. 2019;7(1).

49. Berg G, Rybakova D, Fischer D, Cernava T, Verges MC, Charles T, et al. Microbiome definition re-visited: old concepts and new challenges. Microbiome. 2020;8(1):103.

50. Lynch SV, Wood RA, Boushey H, Bacharier LB, Bloomberg GR, Kattan M, et al. Effects of early-life exposure to allergens and bacteria on recurrent wheeze and atopy in urban children. Journal of Allergy and Clinical Immunology. 2014;134(3):593-601.e12.

51. Abrahamsson TR, Jakobsson HE, Andersson AF, Björkstén B, Engstrand L, Jenmalm MC. Low diversity of the gut microbiota in infants with atopic eczema. Journal of Allergy and Clinical Immunology. 2012;129(2):434-40.e2.

52. Ley RE, Turnbaugh PJ, Klein S, Gordon JI. Human gut microbes associated with obesity. Nature. 2006;444(7122):1022–3.

53. Fu X, Ou Z, Zhang M, Meng Y, Li Y, Wen J, et al. Indoor bacterial, fungal and viral species and functional genes in urban and rural schools in Shanxi Province, China–association with asthma, rhinitis and rhinoconjunctivitis in high school students. Microbiome. 2021;9(1).

54. Weinstock DM, Brown AE. Rhodococcus equi: An Emerging Pathogen. Clinical Infectious Diseases. 2002;34(10):1379–85.

55. Wong KY, Chan, Y.C. and Wong, C.Y.,. Corynebacterium striatum as an emerging pathogen. Corynebacterium striatum as an emerging pathogen. 2010;76(4):371–2.

56. De Bentzmann SaP P. The Pseudomonas aeruginosa opportunistic pathogen and human infections. Environmental microbiology. 2011;13(7):1655–65.

57. Valkonen M, Täubel M, Pekkanen J, Tischer C, Rintala H, Zock J-P, et al. Microbial characteristics in homes of asthmatic and non-asthmatic adults in the ECRHS cohort. Indoor Air. 2018;28(1):16–27.

58. Pujar NK, Premakshi HG, Ganeshkar MP, Kamanavalli CM. Biodegradation of Pesticides Used in Agriculture by Soil Microorganisms. Enzymes for Pollutant Degradation: Springer Nature Singapore; 2022. p. 213–35.

59. Fiocchi A, Burks W, Bahna SL, Bielory L, Boyle RJ, Cocco R, et al. Clinical Use of Probiotics in Pediatric Allergy (cuppa): A World Allergy Organization Position Paper. World Allergy Organization Journal. 2012;5(11):148–67.

60. Kau AL, Ahern PP, Griffin NW, Goodman AL, Gordon JI. Human nutrition, the gut microbiome and the immune system. Nature. 2011;474(7351):327–36.

61. Skillman LC, Evans PN, Strompl C, Joblin KN. 16S rDNA directed PCR primers and detection of methanogens in the bovine rumen. Letters in Applied Microbiology. 2006;42(3):222–8.

62. Hatti-Kaul R, Chen L, Dishisha T, Enshasy HE. Lactic acid bacteria: from starter cultures to producers of chemicals. FEMS Microbiology Letters. 2018;365(20).

63. Kraemer JG, Aebi S, Hilty M, Oppliger A. Nasal microbiota composition dynamics after occupational change in animal farmers suggest major shifts. Sci Total Environ. 2021;782:146842.

64. Shukla SK, Ye Z, Sandberg S, Reyes I, Fritsche TR, Keifer M. The nasal microbiota of dairy farmers is more complex than oral microbiota, reflects occupational exposure, and provides competition for staphylococci. PLOS ONE. 2017;12(8):e0183898.

65. Bull CT, De Boer SH, Denny TP, Firrao G, Fischer-Le Saux M, Saddler GS, et al. List of New Names of Plant Pathogenic Bacteria. Journal of Plant Pathology. 2012;94(1):21–7.

66. Goswami RS, Kistler HC. Heading for disaster: Fusarium graminearum on cereal crops. Molecular Plant Pathology. 2004;5(6):515–25.

67. Staats JJ, Feder I, Okwumabua O, Chengappa MM. Streptococcus suis: Past and Present. Veterinary Research Communications. 1997;21(6):381–407.

68. Bannoehr J, Guardabassi L. Staphylococcus pseudintermedius in the dog: taxonomy, diagnostics, ecology, epidemiology and pathogenicity. Veterinary Dermatology. 2012;23(4):253–e52.

69. Sepich-Poore GD, Zitvogel L, Straussman R, Hasty J, Wargo JA, Knight R. The microbiome and human cancer. Science. 2021;371(6536).

70. Dunn RR, Fierer N, Henley JB, Leff JW, Menninger HL. Home Life: Factors Structuring the Bacterial Diversity Found within and between Homes. PLoS ONE. 2013;8(5):e64133.

