## Supplemental methods for "Metagenomics reveals novel microbial signatures of farm exposures in house dust"

***SM1. Study population selection and inclusion criteria***

The Agricultural Health Study (AHS) is a prospective cohort of licensed pesticide applicators that comprises mainly farmers (n = 52,394) and their spouses (n = 32,345) enrolled between 1993 and 1997 in North Carolina and Iowa (1). We selected participants from the Agricultural Lung Health Study (ALHS), a nested asthma case-control study within the parent AHS. We identified potential ALHS participants from 44,130 AHS participants who responded to a follow-up questionnaire, could provide informed consent, and still lived in either North Carolina or Iowa.

Among the 44,130 AHS participants, we defined asthma cases using three selection criteria to avoid missing undiagnosed asthma and misclassification due to chronic obstructive pulmonary disorder (COPD):

1. Reported previous diagnosis of asthma and currently having asthma, in addition to reporting NOT having COPD or emphysema (n=876)

2. Among self-reported non-smokers or past smokers, reported current asthma symptoms (e.g., wheezing or awakening due to respiratory symptoms) and the use of asthma medication, with no diagnosis of COPD or emphysema to capture potential undiagnosed asthma (n=309)

3. Among reported non-smokers or past smokers, reported current asthma and previous diagnosis of either COPD or emphysema to capture respiratory co-morbidities (n=38)

Among the 2,363 putative asthma cases identified in AHS based on these criteria, 1,223 enrolled in ALHS, representing a 51.7% response rate to the follow-up questionnaire. We randomly selected controls from participants who reported not currently having asthma, experiencing asthma symptoms, or using asthma medications or inhalers in the past 12 months. To achieve a suitably sized comparison group, we included 2,078 controls in ALHS, representing a response rate of 50.0% to the follow-up survey.

***SM2. Bioinformatics and Quality Control***

At each sequencing stage, FastQC v0.11.5 (2) was used to assess the quality of reads by Phred quality score, GC content, the presence of adapters, overrepresented k-mers, duplicated reads rate, and PCR artifacts or contaminations. Low-quality and adaptor sequences were first removed using Atropos (3). Bases with a Phred score <15 and throw-away reads shorter than 100 bp (*-q 15 --minimum-length 100*) were trimmed, starting from the end of the read. The internal sequencing standard PhiX 174 and human sequences were subsequently identified using bowtie2 (4), SAMtools (5), and BEDtools (6) to remove aligned sequences and their mates. The above procedures were performed in the Qiita pipeline (7) by the IGM Genomics Center at the University of California San Diego.

All raw reads of both ends (3’ or 5’) passed the basic FastQC figures (per base sequence quality and per sequence quality scores), and no low-quality sequences remained. Bimodal shape was observed in per sequence GC content, indicating the wide distribution of genome GC content across multiple species in metagenomic samples.

As an additional quality control step, we performed in silico separation of bacterial reads from contaminant reads using KneadData v0.7.10 (https://huttenhower.sph.harvard.edu/kneaddata/) with default settings *(--bypass-trim –serial --bowtie2-options=” --very-sensitive”*). In addition to the human and PhiX genomes, we included farm animals (cows (ARS-UCD1.2), pigs (Sscrofa11.1), chickens (GRCg6a), turkeys (Turkey_5.1), horses (EquCab3.0), goats (ARS1) and sheep (Oar_rambouillet_v1.0)), pets (dogs (CanFam3.1) and cats (Felis_catus_9.0)), and dust mites (Dfa_Genome_UMICH_USM_1.1) as these are potential  contamination sources. We downloaded the corresponding animal reference genomes from the NCBI database and built them for the bowtie2 index (Supplementary Table 1).

We then classified the resulting paired-end reads using Kraken2 v2.1.1 (8) with pre-compiled data comprising RefSeq genomes for bacteria, archaea, eukaryotes, fungi, viruses, and plasmids and NCBI taxonomy information, with a confidence score threshold of 0.05 *(--confidence 0.05*), to enhance the accuracy of taxonomic assignments (9). We then ran the Kraken2 output against Bracken v2.5.0 (10) with default parameters (*-r 100 -l S -t 10*) to quantify abundance at the species level. We built the Bracken database with the default 35-mers length. Supplementary Tables 2 and 3 summarize the overall statistics of read sequences and the proportion of each host genome contaminant across samples.

Due to low biomass in the dust samples, we separately performed and processed two sequencing runs. Metagenomics datasets from low biomass samples are particularly vulnerable to microbial contamination from the sample collection instrument, sequencing kit, and laboratory reagents. We incorporated ‘blank’ controls by sequencing sterile water without adding dust sample DNA extractions (11). This allowed us to use the decontam R package v1.10.0 (12) to identify contaminated DNA sequences not truly present in the sampled community for each run. We then removed contaminants identified in either run. Using this process, we filtered out 168 taxa (Supplementary Table 4). After separately conducting preprocessing and filtering for each run, we generated pooled abundance data by summing the abundance data from both runs.

***SM3. Statistical analysis***

We described the statistical analysis with the following formula. We first fit a baseline univariate regression model for each exposure (xk) to identify those that were significant:

alpha diversity ~ xk + asthma, k=1,…,17.

We then fit regression models by adding individual interactions between the significant exposures (x_sig_i, i=1.,,,.p) and state of residence while retaining the main effects of significant exposures in the model, with the aim of identifying exposure effects that differ by state of residence:

alpha diversity ~ x_all_sig + state + x_sig_i*state + asthma, i=1,…,p.

**References**

1.	Alavanja MC, Sandler DP, McMaster SB, Zahm SH, McDonnell CJ, Lynch CF, et al. The Agricultural Health Study. Environ Health Perspect. 1996;104(4):362-9.

2.	Babraham Institute. FastQC. 2010.

3.	Didion JP, Martin M, Collins FS. Atropos: specific, sensitive, and speedy trimming of sequencing reads. PeerJ. 2017;5:e3720.

4.	Langmead B, Salzberg SL. Fast gapped-read alignment with Bowtie 2. Nat Methods. 2012;9(4):357-9.

5.	Li H, Handsaker B, Wysoker A, Fennell T, Ruan J, Homer N, et al. The Sequence Alignment/Map format and SAMtools. Bioinformatics. 2009;25(16):2078-9.

6.	Quinlan AR, Hall IM. BEDTools: a flexible suite of utilities for comparing genomic features. Bioinformatics. 2010;26(6):841-2.

7.	Gonzalez A, Navas-Molina JA, Kosciolek T, McDonald D, Vázquez-Baeza Y, Ackermann G, et al. Qiita: rapid, web-enabled microbiome meta-analysis. Nat Methods. 2018;15(10):796-8.

8.	Wood DE, Lu J, Langmead B. Improved metagenomic analysis with Kraken 2. Genome Biol. 2019;20(1):257.

9.	Ye SH, Siddle KJ, Park DJ, Sabeti PC. Benchmarking Metagenomics Tools for Taxonomic Classification. Cell. 2019;178(4):779-94.

10.	Lu J, Breitwieser FP, Thielen P, Salzberg SL. Bracken: estimating species abundance in metagenomics data. PeerJ Computer Science. 2017;3:e104.

11.	Quince C, Walker AW, Simpson JT, Loman NJ, Segata N. Shotgun metagenomics, from sampling to analysis. Nat Biotechnol. 2017;35(9):833-44.

12.	Davis NM, Proctor DM, Holmes SP, Relman DA, Callahan BJ. Simple statistical identification and removal of contaminant sequences in marker-gene and metagenomics data. Microbiome. 2018;6(1):226.
