## Supplemental contents for "Metagenomics reveals novel microbial signatures of farm exposures in house dust"

**Supplemental Materials Table of Contents**

**Supplemental Methods**

SM1. Study population selection and inclusion criteria

SM2. Bioinformatics and quality control

SM3. Statistical analysis

**Supplemental tables**

Table S1. Reference genome version

Table S2. Quality control summary statistics

Table S3. Proportion of host genome contaminants across samples

Table S4. Contraoriented DNA filtered by decontam R package (168 taxa)

Table S5. Relative abundance for phylum

Table S6. Relative abundance for genus

Table S7. Full model for alpha diversity association analysis

Table S8. Summary of differentially abundant taxa for ANCOM-BC analysis (780 taxa)

Table S9. The number of differentially abundant taxa to individual exposure (372 taxa)

Table S10. Interaction effect for alpha diversity association analysis

Table S11. Interaction effect for PERMANOVA using unweighted UniFrac distances

Table S12. Interaction effect for PERMANOVA using weighted UniFrac distances

Table S13. Stratified model for alpha diversity association analysis

Table S14. Stratified model for beta diversity association analysis

**Supplemental figures**

Figure S1. Quality control workflow

Figure S2. Sample and rare taxa filtering

Figure S3. Non-metric multidimensional scaling (NMDS) analysis based on unweighted UniFrac distances for all exposures

Figure S4. Non-metric multidimensional scaling (NMDS) analysis based on weighted UniFrac distances for all exposures

Figure S5. Volcano plot of differentially abundant taxa

Figure S6. Volcano plot of differentially abundant taxa stratified in North Carolina

Figure S7. Volcano plot of differentially abundant taxa stratified in Iowa
