## Supplemental figures 1-4 for "Metagenomics reveals novel microbial signatures of farm exposures in house dust"

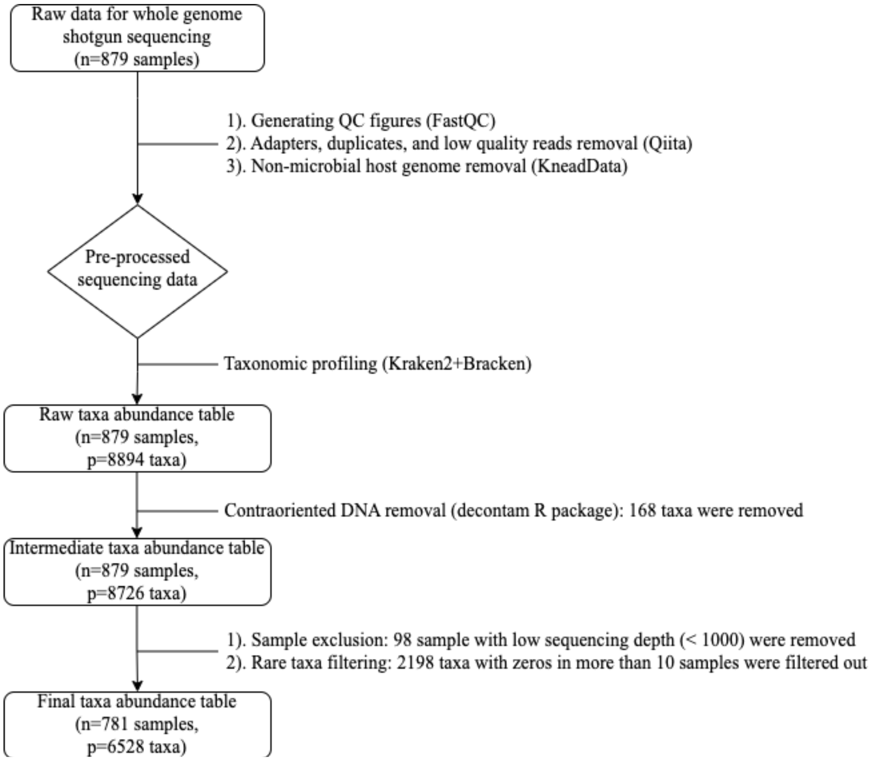


Supplementary Figure 1. Workflow of the Quality Control for WGS.

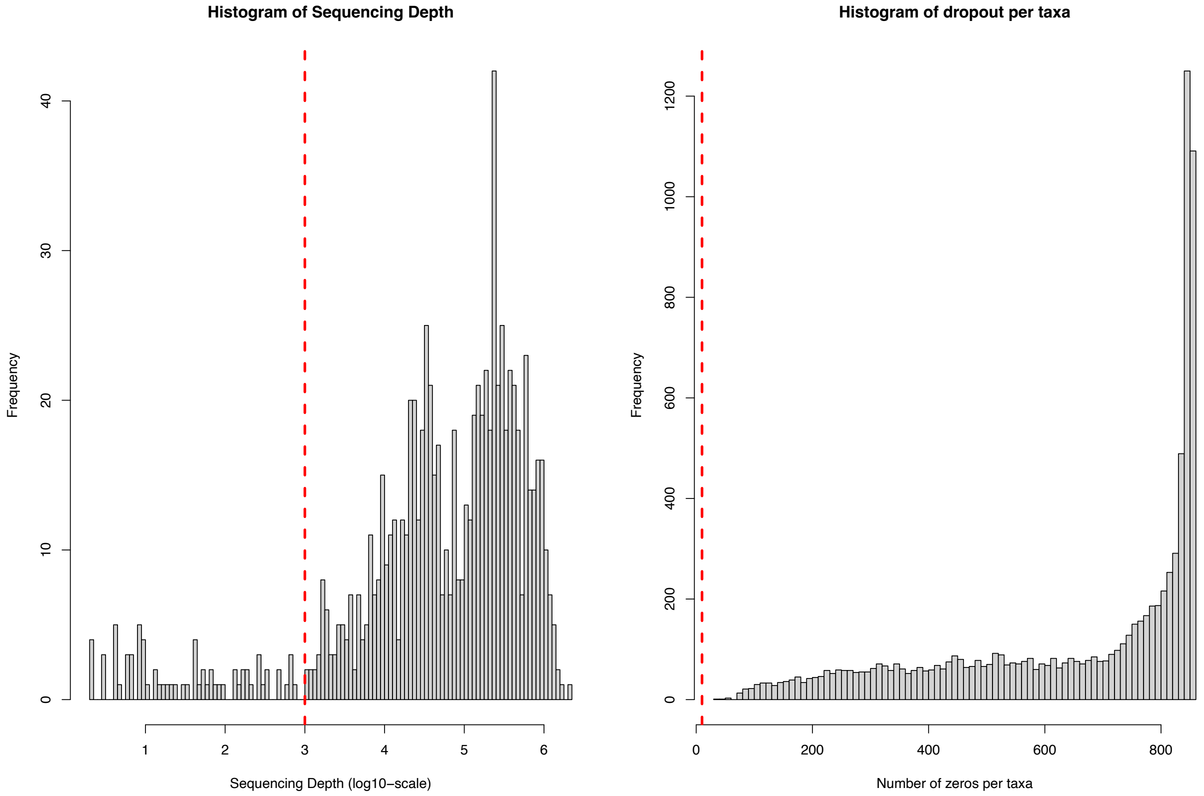


Supplementary Figure 2. Sample and rare taxa filtering criteria for WGS. **(a).** Histogram of sequencing depth. **(b).** Histogram of number of zeros per taxon. Red dotted lines indicate the threshold.

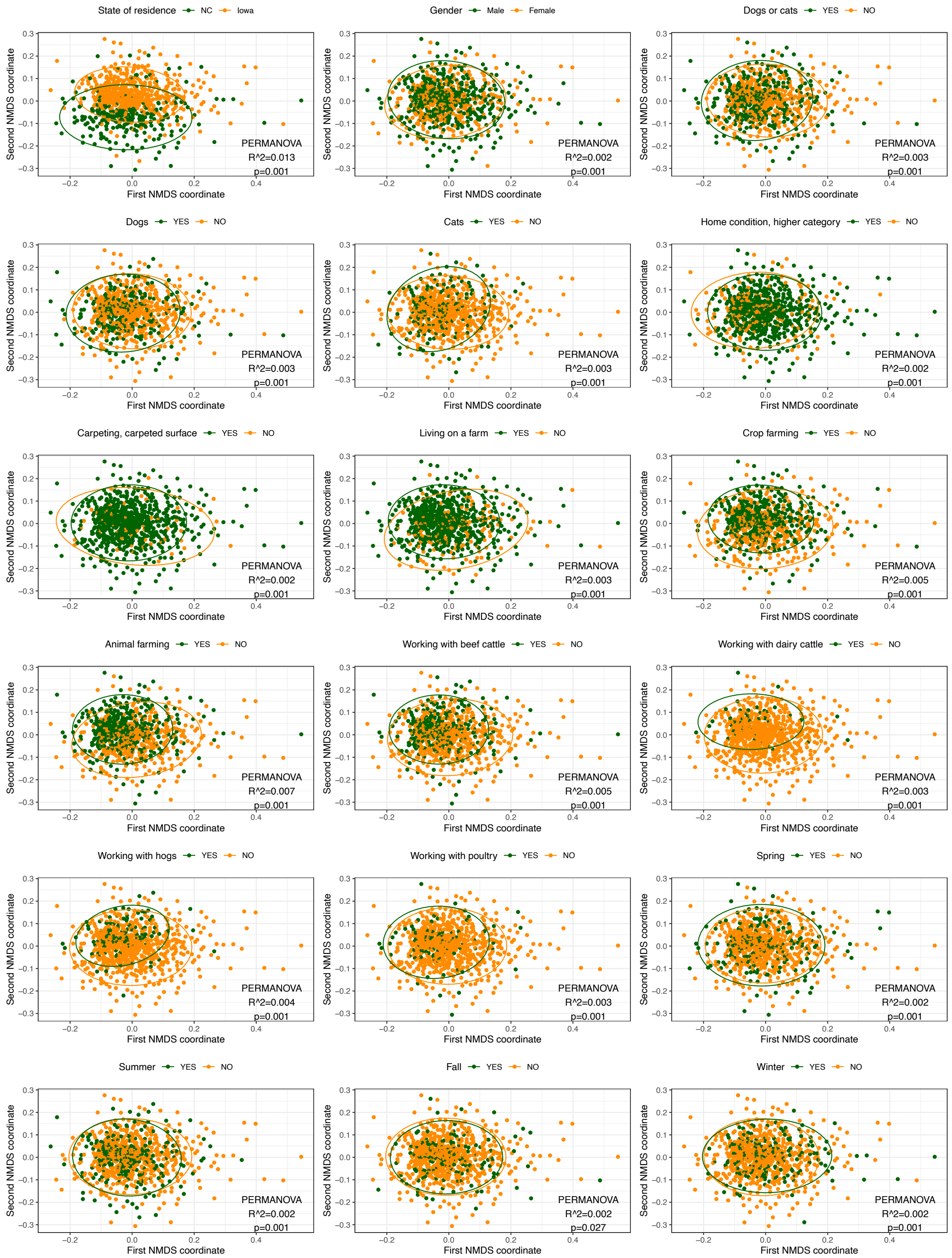


Supplementary Figure 3. Non-metric multidimensional scaling (NMDS) analysis based on unweighted UniFrac distances for all exposures. The dust microbial community of each sample is indicated with one dot. $R^2$ value (percentage of variance explained by exposure) and p-value from the PERMANOVA analysis are reported.

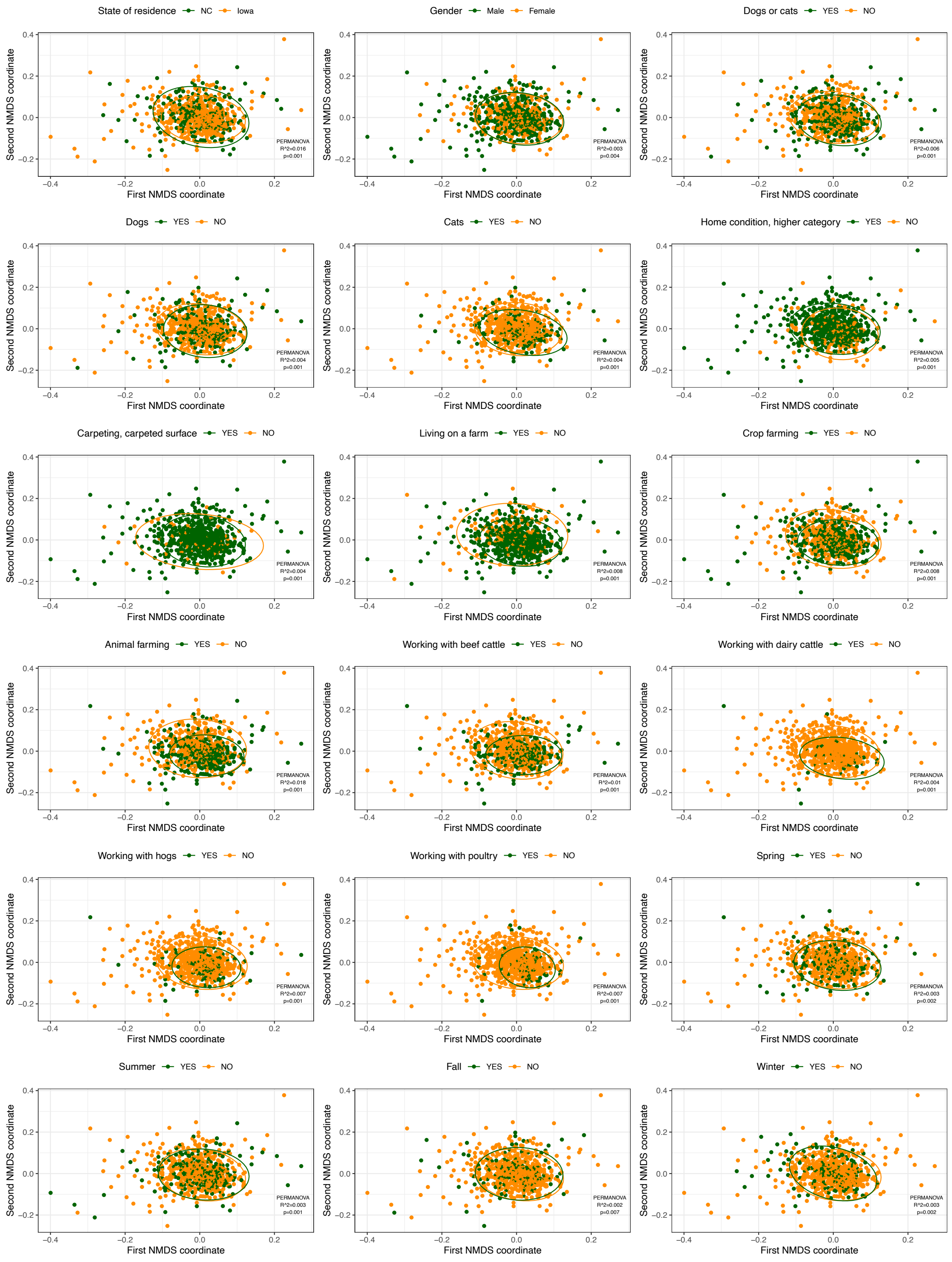


Supplementary Figure 4. Non-metric multidimensional scaling (NMDS) analysis based on weighted UniFrac distances for all exposures. The dust microbial community of each sample is indicated with one dot. $R^2$ value (percentage of variance explained by exposure) and p-value from the PERMANOVA analysis are reported.
