## Supplemental figure 5 for "Metagenomics reveals novel microbial signatures of farm exposures in house dust"

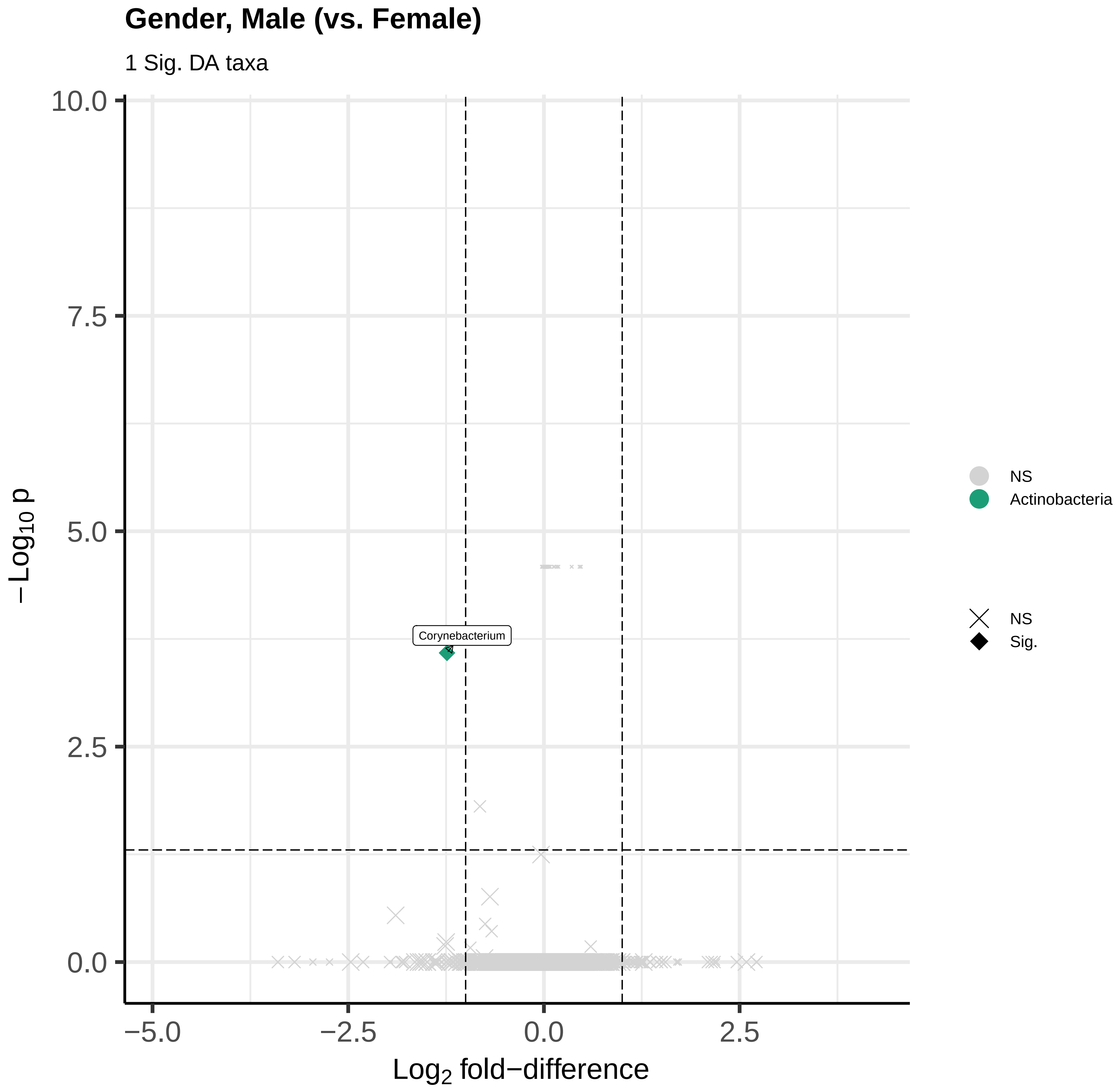

Gender, Male (vs. Female)
1 Sig. DA taxa
10.0
7.5
5.0
2.5
0.0
−Log10 p
Corynebacterium
−5.0
−2.5
0.0
2.5
Log2 fold−difference
NS
Actinobacteria
NS
Sig.

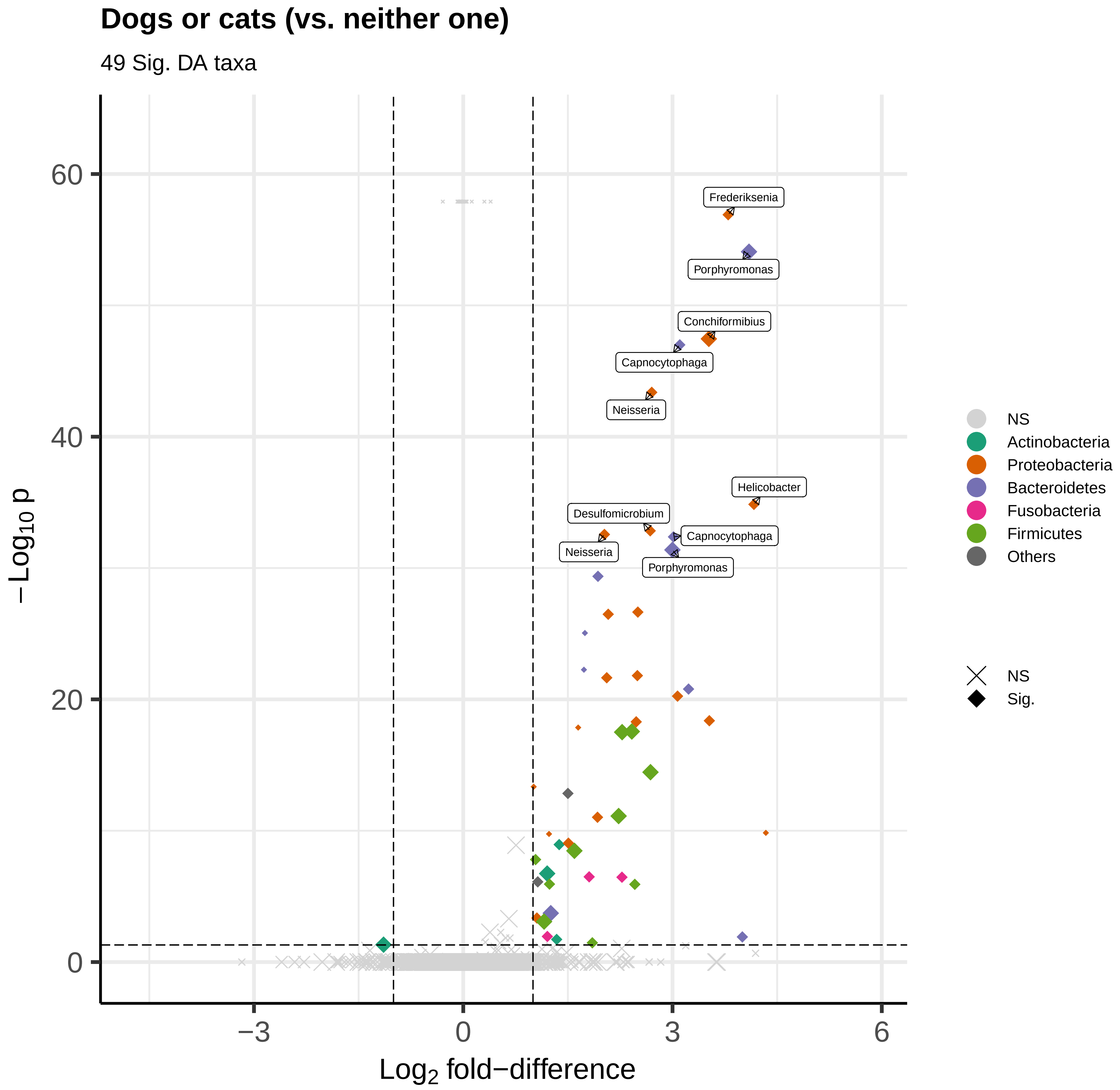

Dogs or cats (vs. neither one)
49 Sig. DA taxa
Frederiksenia
Porphyromonas
Conchiformibius
Capnocytophaga
Neisseria
Helicobacter
Desulfomicrobium
Capnocytophaga
Neisseria
Porphyromonas
60
40
20
0
−3
0
3
6
$-Log_{10}$ p
$Log_2$ fold−difference
NS
Actinobacteria
Proteobacteria
Bacteroidetes
Fusobacteria
Firmicutes
Others
NS
Sig.

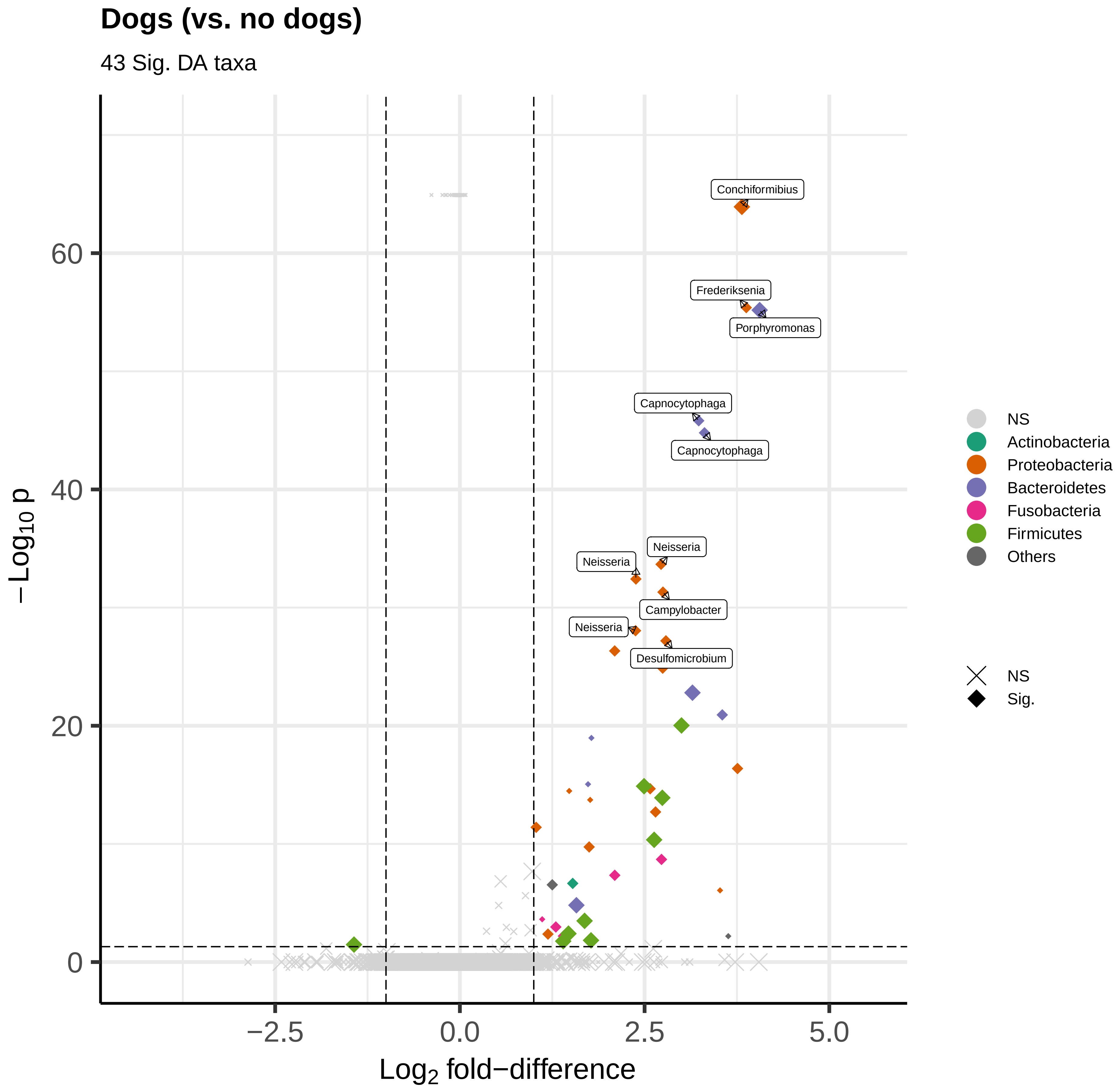

Dogs (vs. no dogs)
43 Sig. DA taxa
Conchiformibius
Frederiksenia
Porphyromonas
Capnocytophaga
Capnocytophaga
Neisseria
Neisseria
Campylobacter
Neisseria
Desulfomicrobium
60
40
20
0
−Log10 p
−2.5
0.0
2.5
5.0
Log2 fold−difference
NS
Actinobacteria
Proteobacteria
Bacteroidetes
Fusobacteria
Firmicutes
Others
NS
Sig.

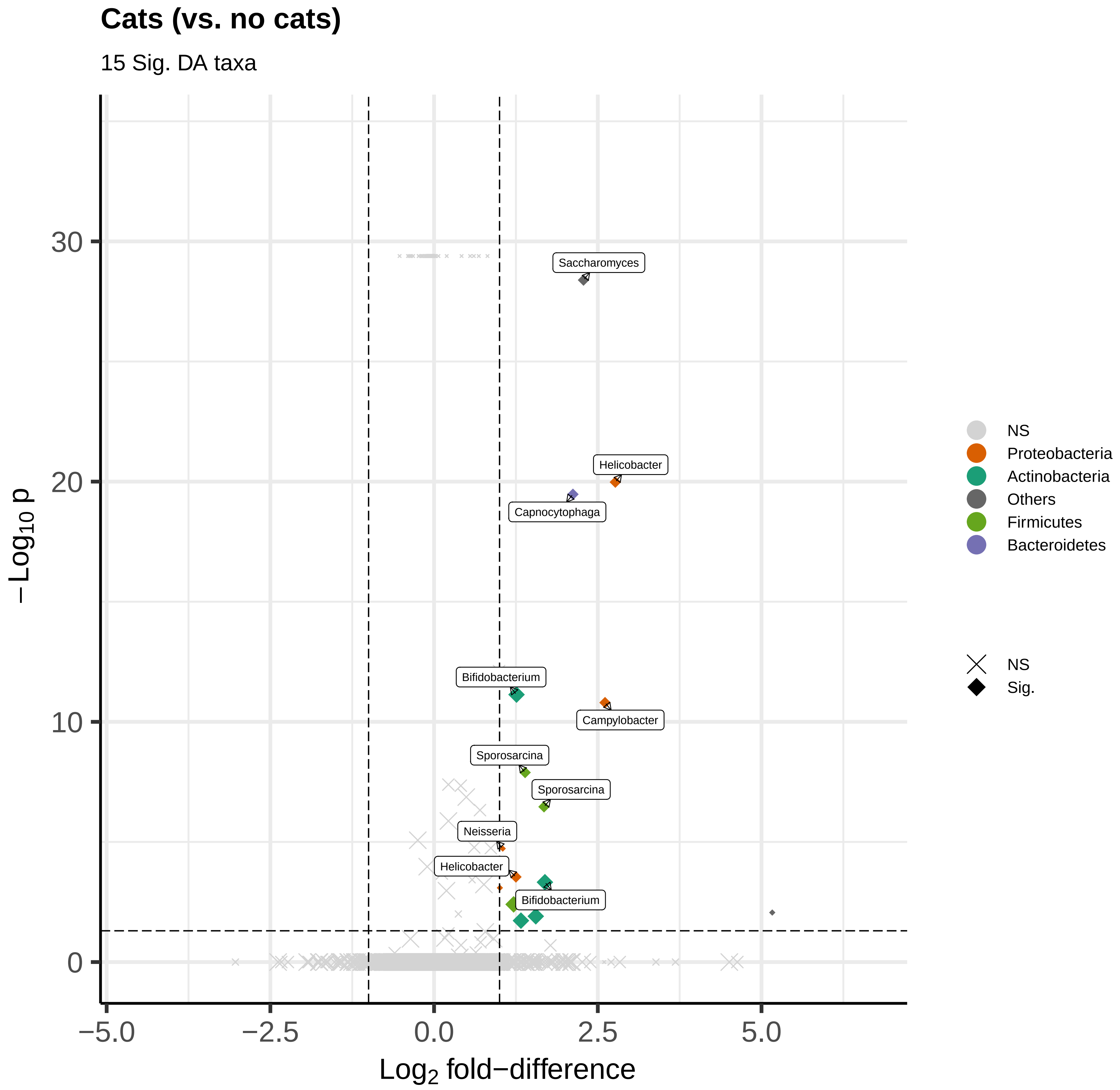
Cats (vs. no cats)
15 Sig. DA taxa
Saccharomyces
Helicobacter
Capnocytophaga
Bifidobacterium
Campylobacter
Sporosarcina
Sporosarcina
Neisseria
Helicobacter
Bifidobacterium
30
20
10
0
−5.0
−2.5
0.0
2.5
5.0
Log2 fold−difference
−Log10 p
NS
Proteobacteria
Actinobacteria
Others
Firmicutes
Bacteroidetes
NS
Sig.

### Home condition, higher category (vs. lower category)

8 Sig. DA taxa

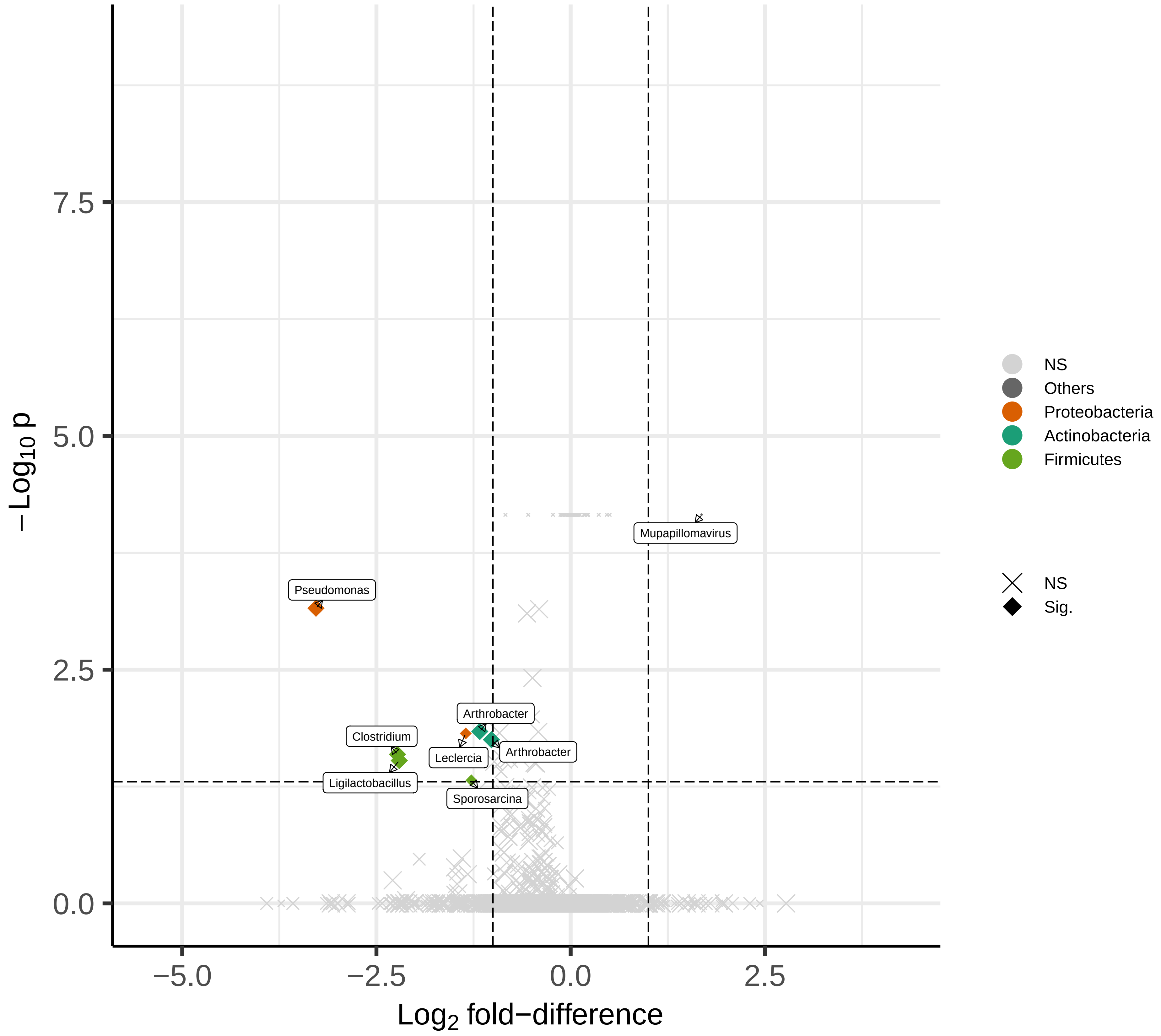

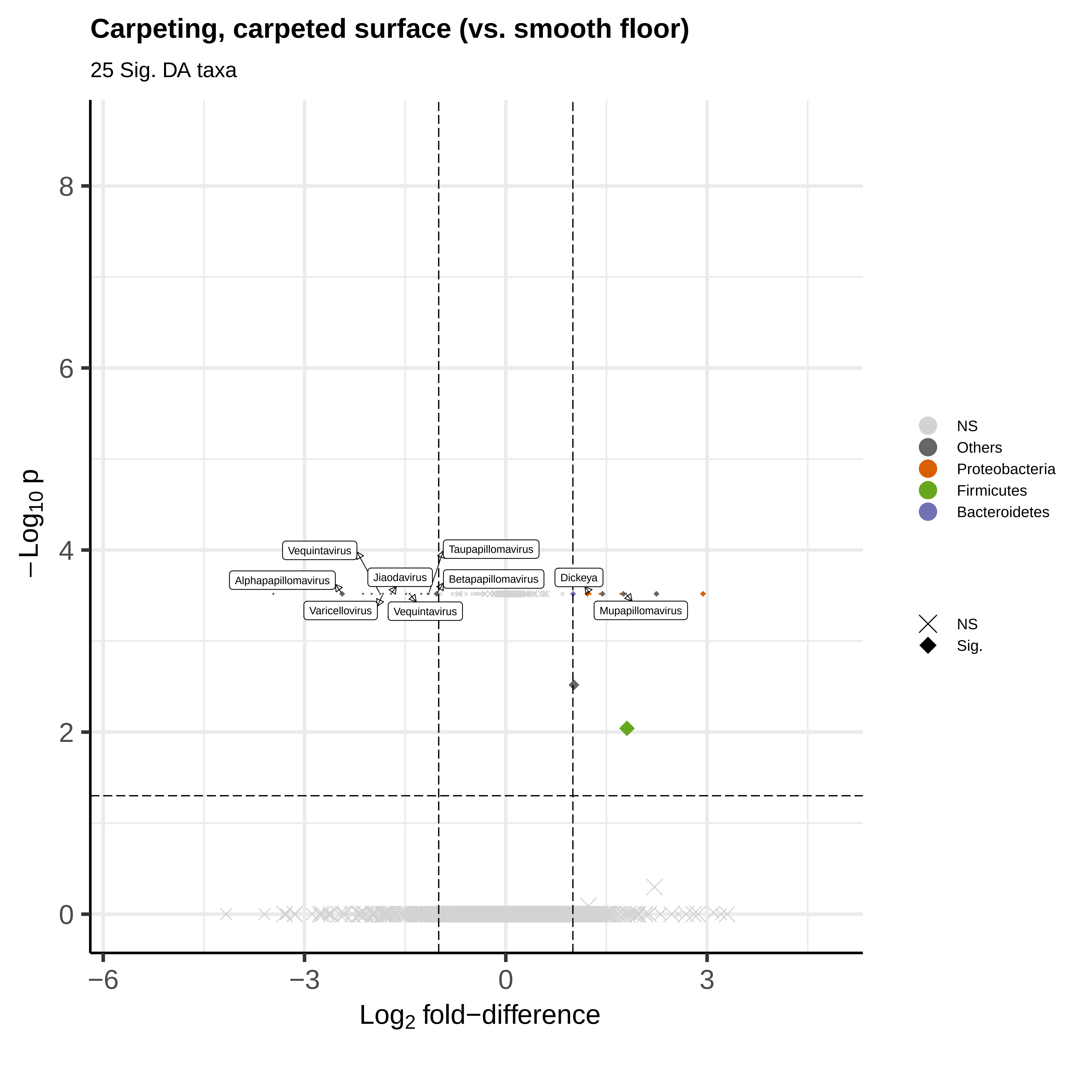

Carpeting, carpeted surface (vs. smooth floor)
25 Sig. DA taxa
−Log10 p
Log2 fold−difference
8
6
4
2
0
−6
−3
0
3
Vequintavirus
Taupapillomavirus
Alphapapillomavirus
Jiaodavirus
Betapapillomavirus
Dickeya
Varicellovirus
Vequintavirus
Mupapillomavirus
NS
Others
Proteobacteria
Firmicutes
Bacteroidetes
NS
Sig.

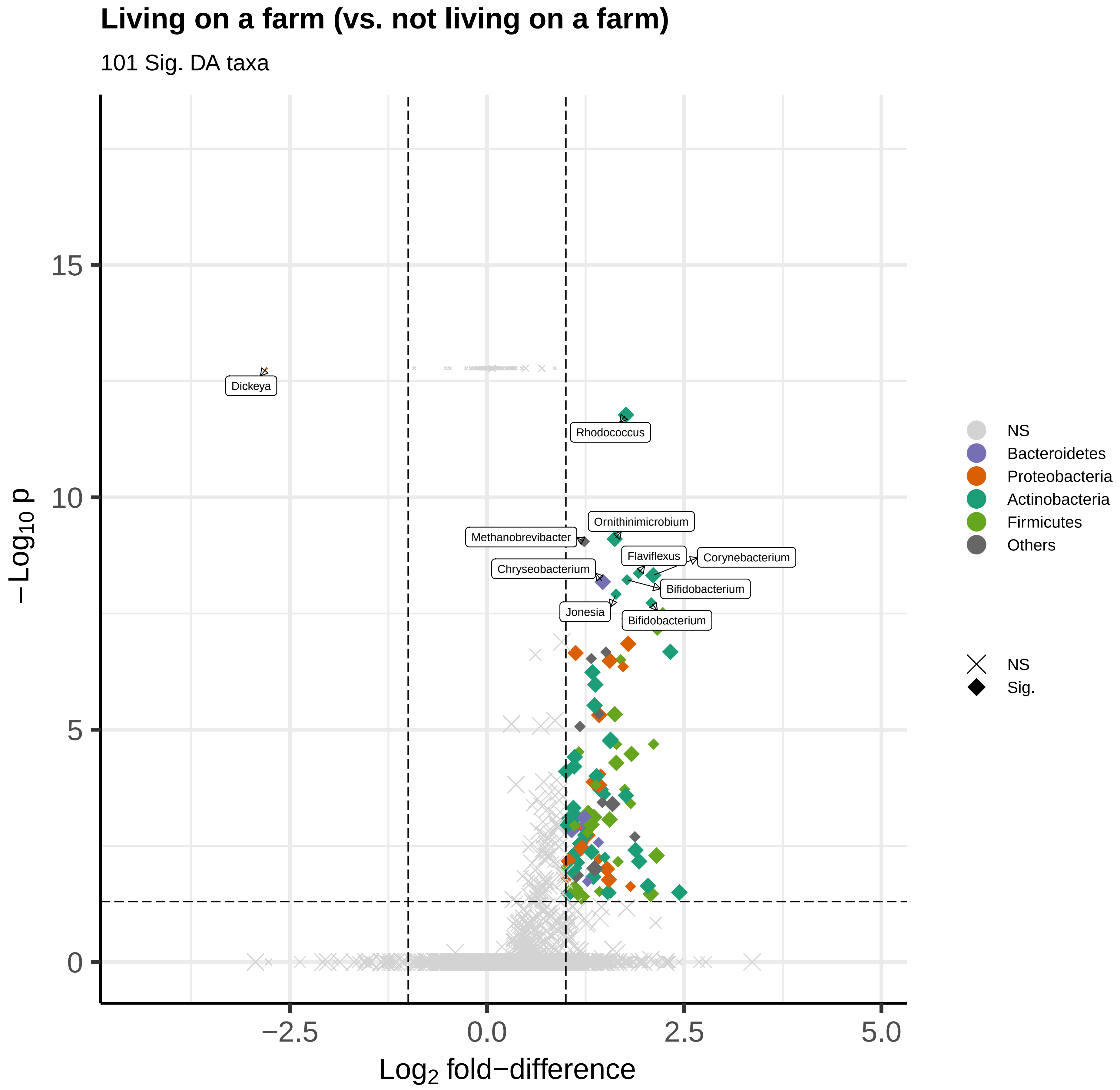
Living on a farm (vs. not living on a farm)
101 Sig. DA taxa
−Log10 p
Log2 fold−difference
Dickeya
Rhodococcus
Ornithinimicrobium
Methanobrevibacter
Flaviflexus
Corynebacterium
Chryseobacterium
Bifidobacterium
Jonesia
Bifidobacterium
NS
Bacteroidetes
Proteobacteria
Actinobacteria
Firmicutes
Others
NS
Sig.
15
10
5
0
−2.5
0.0
2.5
5.0

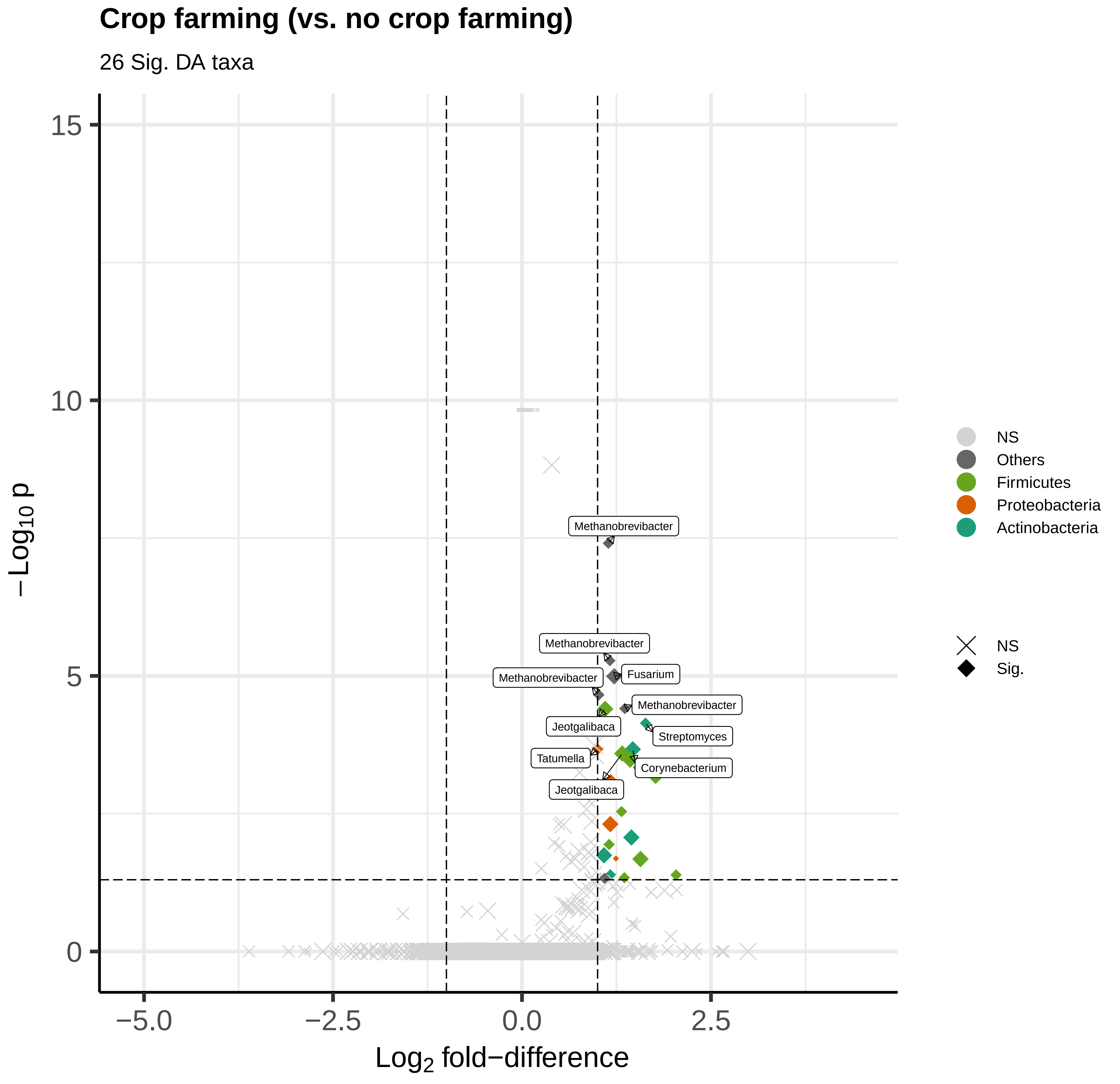

Crop farming (vs. no crop farming)
26 Sig. DA taxa
Methanobrevibacter
Methanobrevibacter
Fusarium
Methanobrevibacter
Methanobrevibacter
Jeotgalibaca
Streptomyces
Tatumella
Corynebacterium
Jeotgalibaca
15
10
5
0
−Log10 p
−5.0
−2.5
0.0
2.5
Log2 fold−difference
NS
Others
Firmicutes
Proteobacteria
Actinobacteria
NS
Sig.

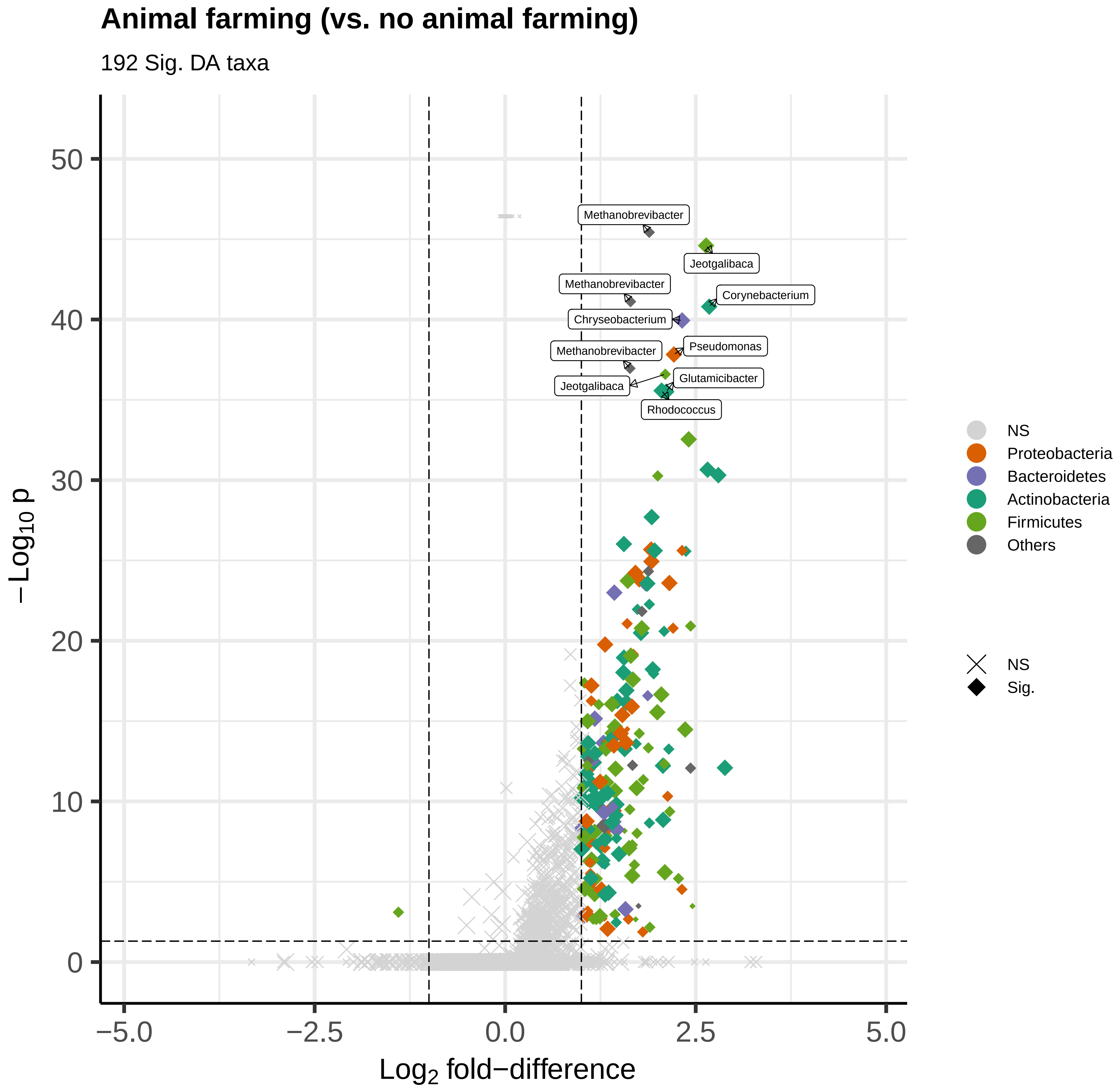

Animal farming (vs. no animal farming)
192 Sig. DA taxa
Methanobrevibacter
Jeotgalibaca
Methanobrevibacter
Corynebacterium
Chryseobacterium
Pseudomonas
Methanobrevibacter
Glutamicibacter
Jeotgalibaca
Rhodococcus
NS
Proteobacteria
Bacteroidetes
Actinobacteria
Firmicutes
Others
NS
Sig.
−Log10 p
Log2 fold−difference
0
10
20
30
40
50
−5.0
−2.5
0.0
2.5
5.0

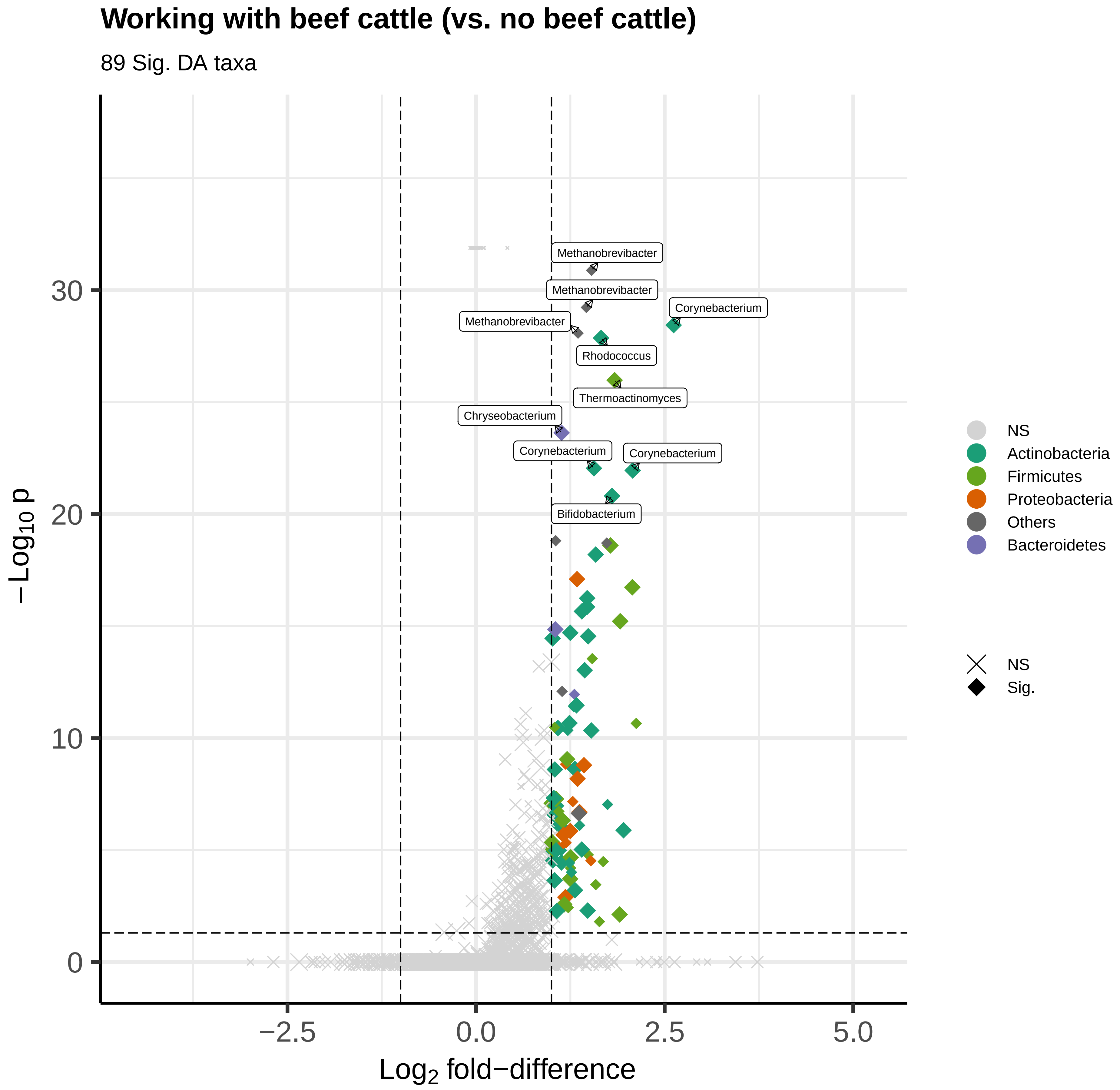

Working with beef cattle (vs. no beef cattle)
89 Sig. DA taxa
Methanobrevibacter
Methanobrevibacter
Corynebacterium
Methanobrevibacter
Rhodococcus
Thermoactinomyces
Chryseobacterium
Corynebacterium
Corynebacterium
Bifidobacterium
−Log10 p
Log2 fold−difference
NS
Actinobacteria
Firmicutes
Proteobacteria
Others
Bacteroidetes
NS
Sig.

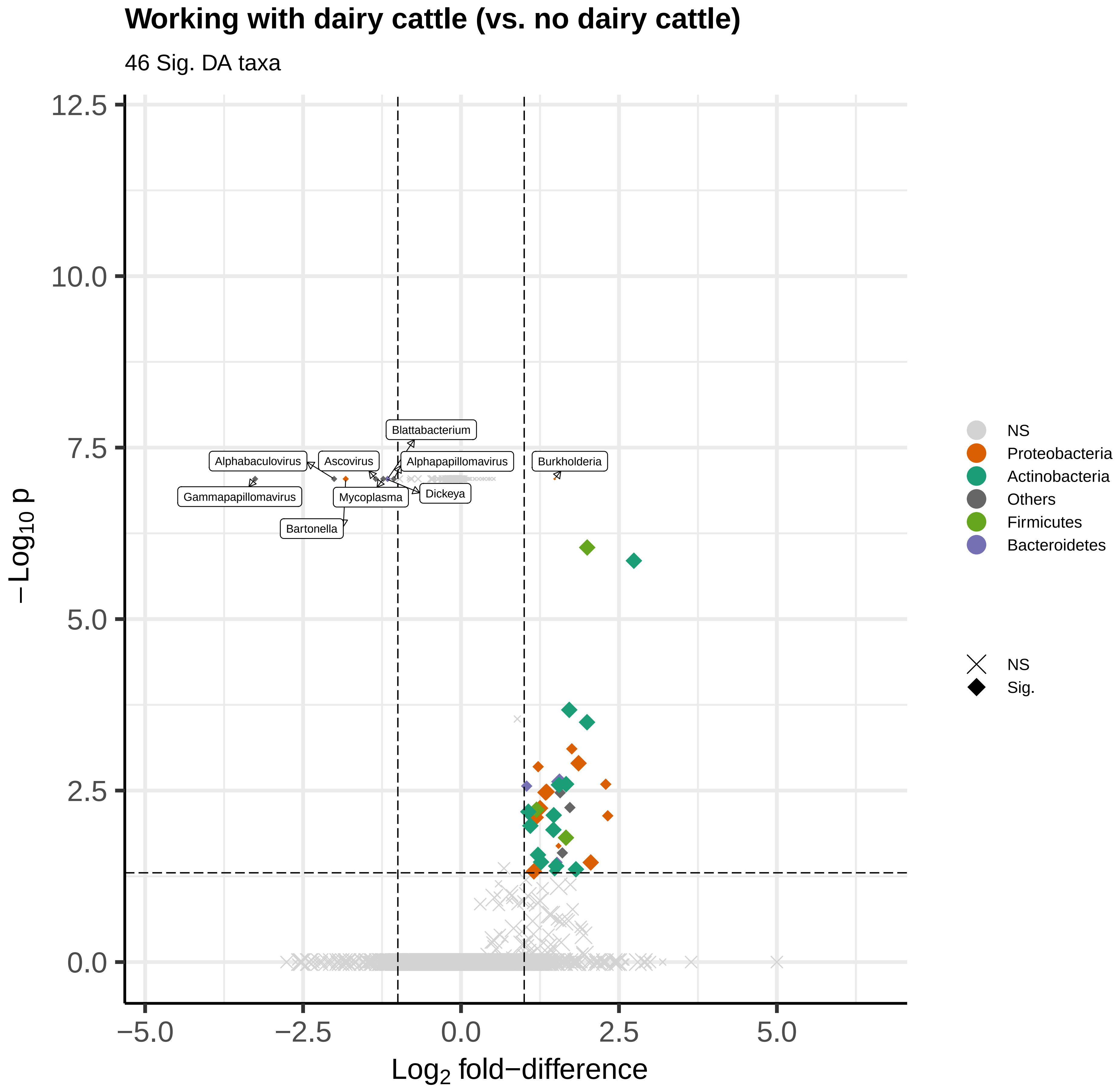

Working with dairy cattle (vs. no dairy cattle)
46 Sig. DA taxa
12.5
10.0
7.5
5.0
2.5
0.0
−5.0
−2.5
0.0
2.5
5.0
$-Log_{10}$ p
$Log_2$ fold−difference
Blattabacterium
Alphabaculovirus
Ascovirus
Alphapapillomavirus
Burkholderia
Gammapapillomavirus
Mycoplasma
Dickeya
Bartonella
NS
Proteobacteria
Actinobacteria
Others
Firmicutes
Bacteroidetes
NS
Sig.

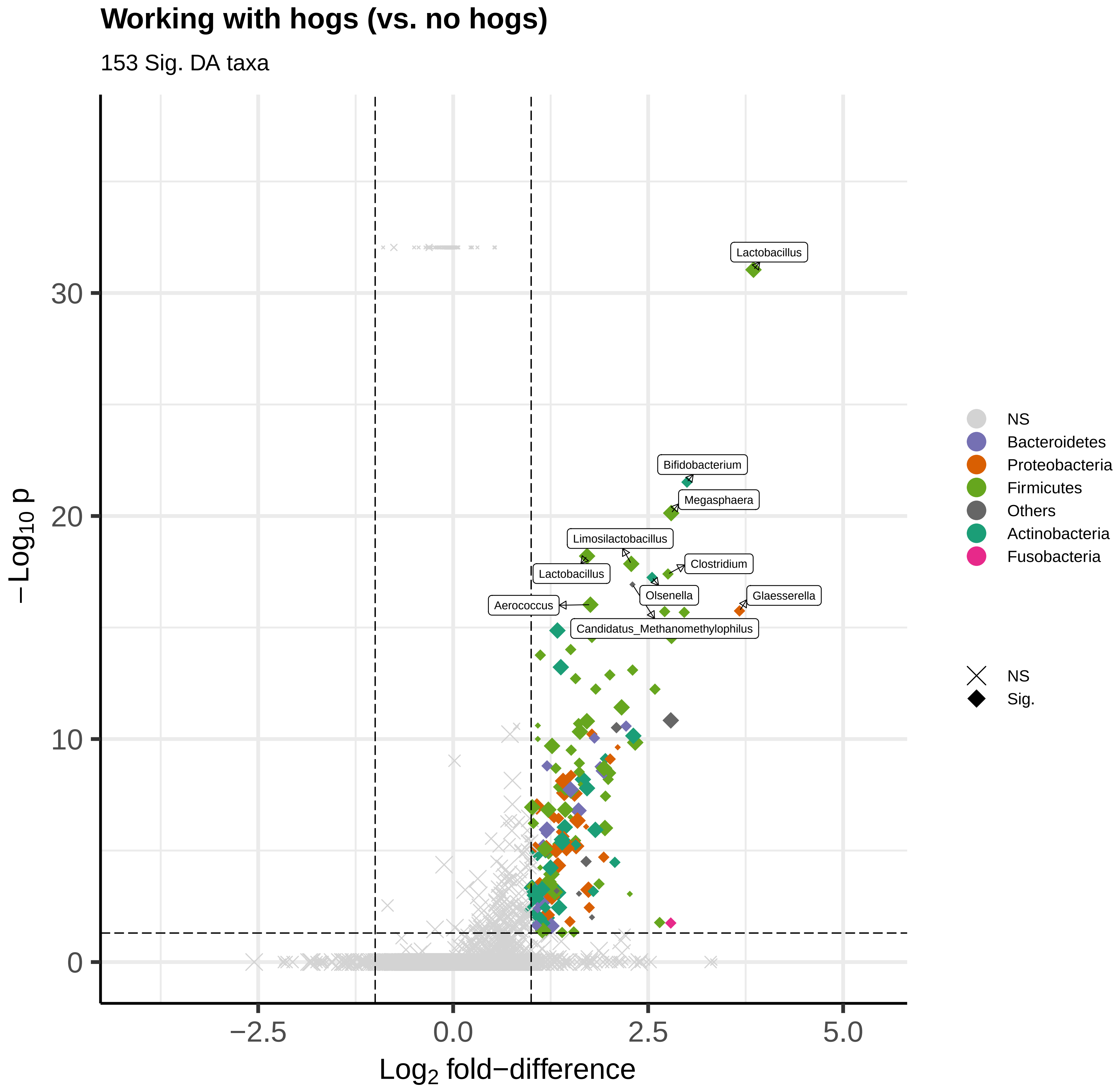
Working with hogs (vs. no hogs)
153 Sig. DA taxa
Lactobacillus
Bifidobacterium
Megasphaera
Limosilactobacillus
Clostridium
Lactobacillus
Olsenella
Glaesserella
Aerococcus
Candidatus_Methanomethylophilus
−Log10 p
Log2 fold−difference
30
20
10
0
−2.5
0.0
2.5
5.0
NS
Bacteroidetes
Proteobacteria
Firmicutes
Others
Actinobacteria
Fusobacteria
NS
Sig.

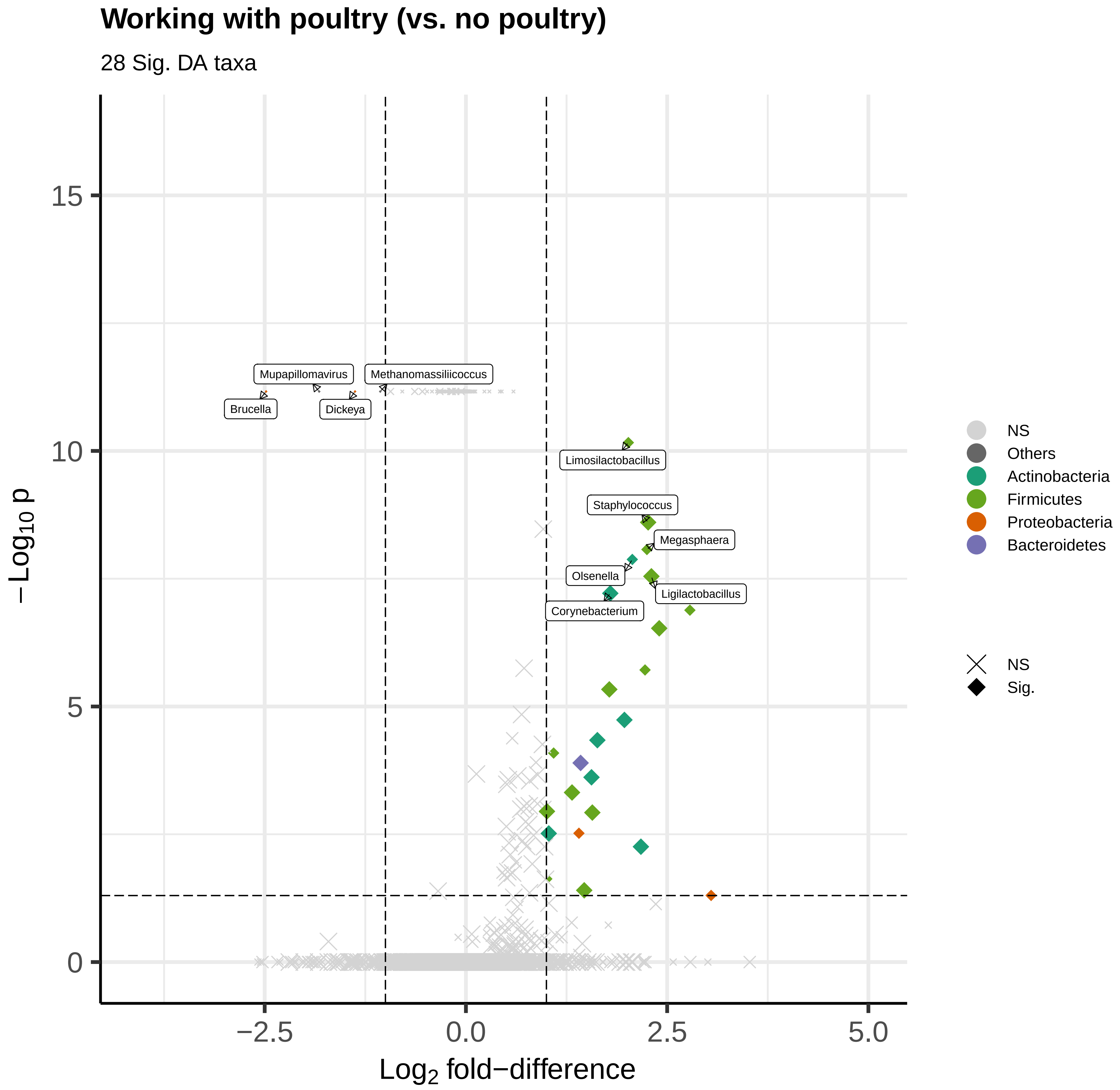

Working with poultry (vs. no poultry)
28 Sig. DA taxa
Mupapillomavirus
Methanomassiliicoccus
Brucella
Dickeya
Limosilactobacillus
Staphylococcus
Megasphaera
Olsenella
Ligilactobacillus
Corynebacterium
15
10
5
0
−2.5
0.0
2.5
5.0
Log2 fold−difference
−Log10 p
NS
Others
Actinobacteria
Firmicutes
Proteobacteria
Bacteroidetes
NS
Sig.

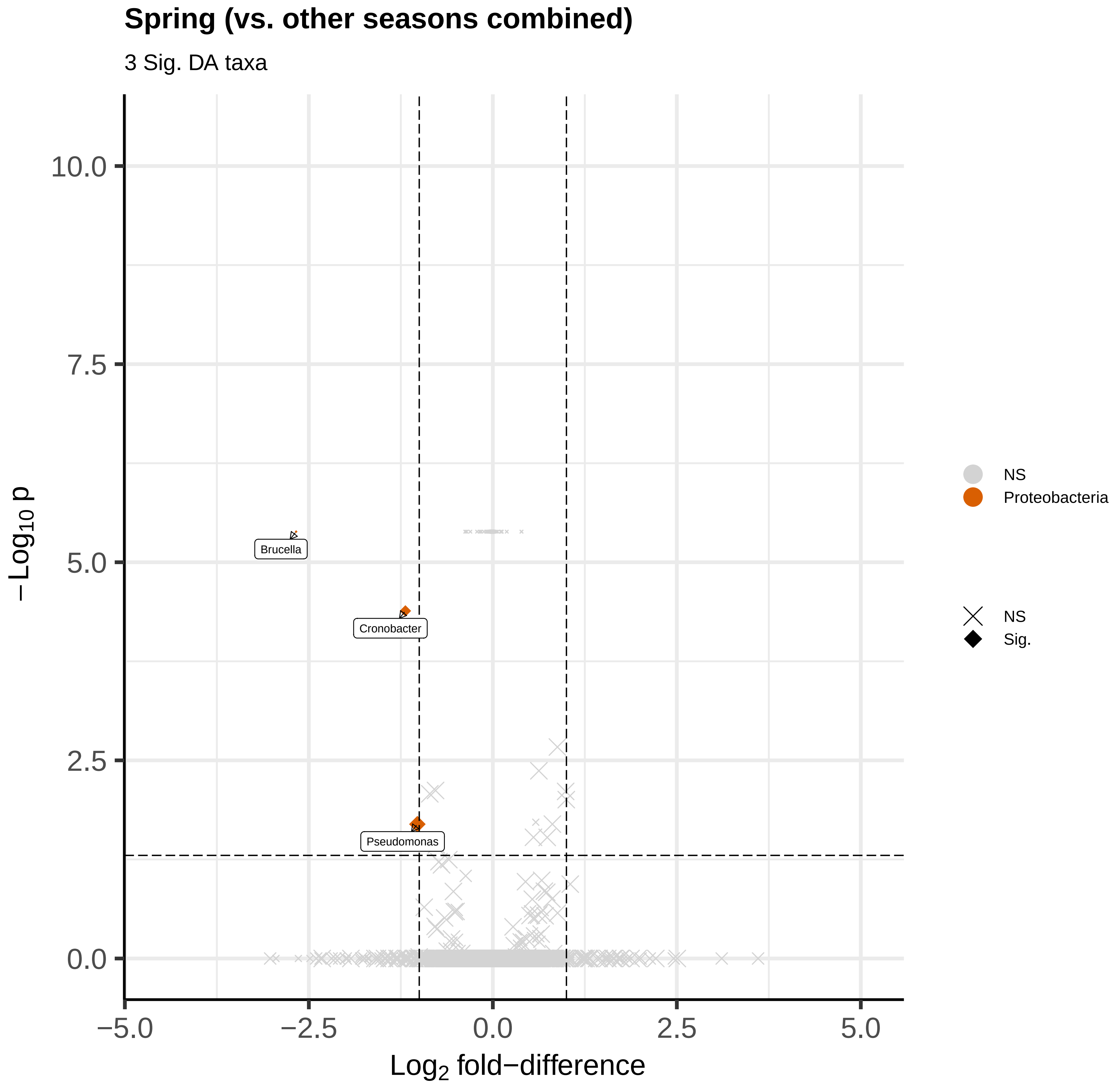
Spring (vs. other seasons combined)
3 Sig. DA taxa
Brucella
Cronobacter
Pseudomonas
10.0
7.5
5.0
2.5
0.0
−5.0
−2.5
0.0
2.5
5.0
$-Log_{10}$ p
$Log_2$ fold−difference
NS
Proteobacteria
NS
Sig.

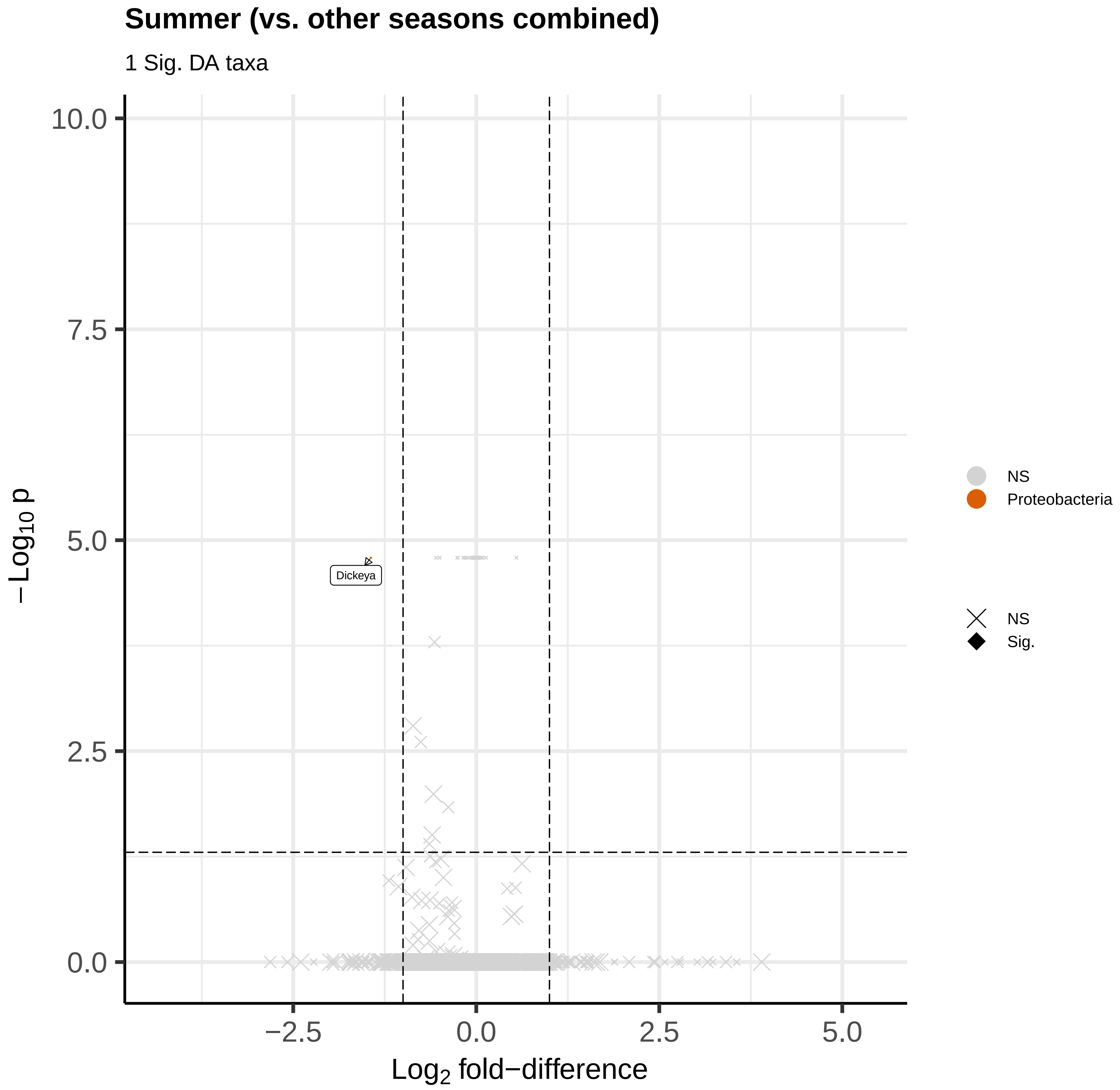

Summer (vs. other seasons combined)
1 Sig. DA taxa
10.0
7.5
5.0
2.5
0.0
−Log10 p
Dickeya
−2.5
0.0
2.5
5.0
Log2 fold−difference
NS
Proteobacteria
NS
Sig.

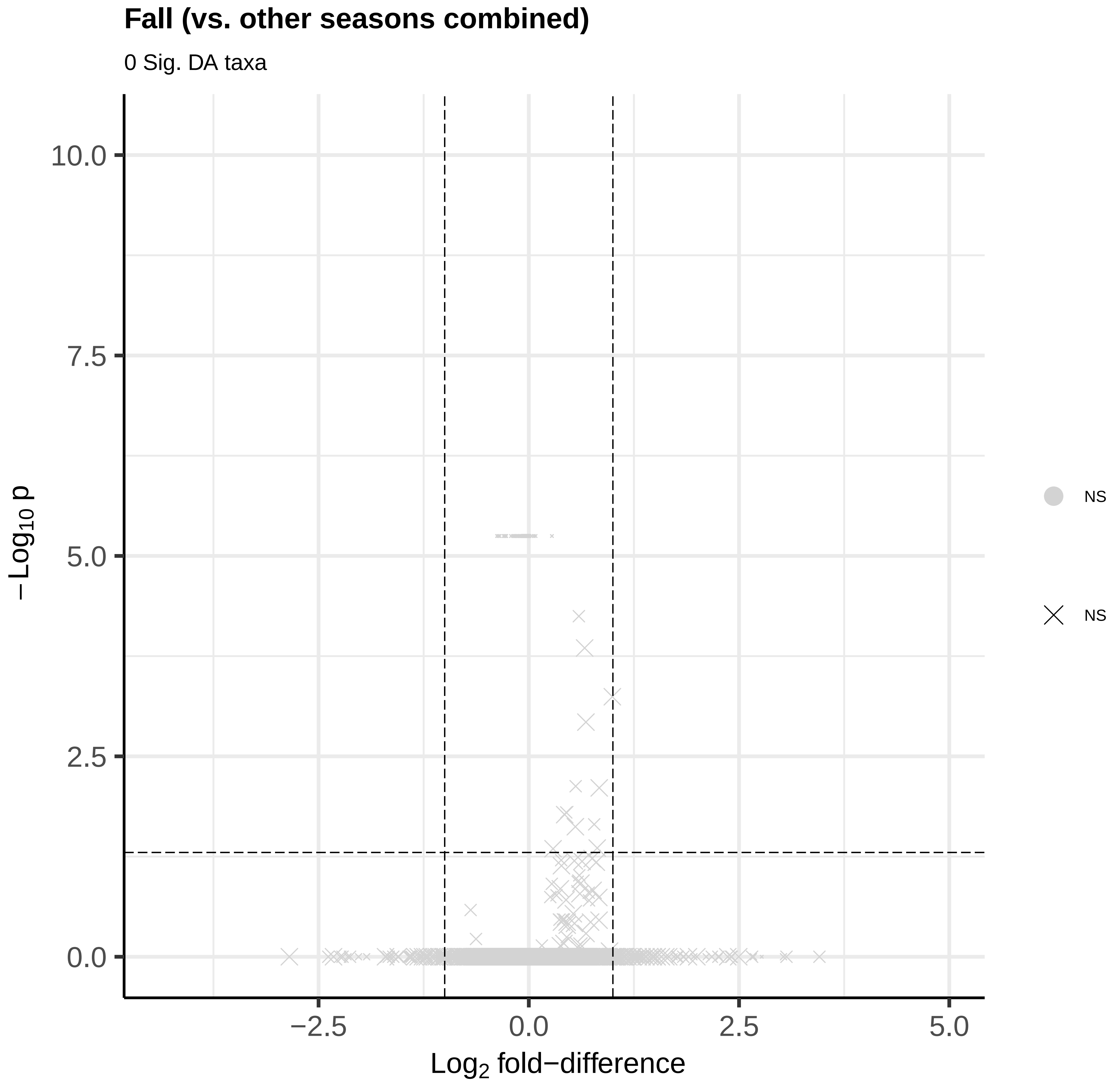
Fall (vs. other seasons combined)
0 Sig. DA taxa
10.0
7.5
5.0
2.5
0.0
−Log10 p
−2.5
0.0
2.5
5.0
Log2 fold−difference
NS
NS

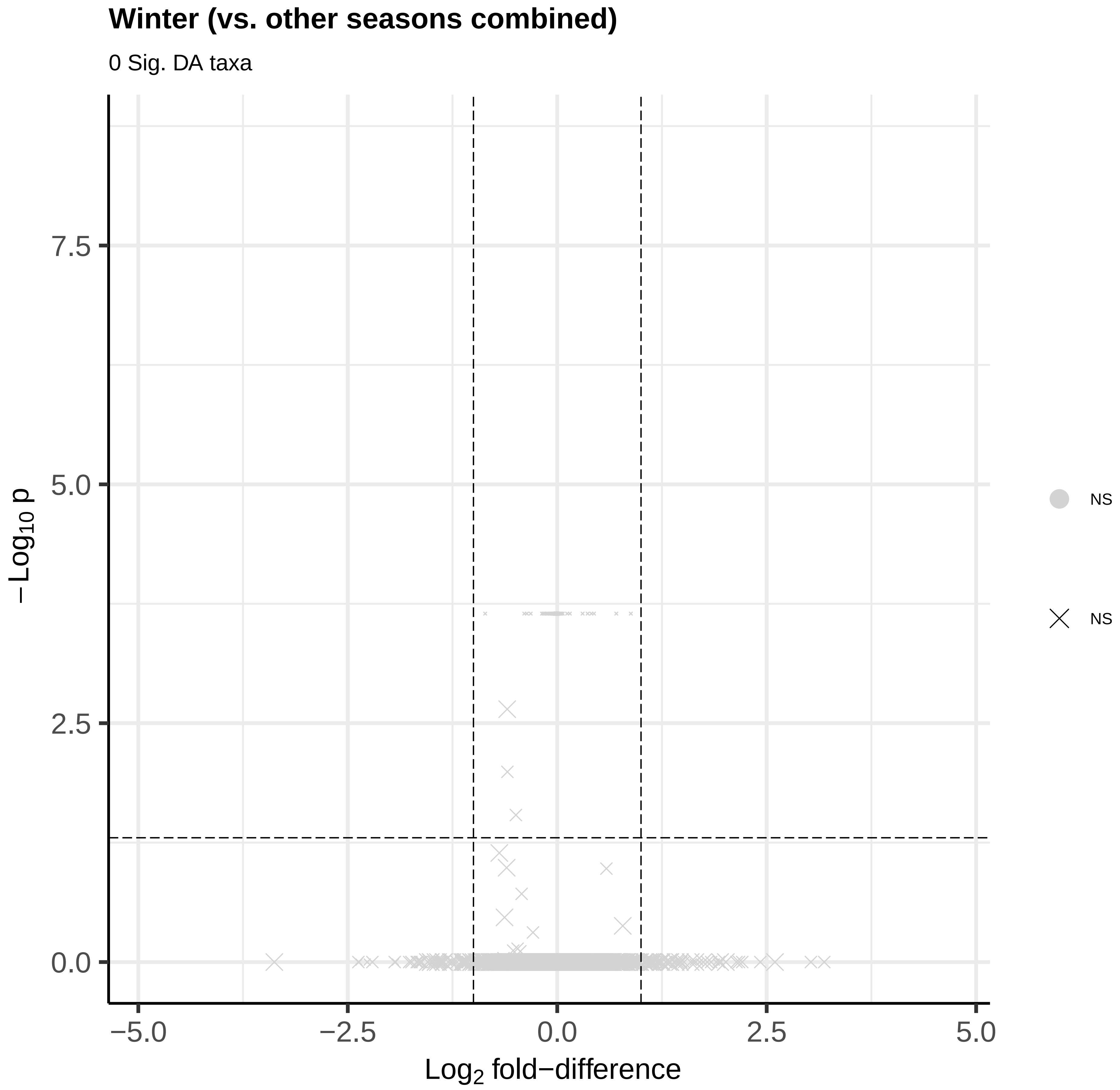
Winter (vs. other seasons combined)
0 Sig. DA taxa
7.5
5.0
2.5
0.0
−Log10 p
−5.0
−2.5
0.0
2.5
5.0
Log2 fold−difference
NS
NS
