## Supplemental figure 6 for "Metagenomics reveals novel microbial signatures of farm exposures in house dust"

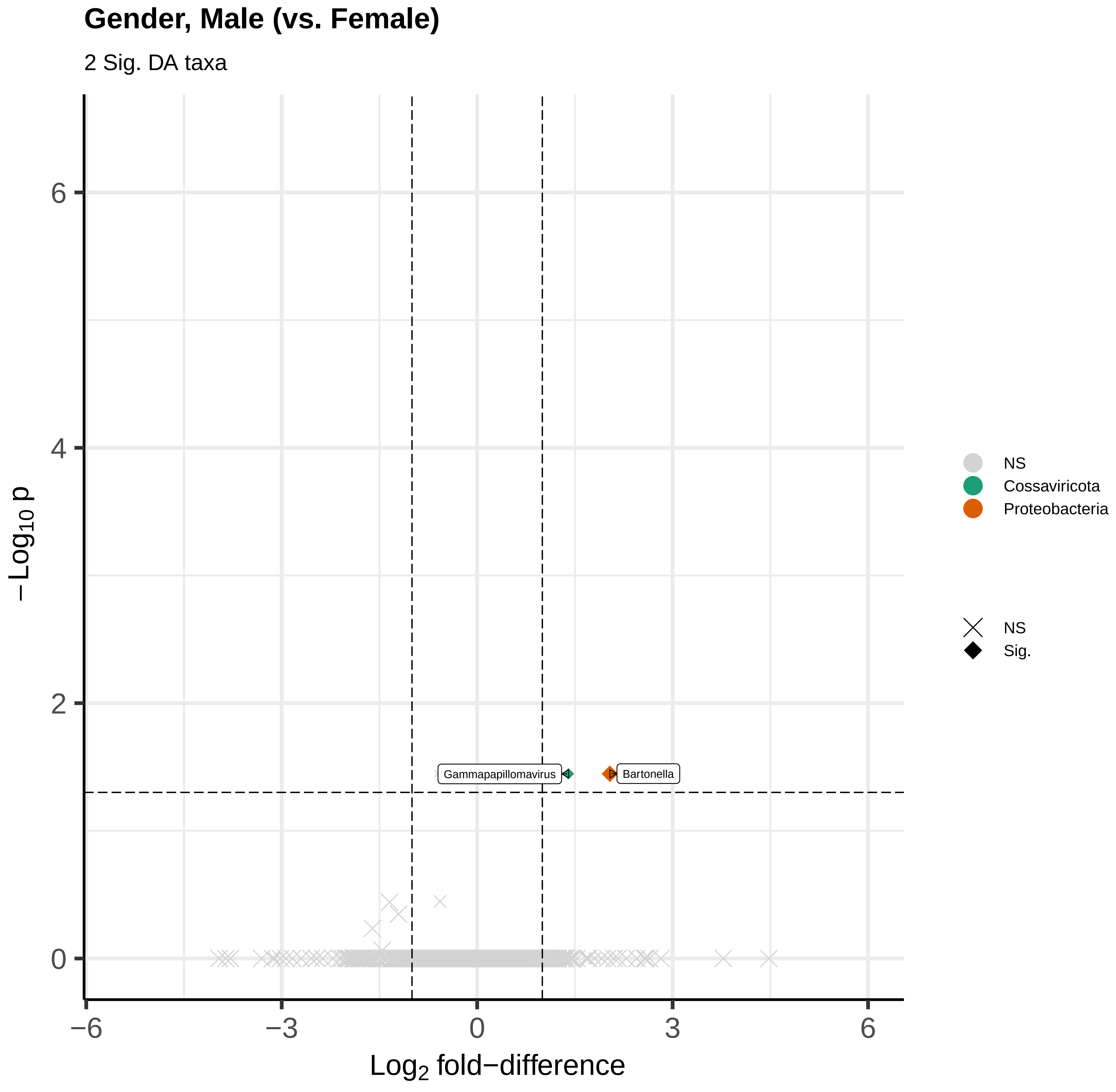
Gender, Male (vs. Female)
2 Sig. DA taxa
Gammapapillomavirus
Bartonella
−Log10 p
Log2 fold−difference
6
4
2
0
−6
−3
0
3
6
NS
Cossaviricota
Proteobacteria
NS
Sig.

**Dogs or cats (vs. neither one)**

38 Sig. DA taxa

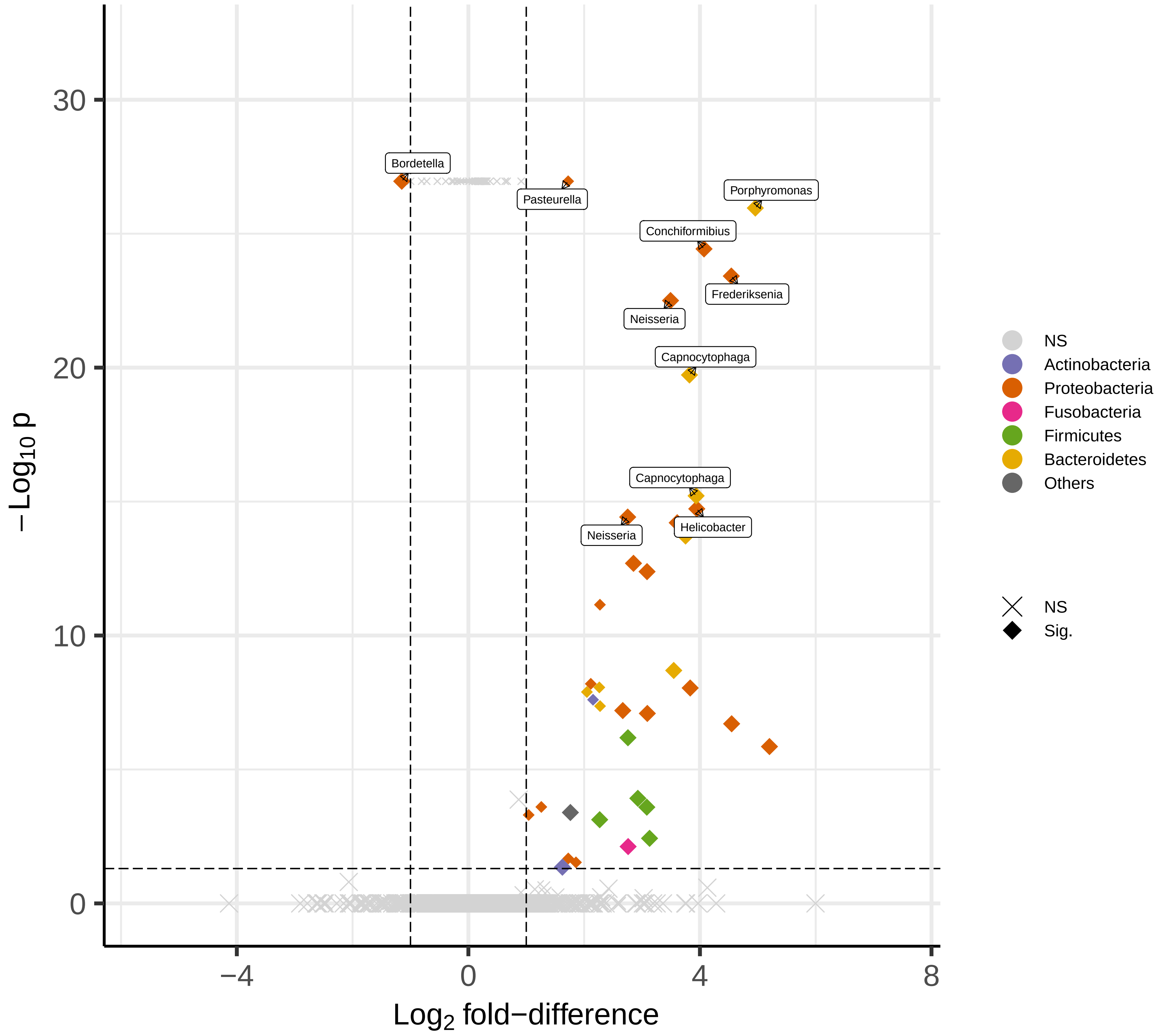

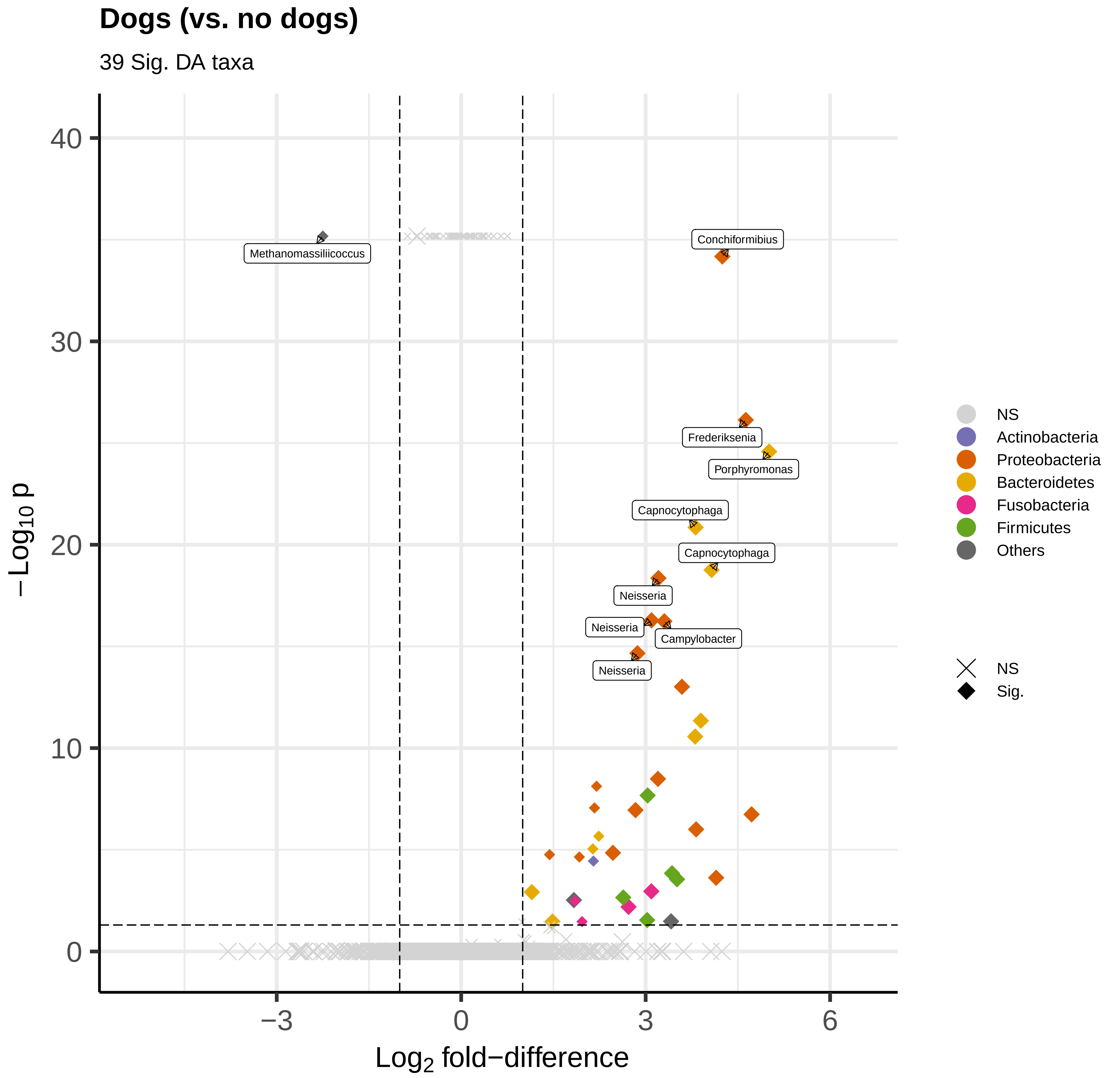

Dogs (vs. no dogs)
39 Sig. DA taxa
−Log10 p
Log2 fold−difference
Methanomassiliicoccus
Conchiformibius
Frederiksenia
Porphyromonas
Capnocytophaga
Capnocytophaga
Neisseria
Neisseria
Campylobacter
Neisseria
NS
Actinobacteria
Proteobacteria
Bacteroidetes
Fusobacteria
Firmicutes
Others
NS
Sig.

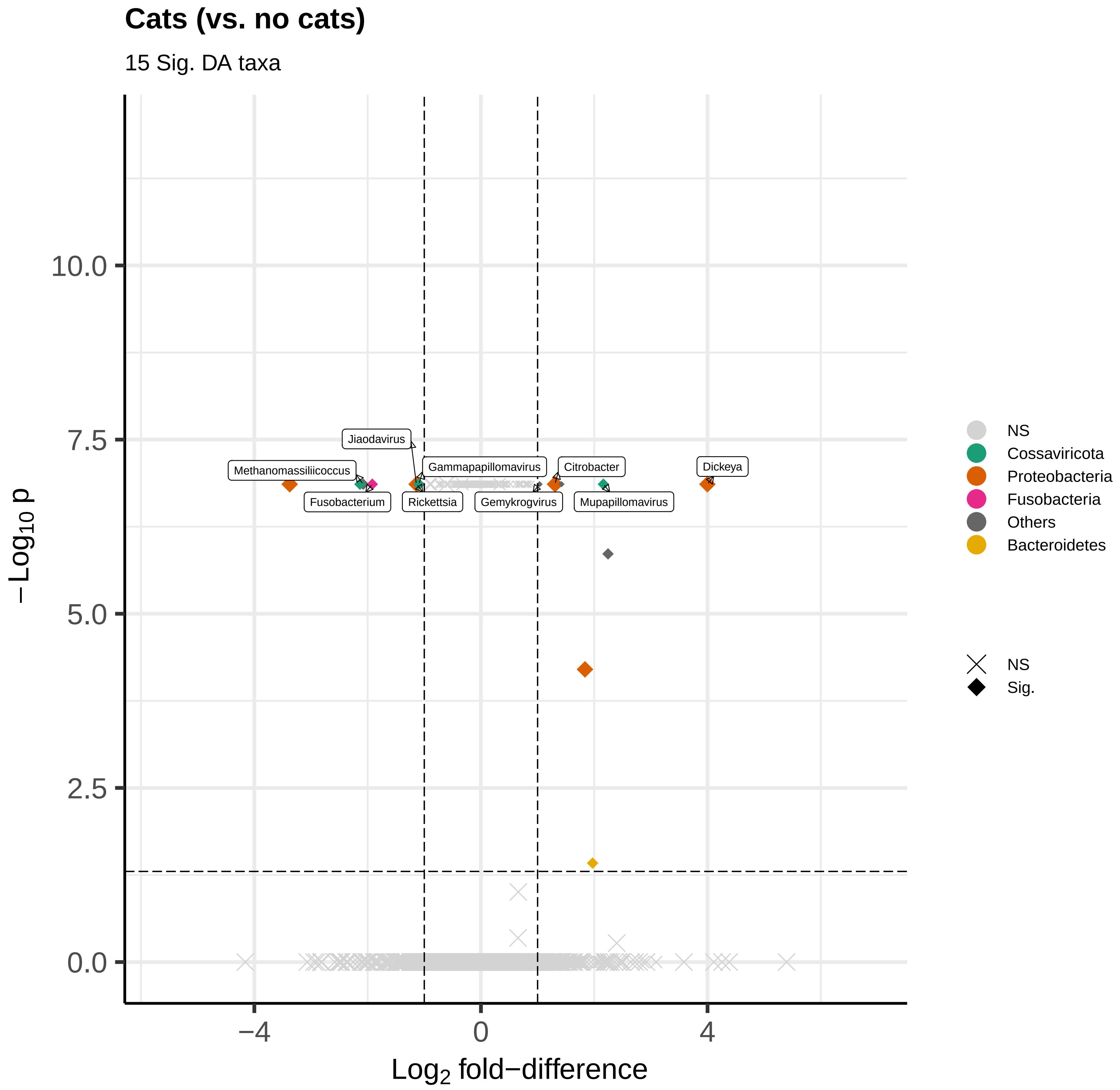
Cats (vs. no cats)
15 Sig. DA taxa
Jiaodavirus
Methanomassiliicoccus
Gammapapillomavirus
Citrobacter
Dickeya
Fusobacterium
Rickettsia
Gemykrogvirus
Mupapillomavirus
10.0
7.5
5.0
2.5
0.0
−4
0
4
−Log10 p
Log2 fold−difference
NS
Cossaviricota
Proteobacteria
Fusobacteria
Others
Bacteroidetes
NS
Sig.

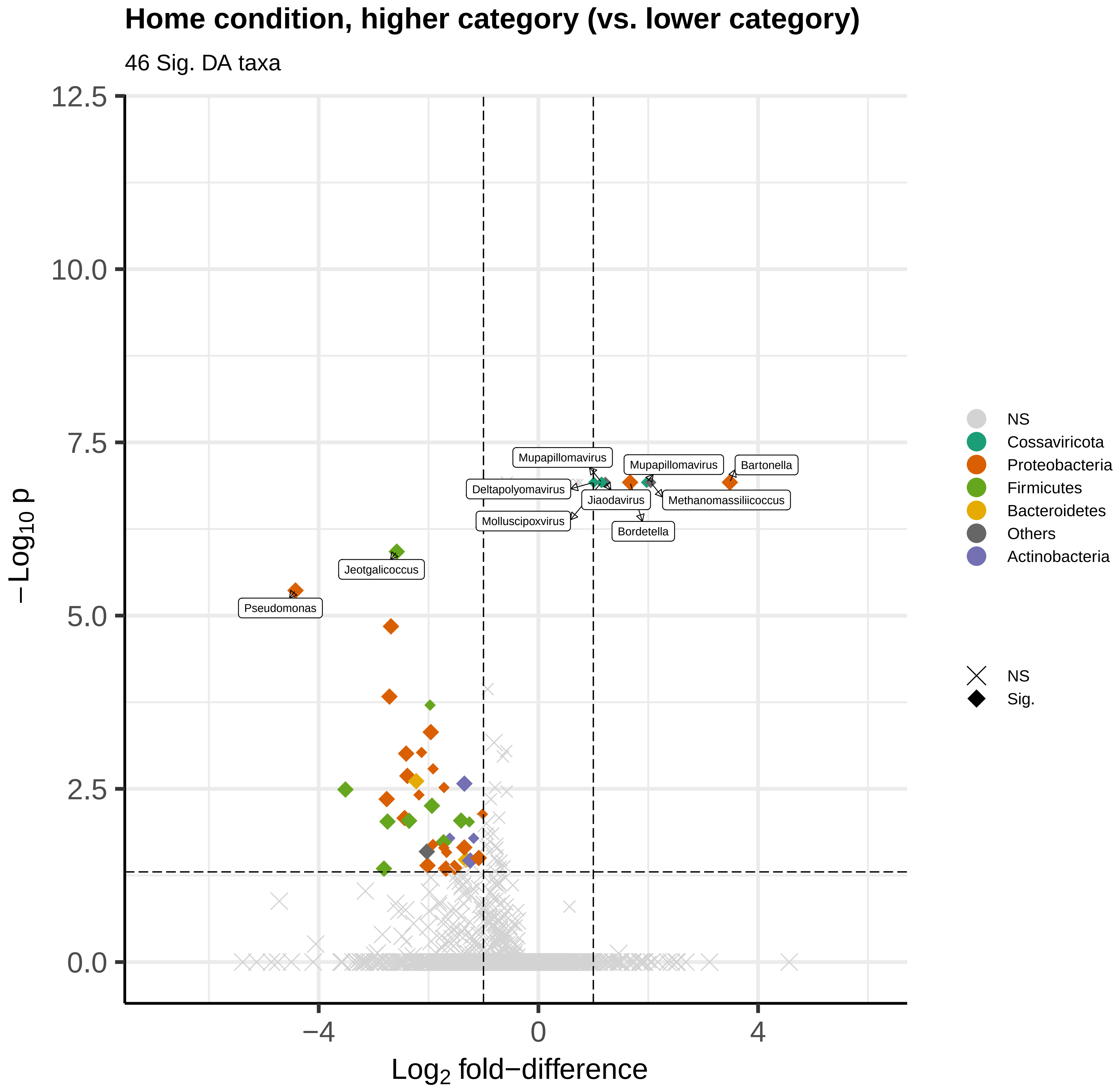

Home condition, higher category (vs. lower category)
46 Sig. DA taxa
12.5
10.0
7.5
5.0
2.5
0.0
−Log10 p
−4
0
4
Log2 fold−difference
Mupapillomavirus
Mupapillomavirus
Bartonella
Deltapolyomavirus
Jiaodavirus
Methanomassiliicoccus
Molluscipoxvirus
Bordetella
Jeotgalicoccus
Pseudomonas
NS
Cossaviricota
Proteobacteria
Firmicutes
Bacteroidetes
Others
Actinobacteria
NS
Sig.

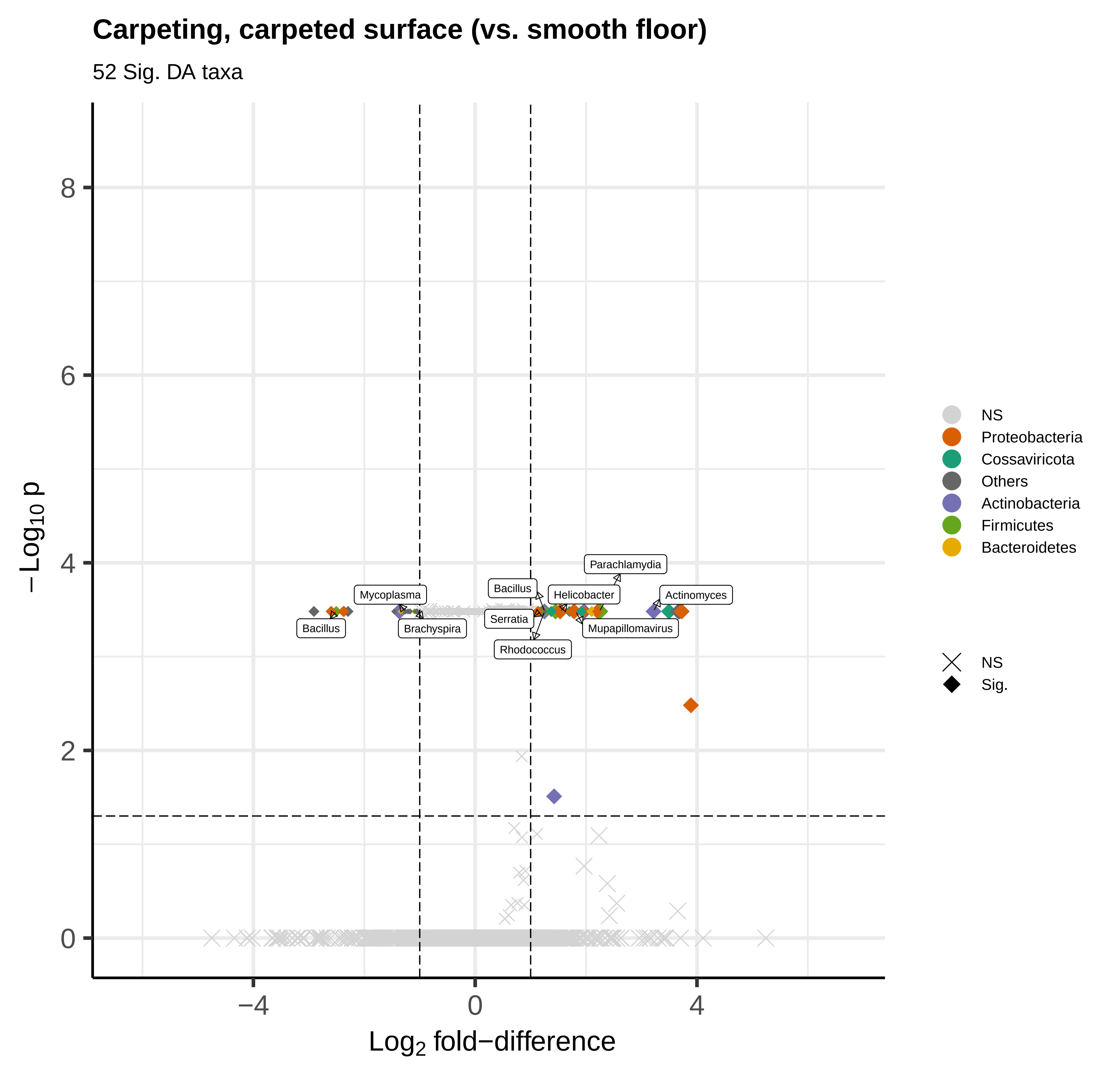

Carpeting, carpeted surface (vs. smooth floor)
52 Sig. DA taxa
−Log10 p
Log2 fold−difference
8
6
4
2
0
−4
0
4
Parachlamydia
Mycoplasma
Bacillus
Helicobacter
Actinomyces
Serratia
Bacillus
Brachyspira
Mupapillomavirus
Rhodococcus
NS
Proteobacteria
Cossaviricota
Others
Actinobacteria
Firmicutes
Bacteroidetes
NS
Sig.

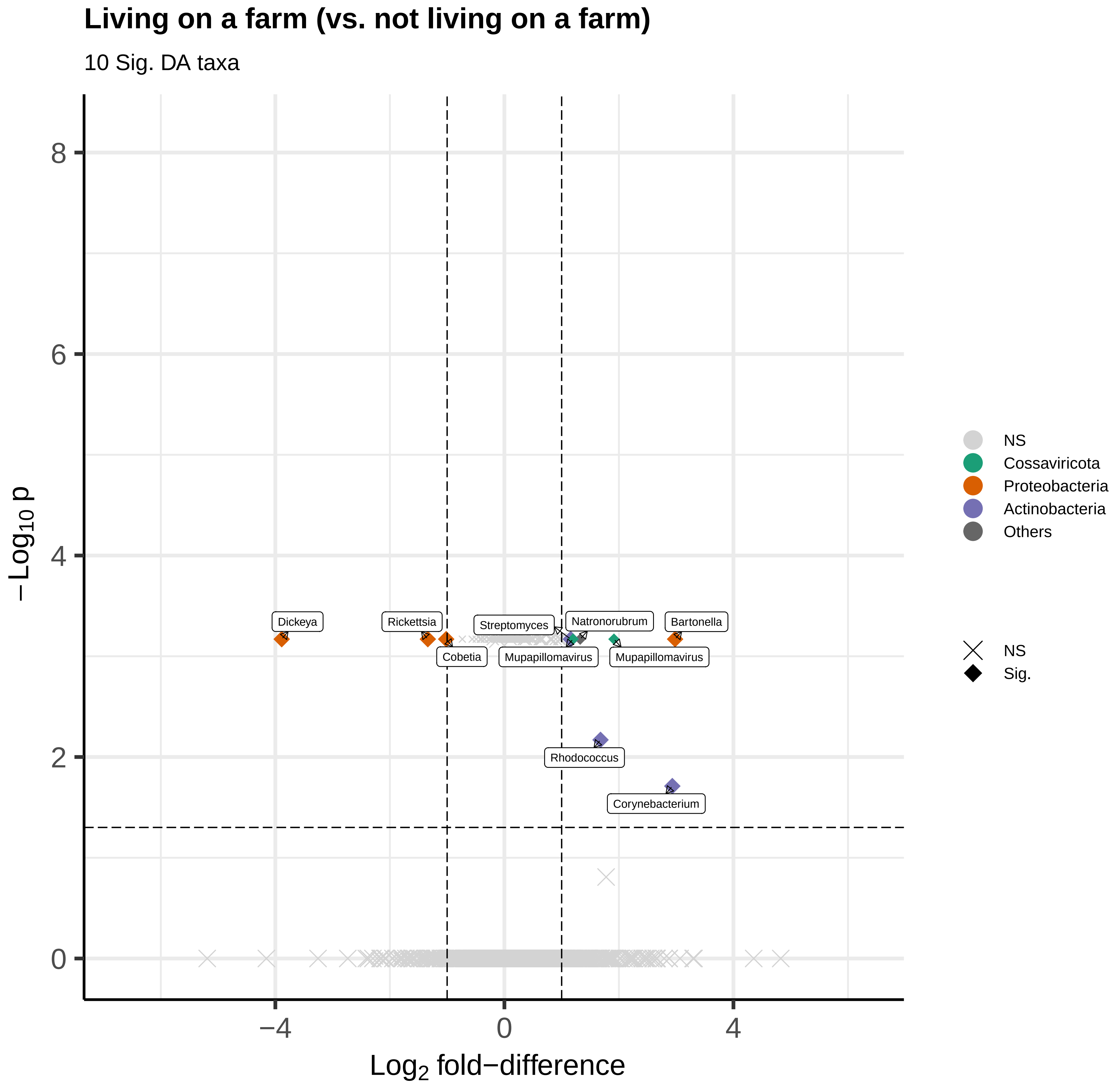

Living on a farm (vs. not living on a farm)
10 Sig. DA taxa
−Log10 p
Log2 fold−difference
8
6
4
2
0
−4
0
4
Dickeya
Rickettsia
Cobetia
Streptomyces
Natronorubrum
Mupapillomavirus
Mupapillomavirus
Bartonella
Rhodococcus
Corynebacterium
NS
Cossaviricota
Proteobacteria
Actinobacteria
Others
NS
Sig.

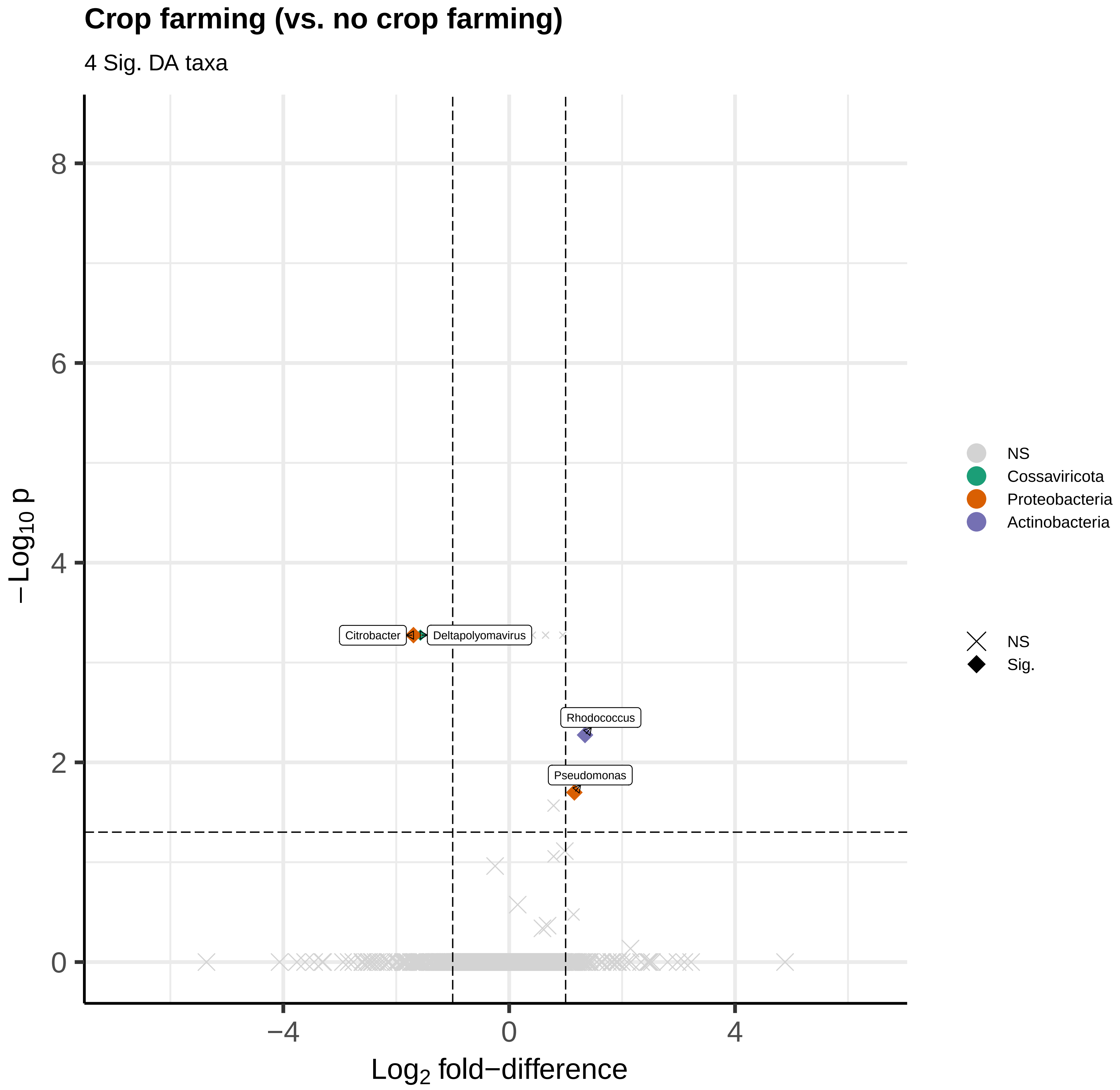

Crop farming (vs. no crop farming)
4 Sig. DA taxa
−Log10 p
Log2 fold−difference
8
6
4
2
0
−4
0
4
Citrobacter
Deltapolyomavirus
Rhodococcus
Pseudomonas
NS
Cossaviricota
Proteobacteria
Actinobacteria
NS
Sig.

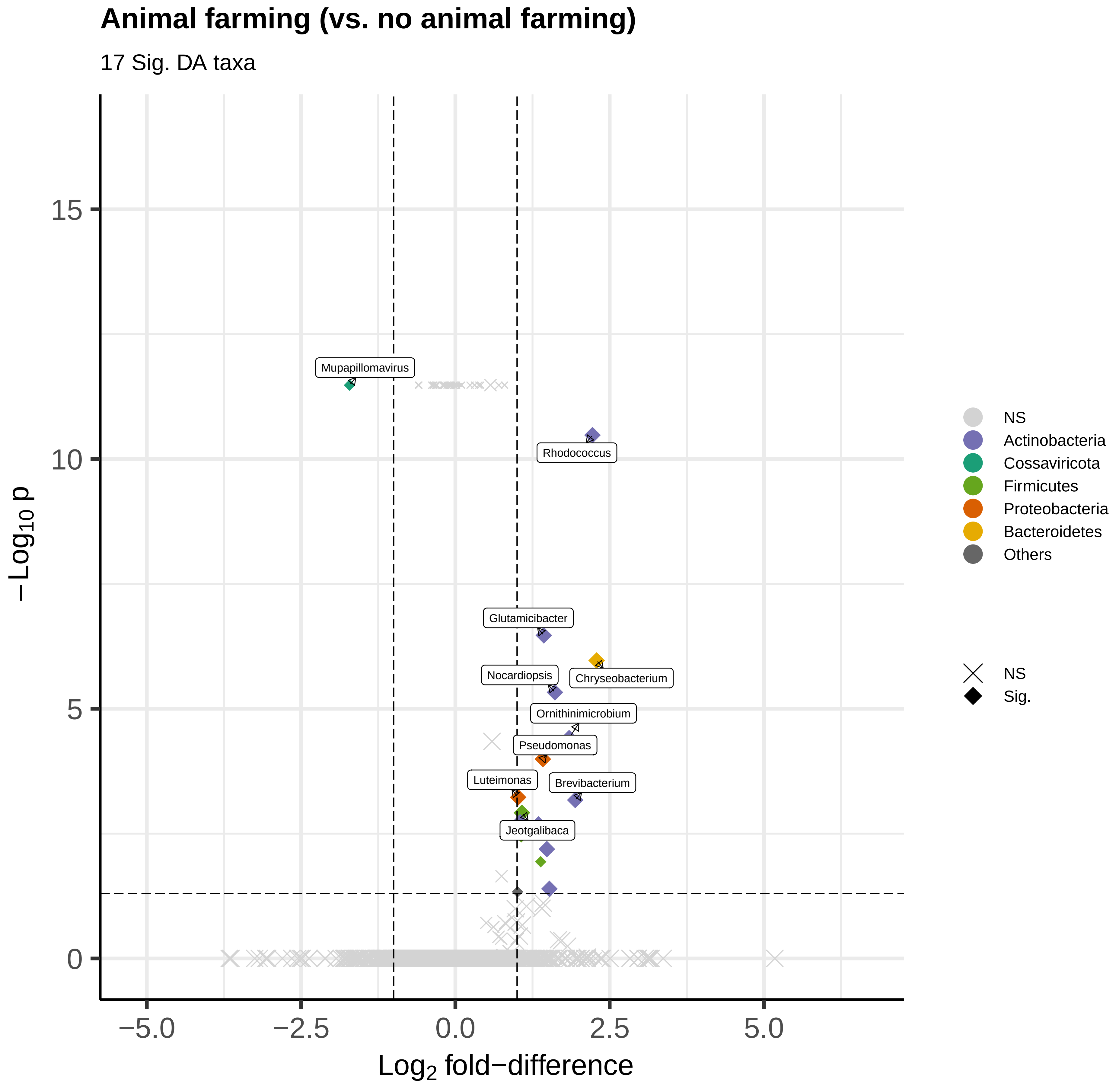

Animal farming (vs. no animal farming)
17 Sig. DA taxa
Mupapillomavirus
Rhodococcus
Glutamicibacter
Chryseobacterium
Nocardiopsis
Ornithinimicrobium
Pseudomonas
Luteimonas
Brevibacterium
Jeotgalibaca
15
10
5
0
−Log10 p
−5.0
−2.5
0.0
2.5
5.0
Log2 fold−difference
NS
Actinobacteria
Cossaviricota
Firmicutes
Proteobacteria
Bacteroidetes
Others
NS
Sig.

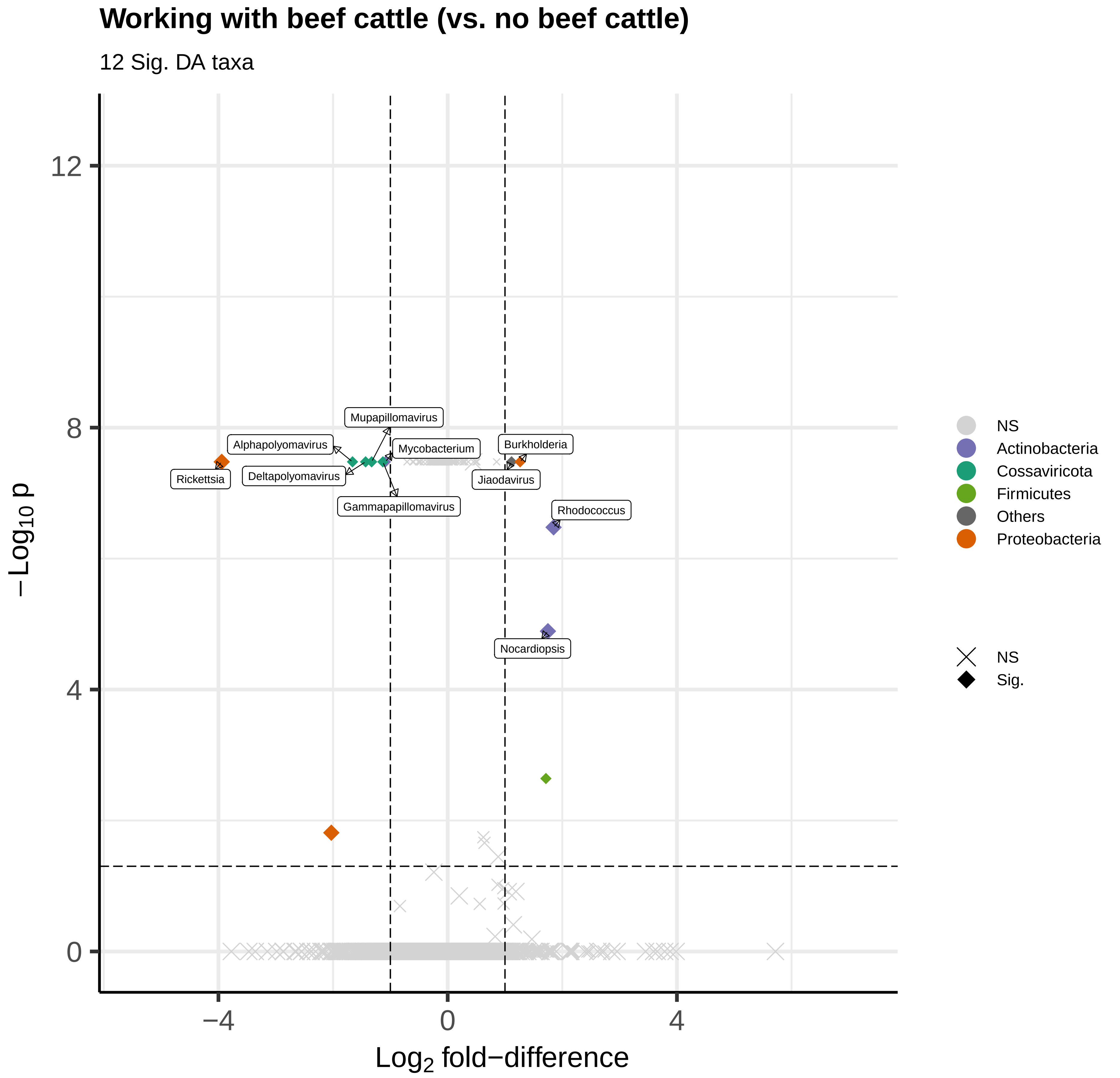
Working with beef cattle (vs. no beef cattle)
12 Sig. DA taxa
12
8
4
0
−4
0
4
−Log10 p
Log2 fold−difference
Mupapillomavirus
Alphapolyomavirus
Mycobacterium
Burkholderia
Rickettsia
Deltapolyomavirus
Jiaodavirus
Gammapapillomavirus
Rhodococcus
Nocardiopsis
NS
Actinobacteria
Cossaviricota
Firmicutes
Others
Proteobacteria
NS
Sig.

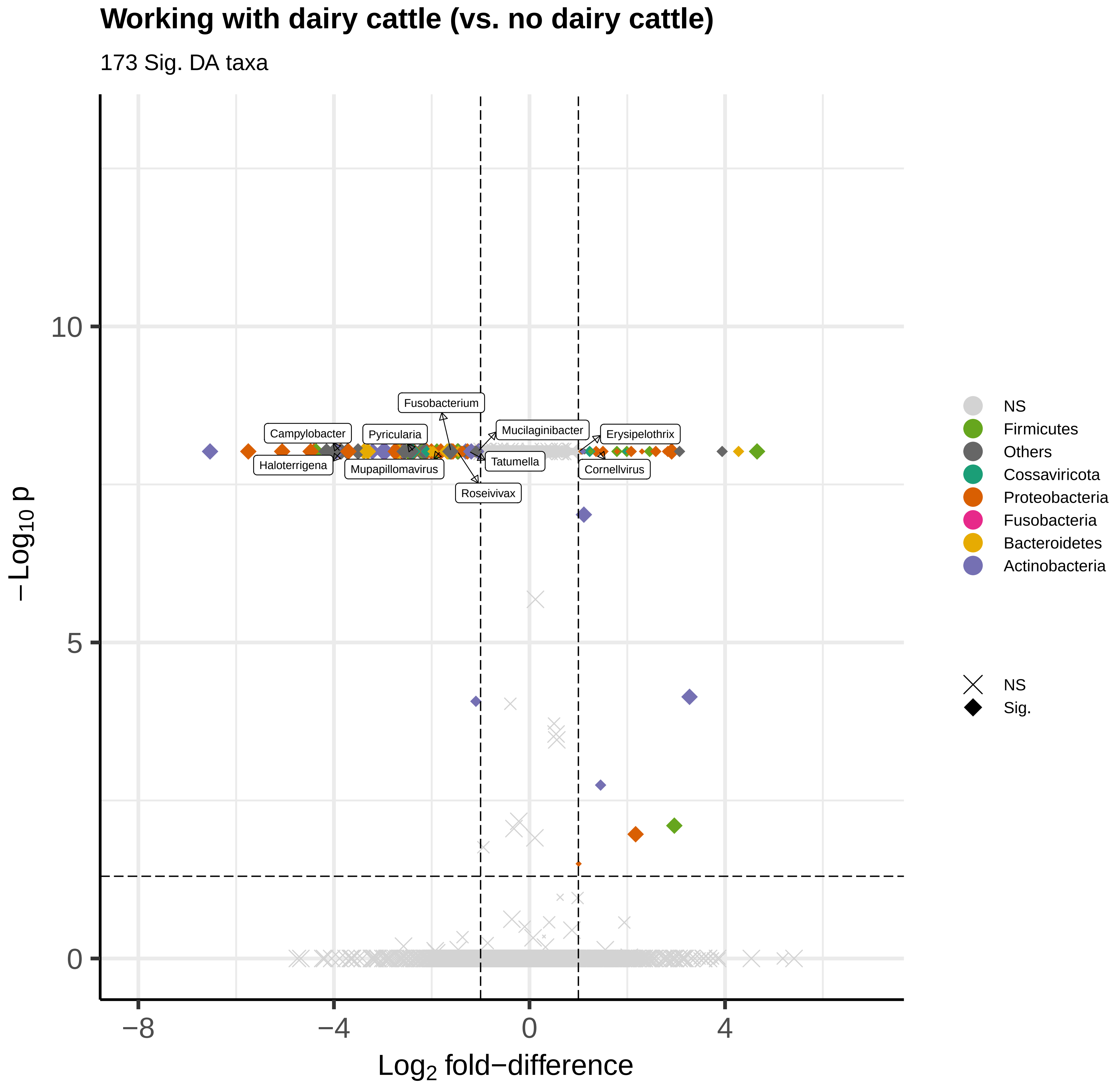
Working with dairy cattle (vs. no dairy cattle)
173 Sig. DA taxa
Fusobacterium
Campylobacter
Pyricularia
Mucilaginibacter
Erysipelothrix
Haloterrigena
Mupapillomavirus
Tatumella
Cornellvirus
Roseivivax
10
5
0
−8
−4
0
4
−Log10 p
Log2 fold−difference
NS
Firmicutes
Others
Cossaviricota
Proteobacteria
Fusobacteria
Bacteroidetes
Actinobacteria
NS
Sig.

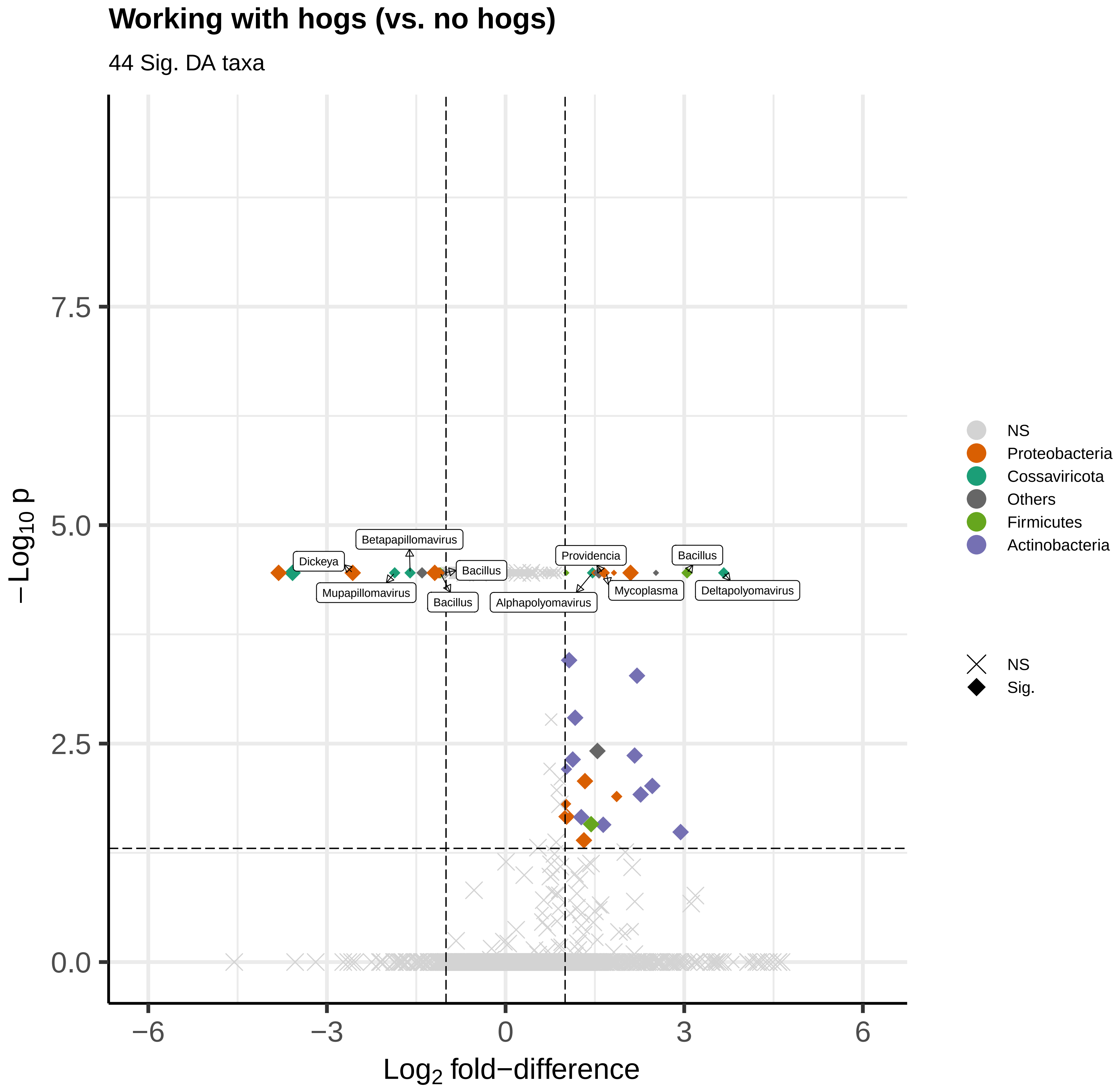
Working with hogs (vs. no hogs)
44 Sig. DA taxa
Betapapillomavirus
Dickeya
Bacillus
Mupapillomavirus
Bacillus
Providencia
Bacillus
Mycoplasma
Deltapolyomavirus
Alphapolyomavirus
0.0
2.5
5.0
7.5
−6
−3
0
3
6
$-\log_{10}$ p
$\log_2$ fold−difference
NS
Proteobacteria
Cossaviricota
Others
Firmicutes
Actinobacteria
NS
Sig.

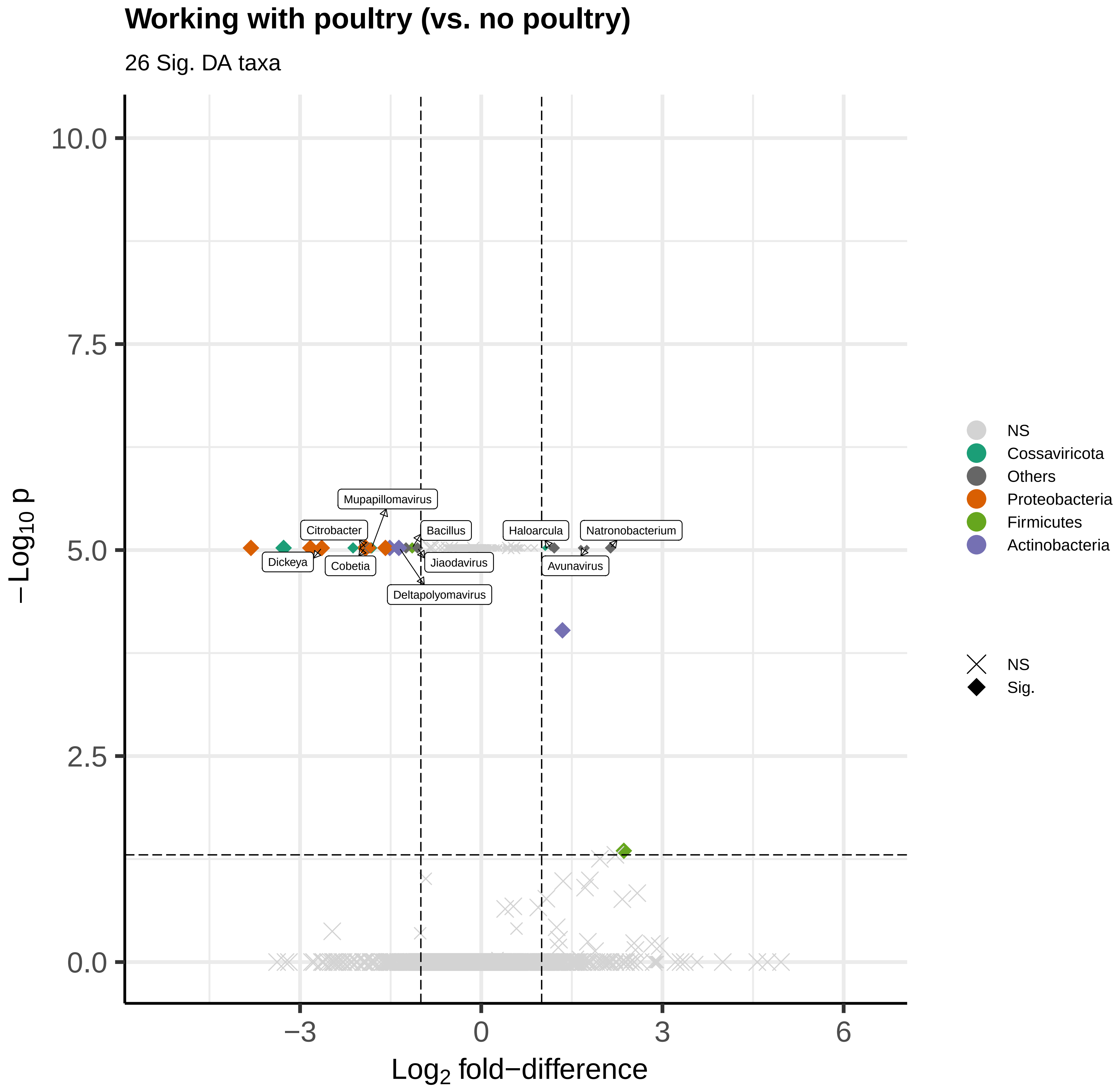

Working with poultry (vs. no poultry)
26 Sig. DA taxa
−Log10 p
Log2 fold−difference
10.0
7.5
5.0
2.5
0.0
−3
0
3
6
Mupapillomavirus
Citrobacter
Bacillus
Haloarcula
Natronobacterium
Dickeya
Cobetia
Jiaodavirus
Avunavirus
Deltapolyomavirus
NS
Cossaviricota
Others
Proteobacteria
Firmicutes
Actinobacteria
NS
Sig.

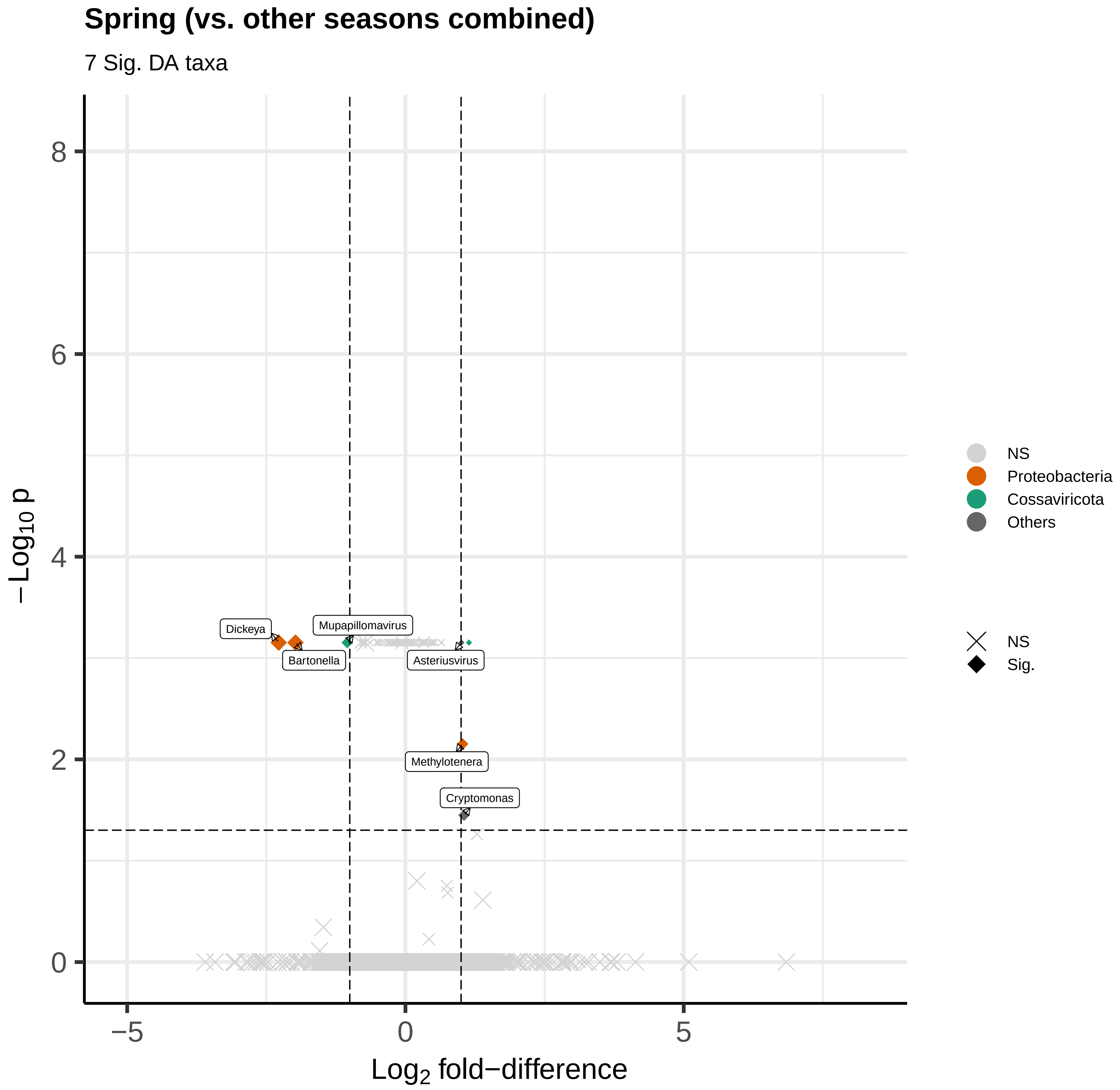
Spring (vs. other seasons combined)
7 Sig. DA taxa
−Log10 p
Log2 fold−difference
8
6
4
2
0
−5
0
5
Dickeya
Mupapillomavirus
Bartonella
Asteriusvirus
Methylotenera
Cryptomonas
NS
Proteobacteria
Cossaviricota
Others
NS
Sig.

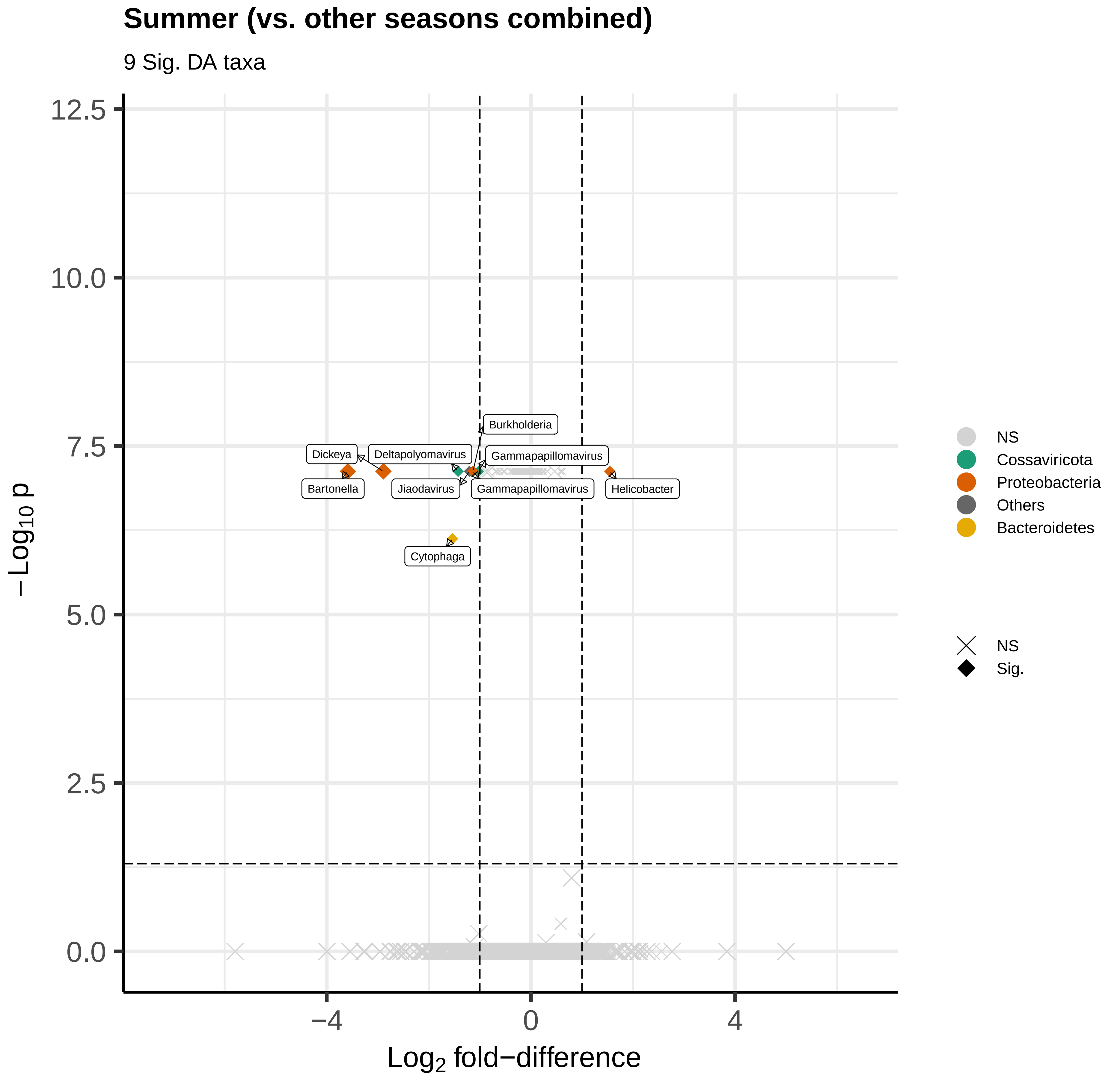

Summer (vs. other seasons combined)
9 Sig. DA taxa
12.5
10.0
7.5
5.0
2.5
0.0
−Log10 p
−4
0
4
Log2 fold−difference
Burkholderia
Dickeya
Deltapolyomavirus
Gammapapillomavirus
Bartonella
Jiaodavirus
Gammapapillomavirus
Helicobacter
Cytophaga
NS
Cossaviricota
Proteobacteria
Others
Bacteroidetes
NS
Sig.

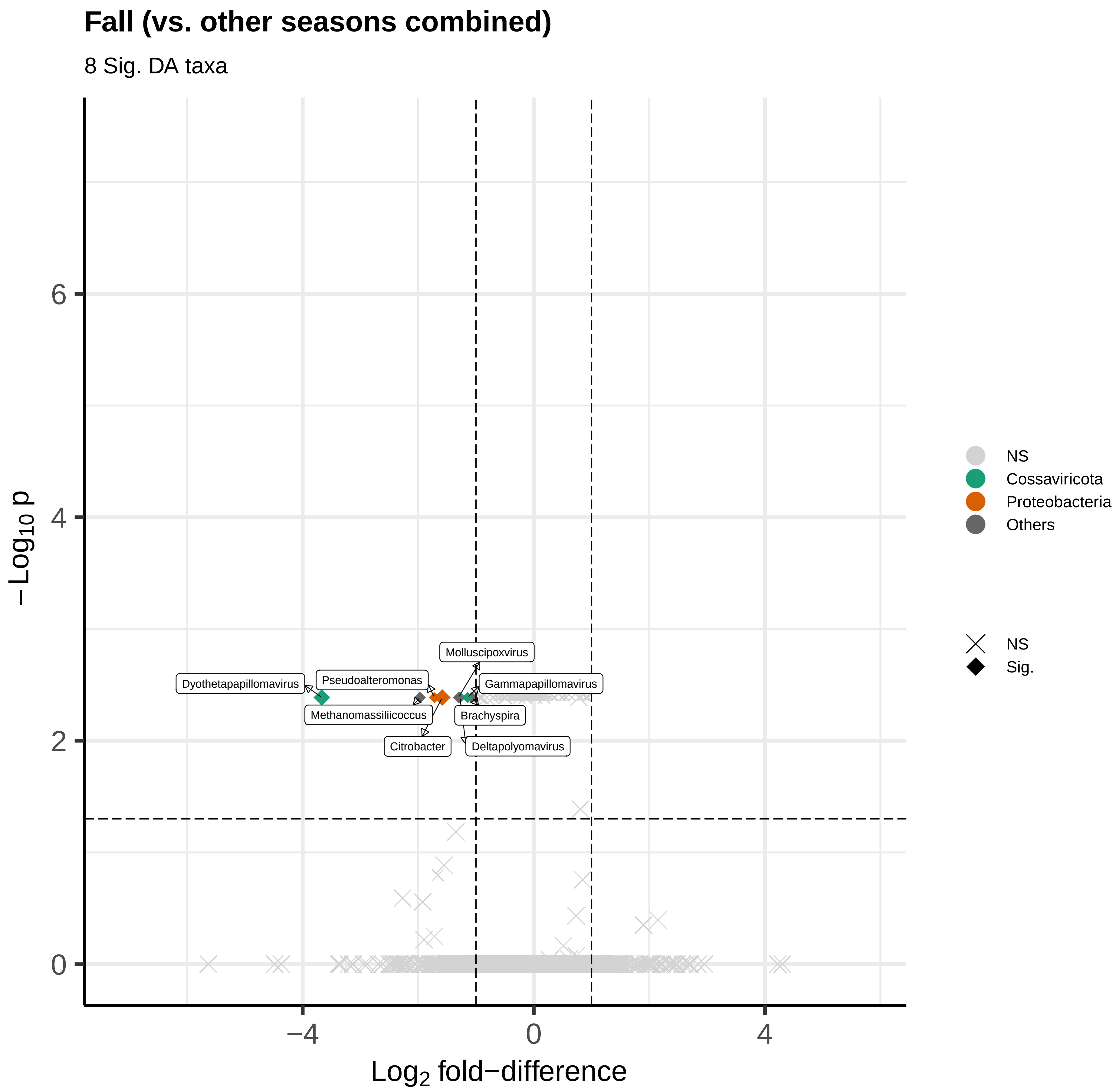

Fall (vs. other seasons combined)
8 Sig. DA taxa
−Log10 p
Log2 fold−difference
0
2
4
6
−4
0
4
Dyothetapapillomavirus
Pseudoalteromonas
Methanomassiliicoccus
Citrobacter
Molluscipoxvirus
Gammapapillomavirus
Brachyspira
Deltapolyomavirus
NS
Cossaviricota
Proteobacteria
Others
NS
Sig.

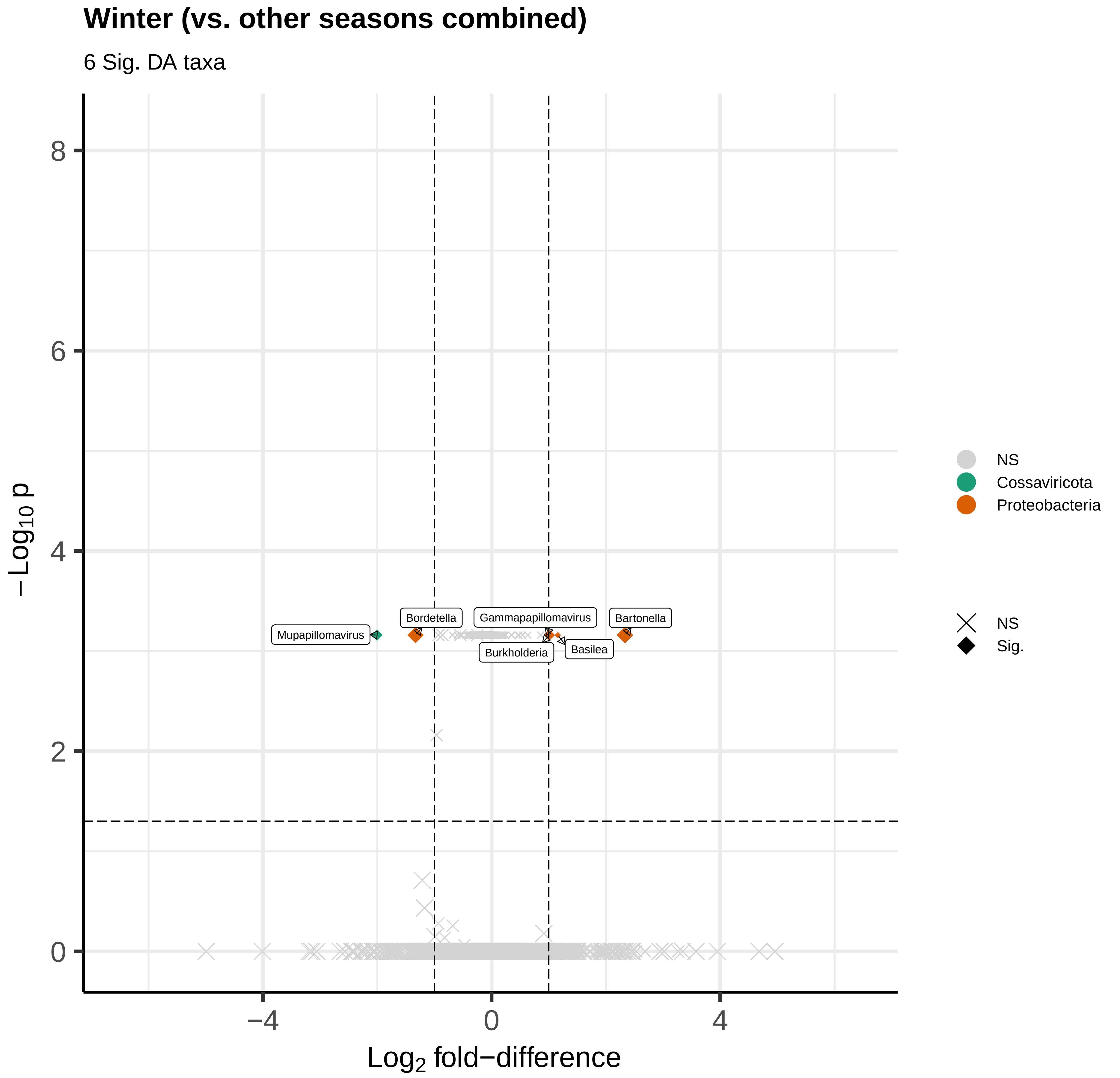
Winter (vs. other seasons combined)
6 Sig. DA taxa
−Log10 p
8
6
4
2
0
−4
0
4
Log2 fold−difference
Mupapillomavirus
Bordetella
Gammapapillomavirus
Bartonella
Burkholderia
Basilea
NS
Cossaviricota
Proteobacteria
NS
Sig.
