## Supplemental figure 7 for "Metagenomics reveals novel microbial signatures of farm exposures in house dust"

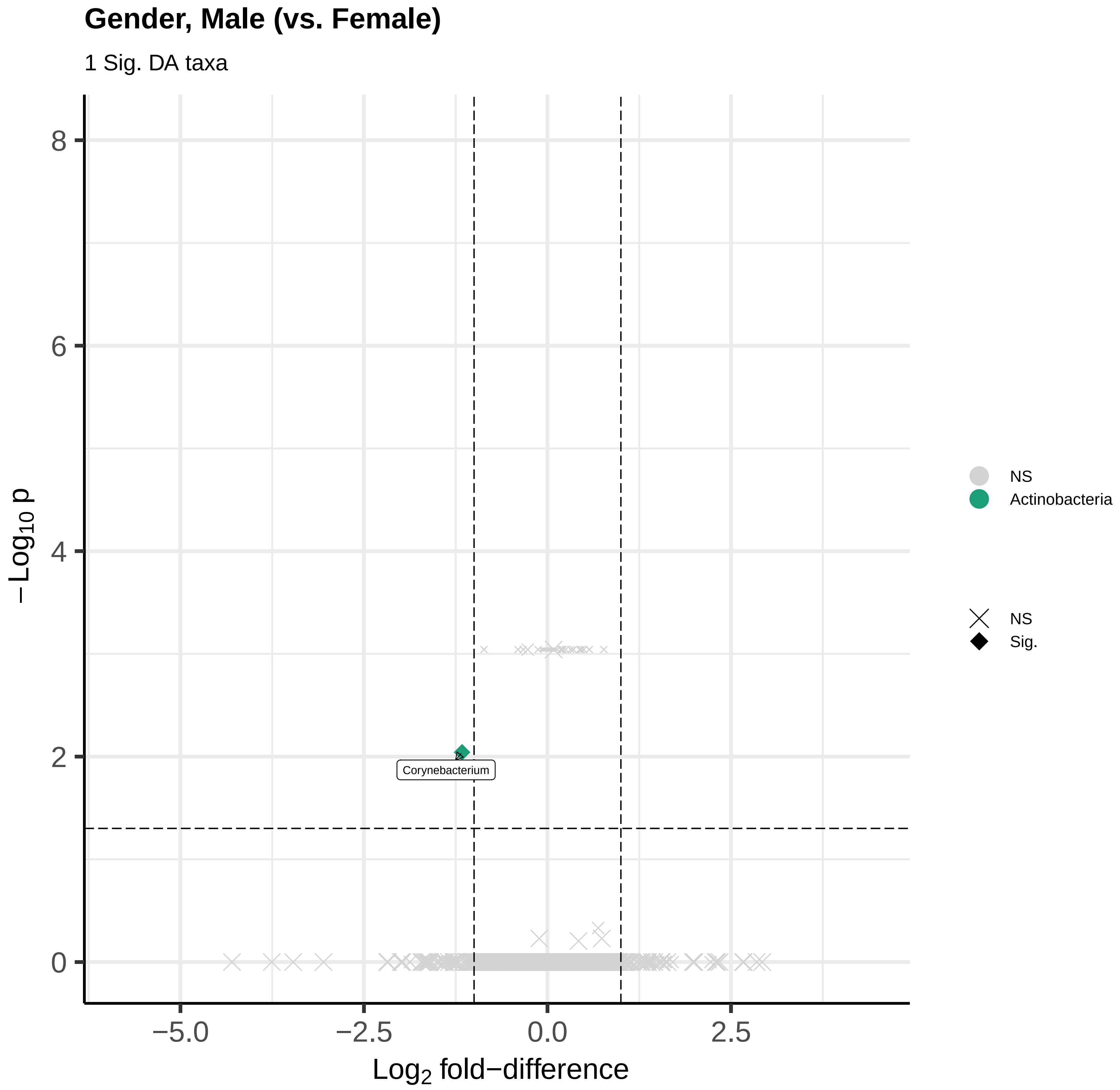
Gender, Male (vs. Female)
1 Sig. DA taxa
8
6
4
2
0
−Log10 p
Corynebacterium
−5.0
−2.5
0.0
2.5
Log2 fold−difference
NS
Actinobacteria
NS
Sig.

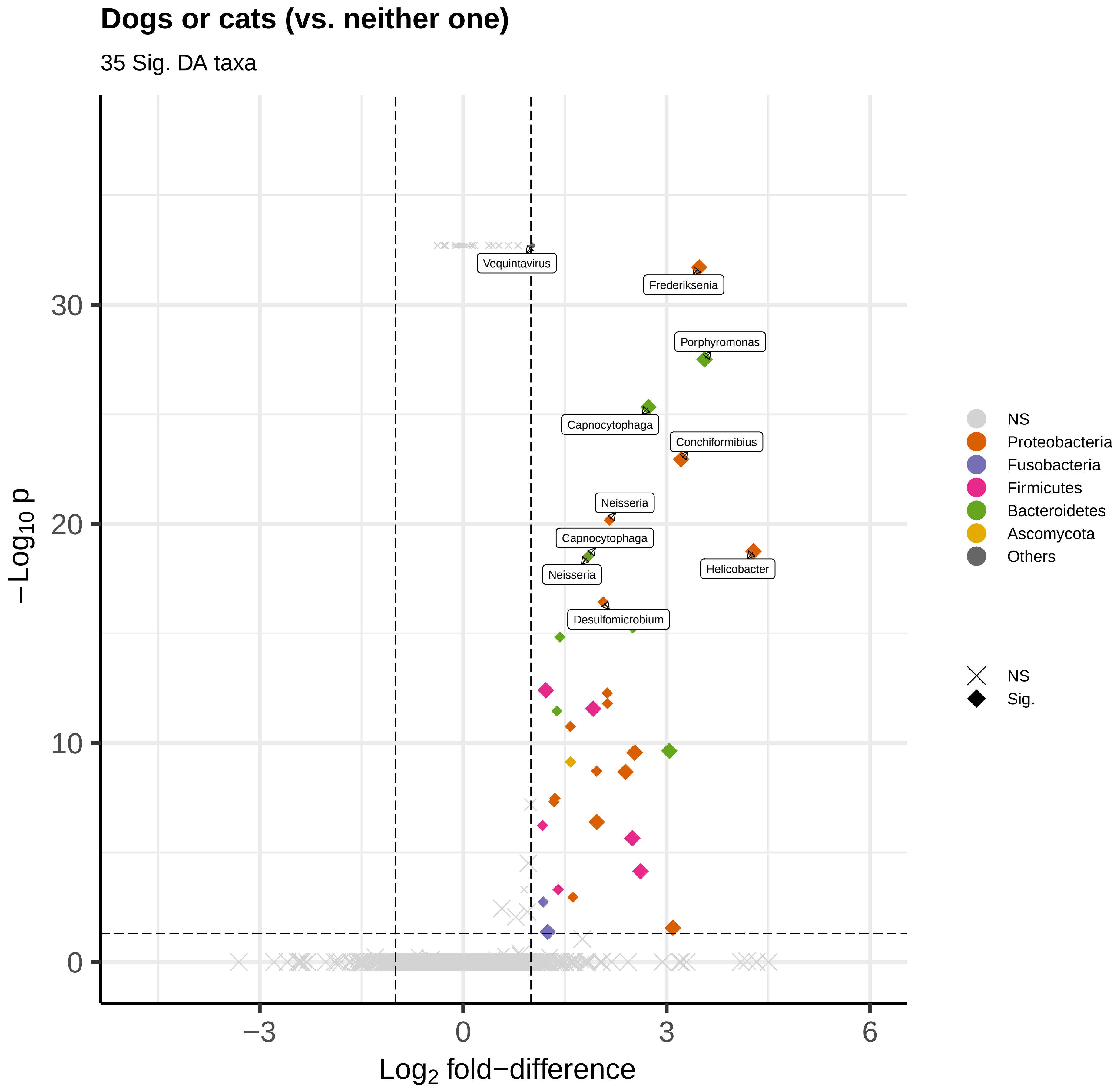

Dogs or cats (vs. neither one)
35 Sig. DA taxa
Vequintavirus
Frederiksenia
Porphyromonas
Capnocytophaga
Conchiformibius
Neisseria
Capnocytophaga
Neisseria
Helicobacter
Desulfomicrobium
30
20
10
0
−3
0
3
6
Log2 fold−difference
−Log10 p
NS
Proteobacteria
Fusobacteria
Firmicutes
Bacteroidetes
Ascomycota
Others
NS
Sig.

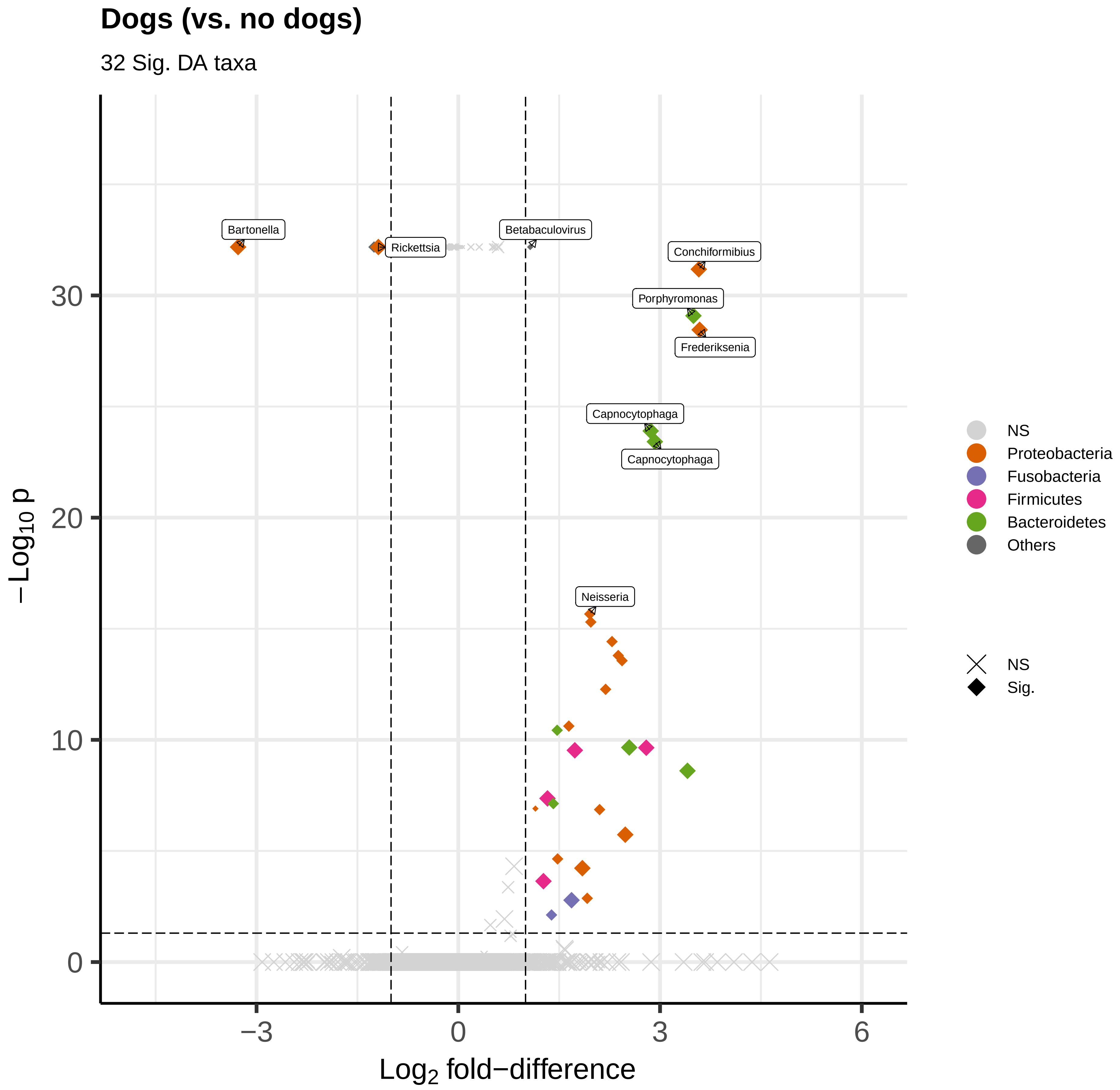

Dogs (vs. no dogs)
32 Sig. DA taxa
Bartonella
Rickettsia
Betabaculovirus
Conchiformibius
Porphyromonas
Frederiksenia
Capnocytophaga
Capnocytophaga
Neisseria
30
20
10
0
−3
0
3
6
$-Log_{10}$ p
$Log_2$ fold−difference
NS
Proteobacteria
Fusobacteria
Firmicutes
Bacteroidetes
Others
NS
Sig.

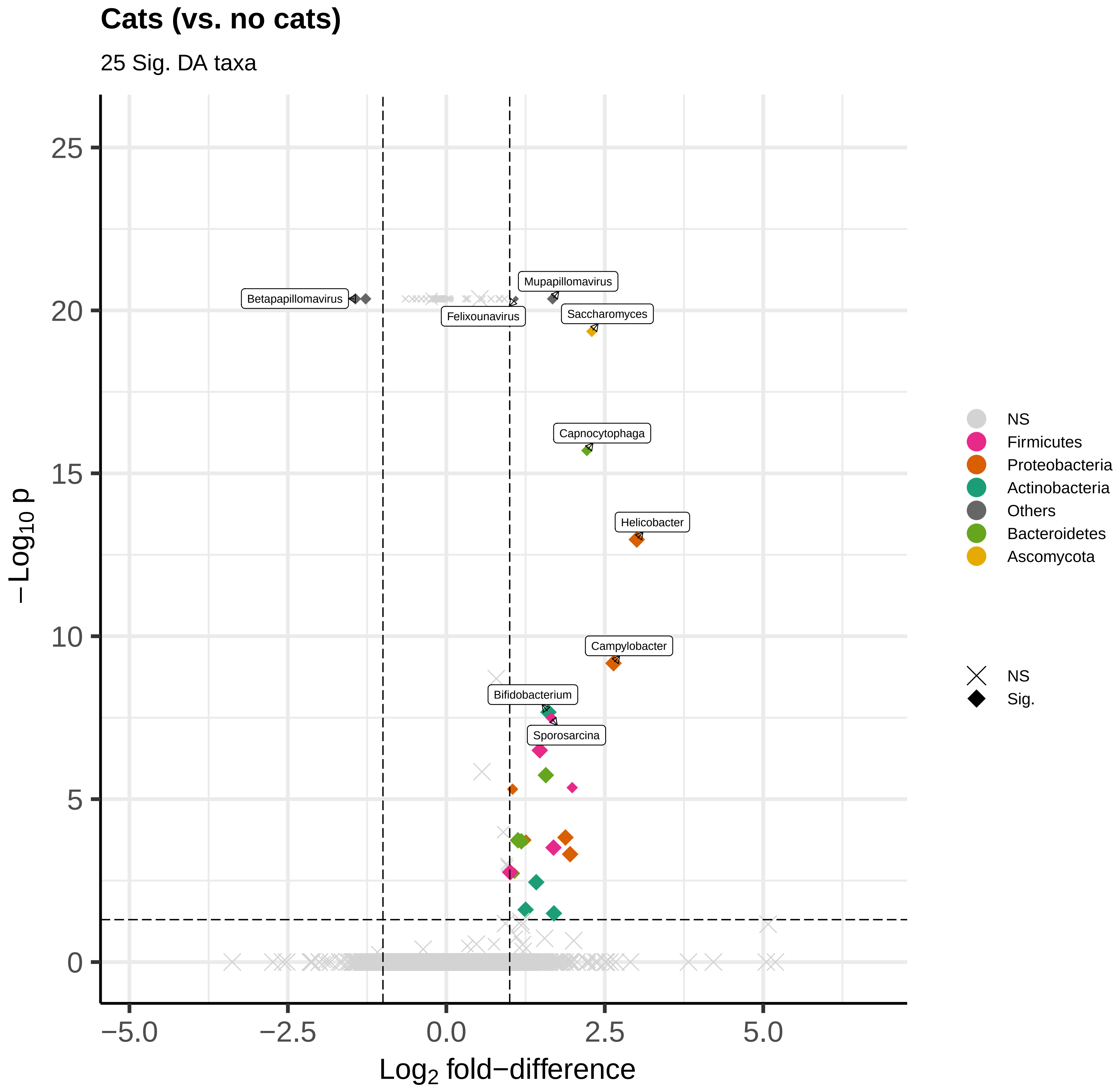
Cats (vs. no cats)
25 Sig. DA taxa
Betapapillomavirus
Mupapillomavirus
Felixounavirus
Saccharomyces
Capnocytophaga
Helicobacter
Campylobacter
Bifidobacterium
Sporosarcina
25
20
15
10
5
0
−5.0
−2.5
0.0
2.5
5.0
$-\mathrm{Log}_{10}$ p
$\mathrm{Log}_2$ fold−difference
NS
Firmicutes
Proteobacteria
Actinobacteria
Others
Bacteroidetes
Ascomycota
NS
Sig.

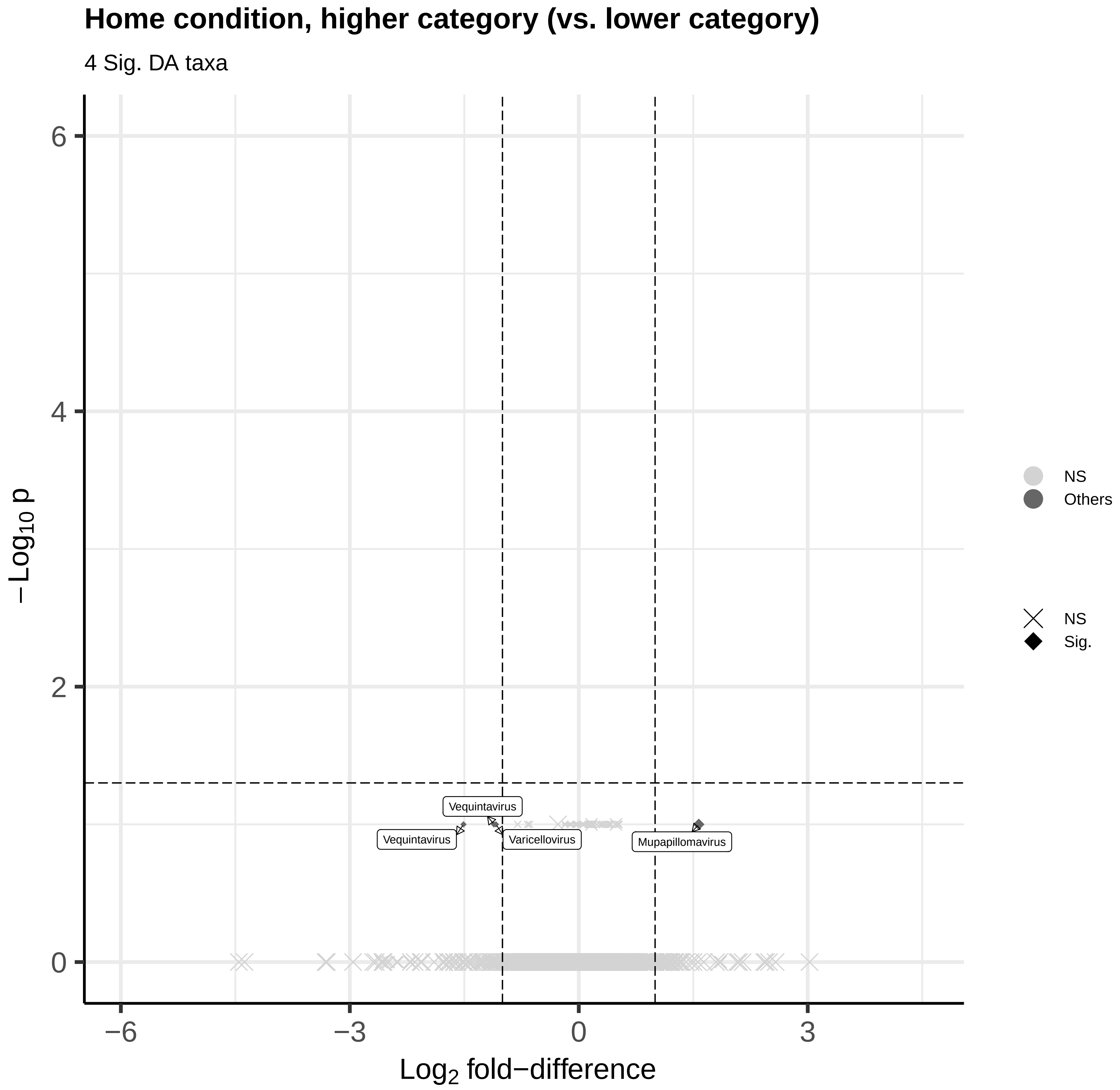
Home condition, higher category (vs. lower category)
4 Sig. DA taxa
−Log10 p
6
4
2
0
−6
−3
0
3
Log2 fold−difference
Vequintavirus
Vequintavirus
Varicellovirus
Mupapillomavirus
NS
Others
NS
Sig.

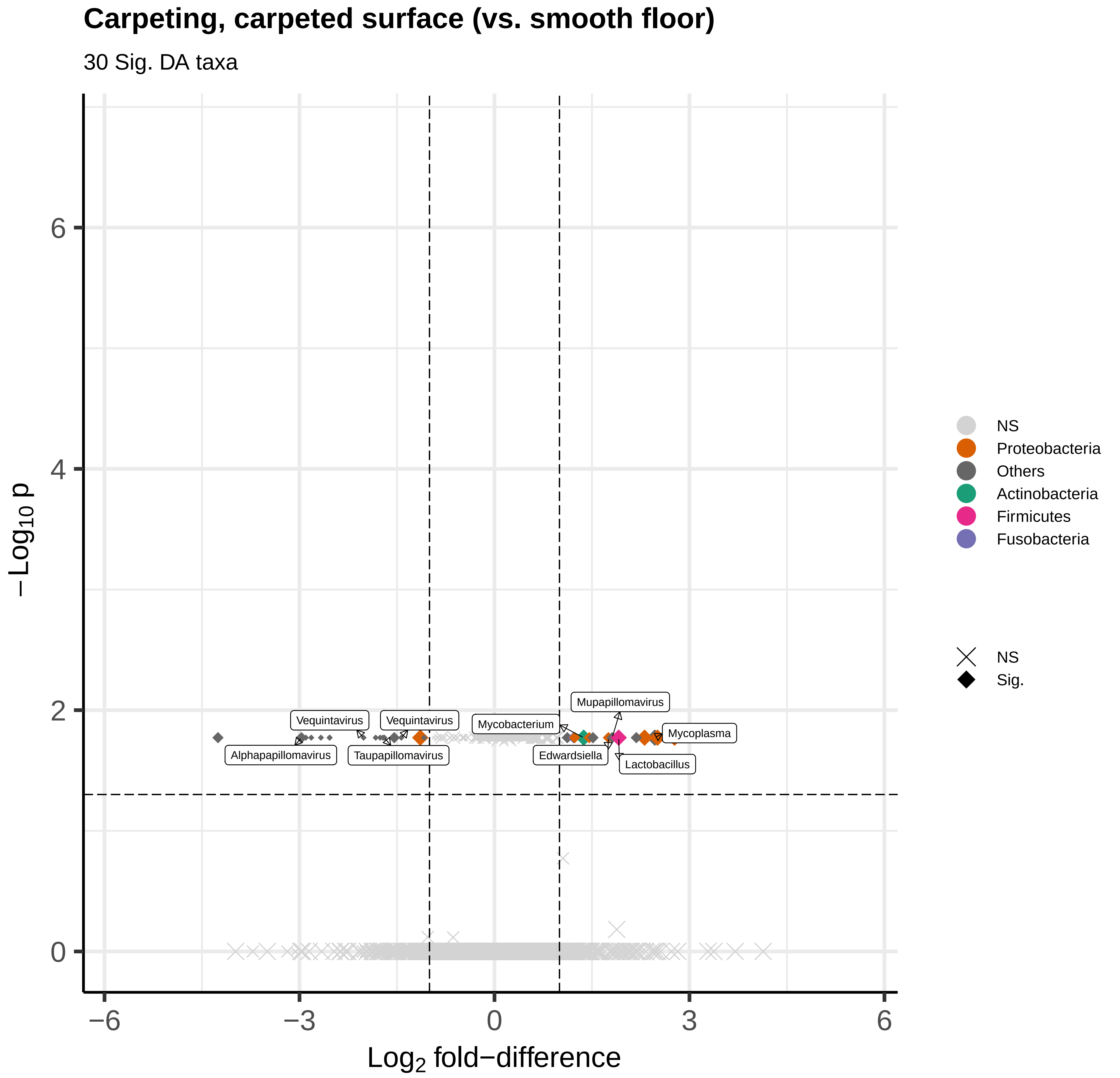

Carpeting, carpeted surface (vs. smooth floor)
30 Sig. DA taxa
−Log10 p
Log2 fold−difference
0
2
4
6
−6
−3
0
3
6
Vequintavirus
Vequintavirus
Alphapapillomavirus
Taupapillomavirus
Mycobacterium
Mupapillomavirus
Mycoplasma
Edwardsiella
Lactobacillus
NS
Proteobacteria
Others
Actinobacteria
Firmicutes
Fusobacteria
NS
Sig.

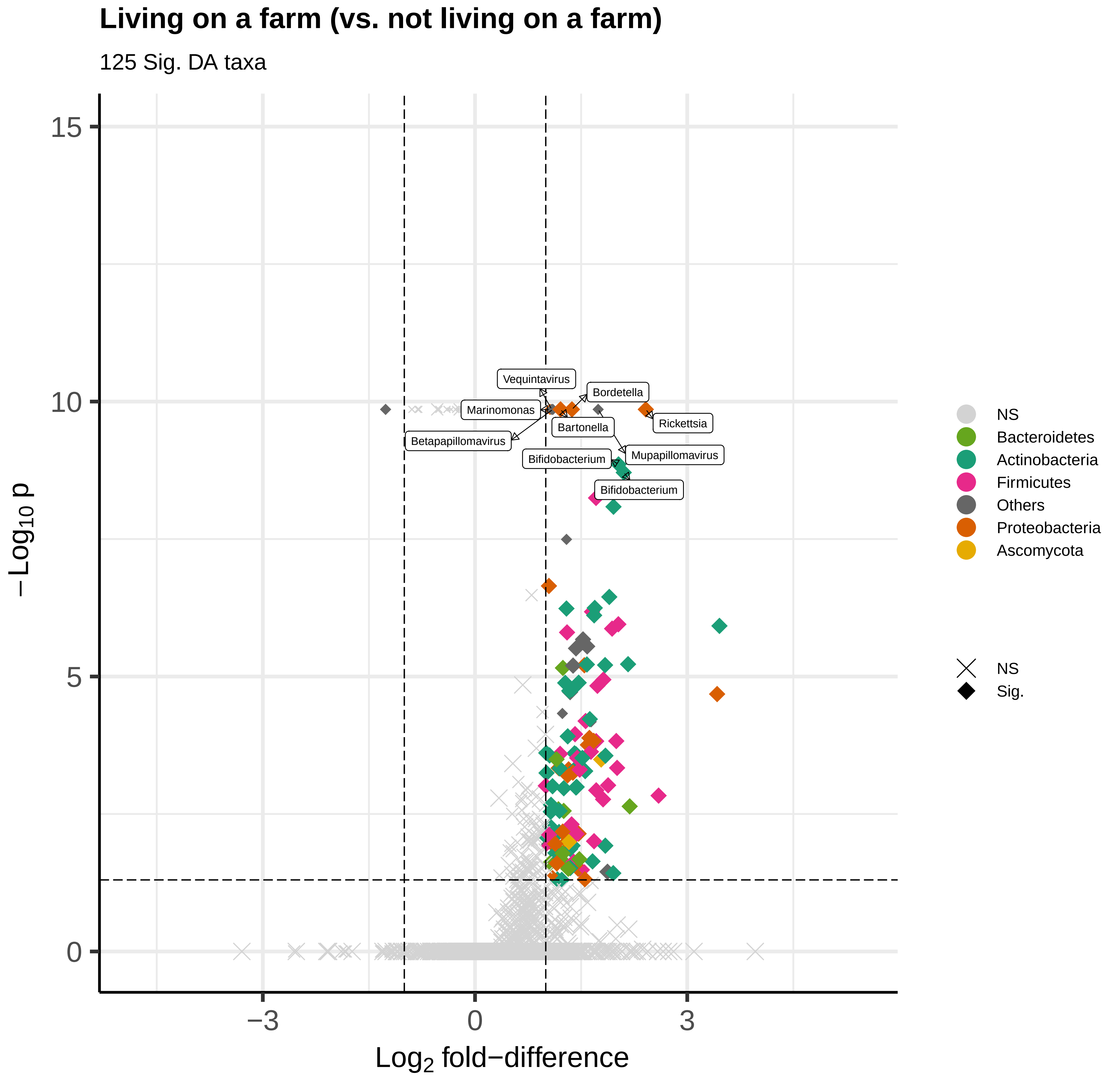

Living on a farm (vs. not living on a farm)
125 Sig. DA taxa
15
10
5
0
−Log10 p
−3
0
3
Log2 fold−difference
Vequintavirus
Bordetella
Marinomonas
Rickettsia
Bartonella
Betapapillomavirus
Mupapillomavirus
Bifidobacterium
Bifidobacterium
NS
Bacteroidetes
Actinobacteria
Firmicutes
Others
Proteobacteria
Ascomycota
NS
Sig.

### Crop farming (vs. no crop farming)

24 Sig. DA taxa

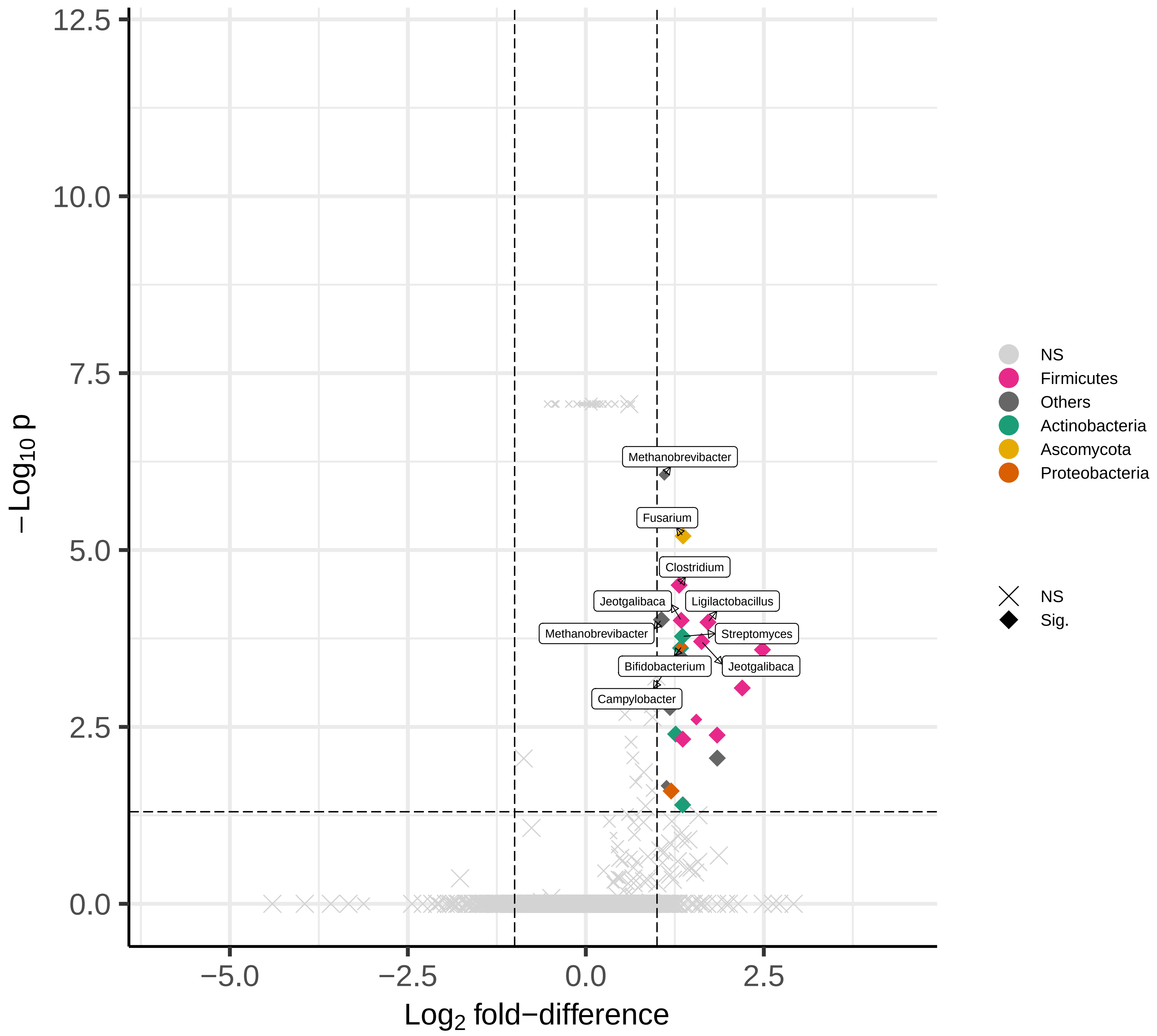

**Animal farming (vs. no animal farming)**

238 Sig. DA taxa

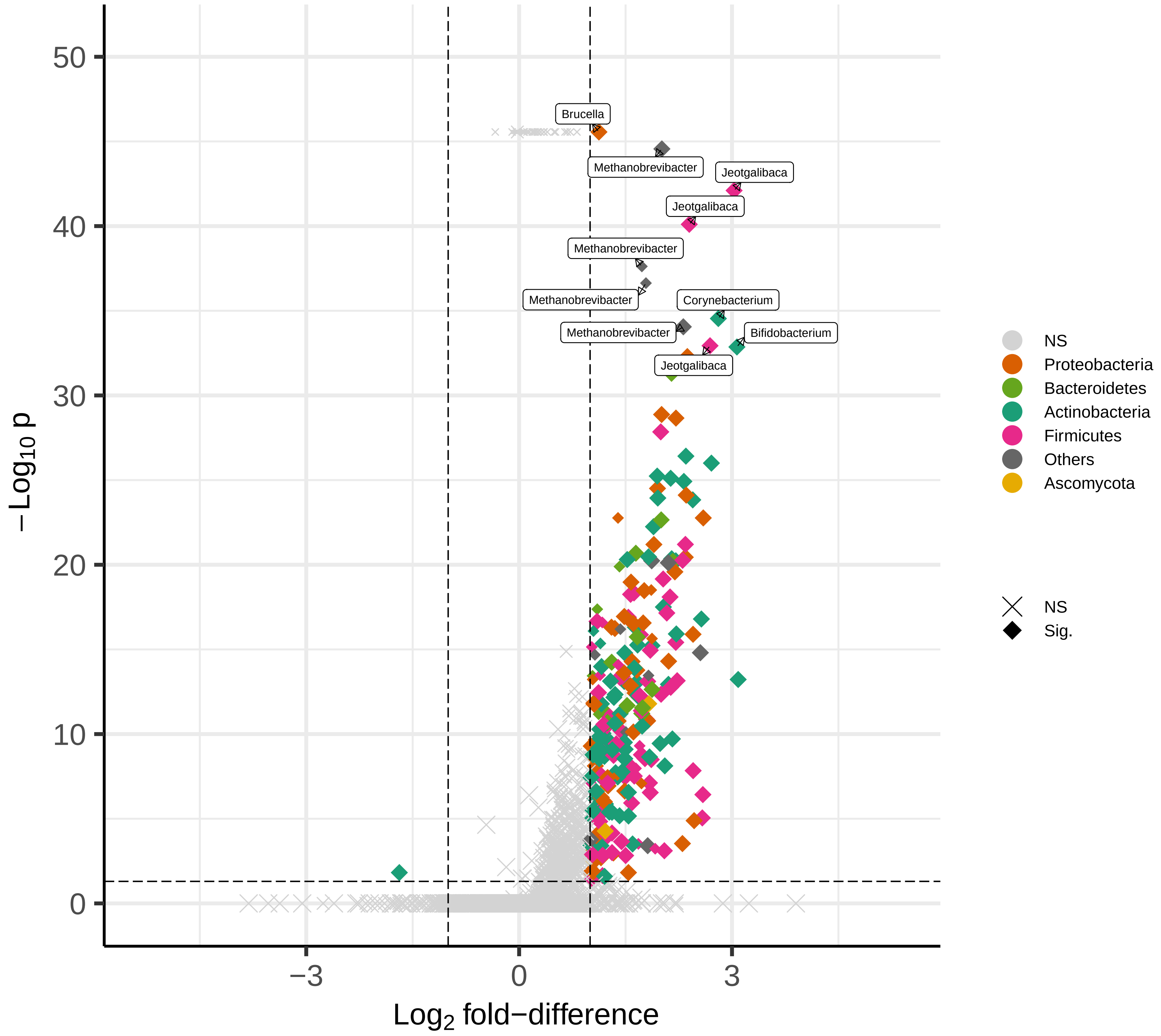

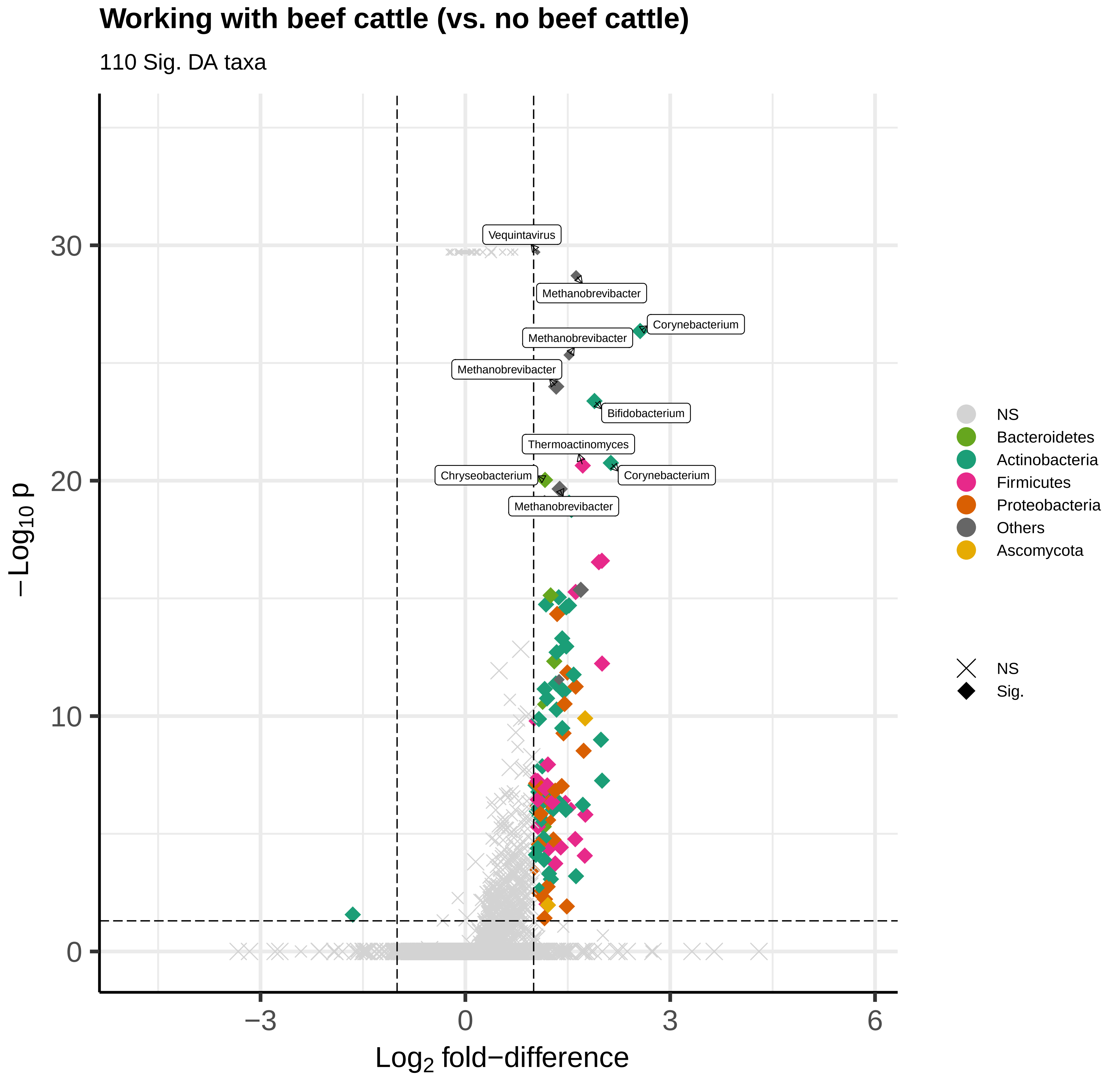
Working with beef cattle (vs. no beef cattle)
110 Sig. DA taxa
Vequintavirus
Methanobrevibacter
Corynebacterium
Methanobrevibacter
Methanobrevibacter
Bifidobacterium
Thermoactinomyces
Chryseobacterium
Corynebacterium
Methanobrevibacter
30
20
10
0
−3
0
3
6
$-\text{Log}_{10}\ p$
$\text{Log}_2$ fold−difference
NS
Bacteroidetes
Actinobacteria
Firmicutes
Proteobacteria
Others
Ascomycota
NS
Sig.

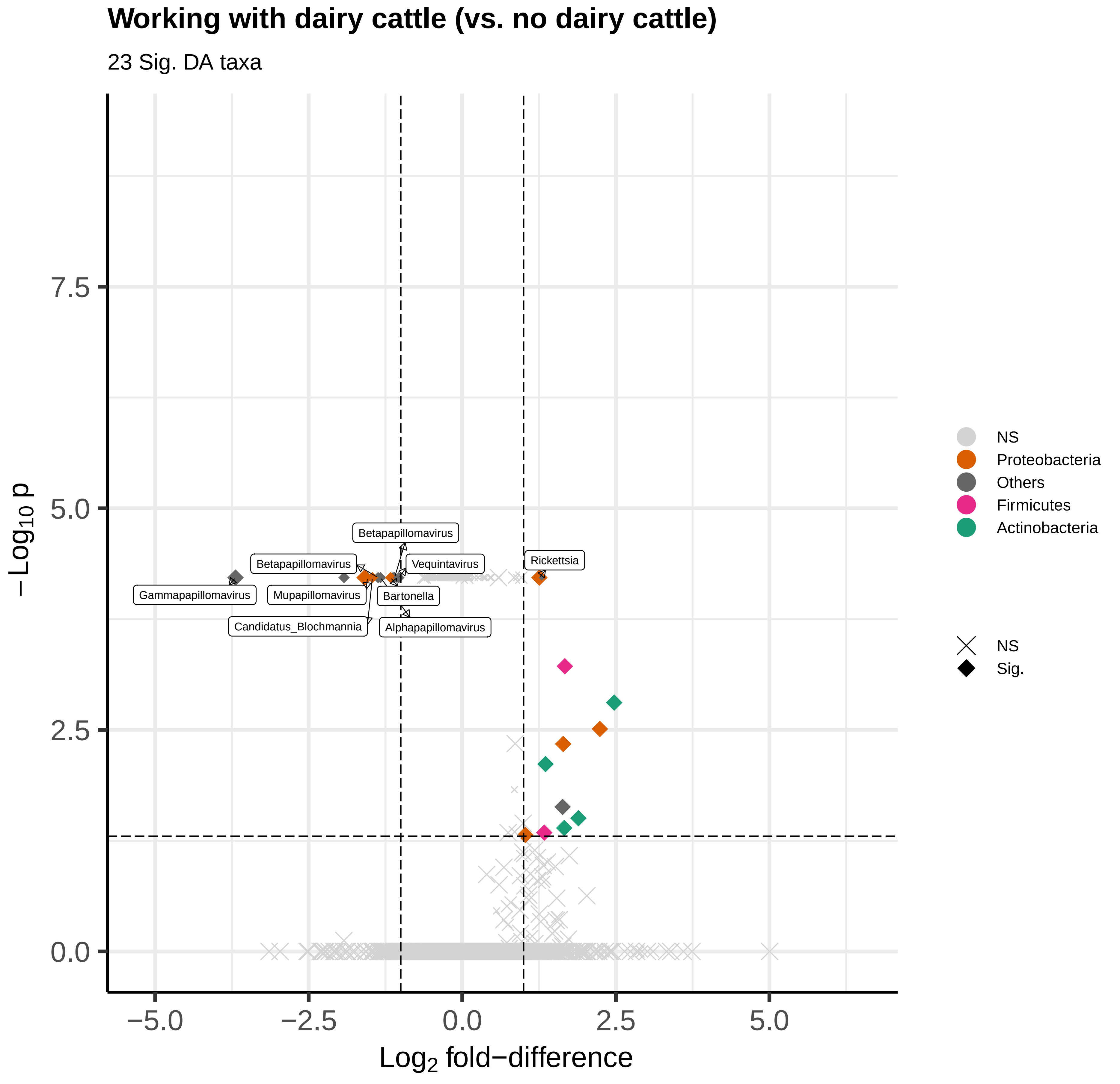
Working with dairy cattle (vs. no dairy cattle)
23 Sig. DA taxa
−Log10 p
Log2 fold−difference
7.5
5.0
2.5
0.0
−5.0
−2.5
0.0
2.5
5.0
Betapapillomavirus
Betapapillomavirus
Vequintavirus
Rickettsia
Gammapapillomavirus
Mupapillomavirus
Bartonella
Candidatus_Blochmannia
Alphapapillomavirus
NS
Proteobacteria
Others
Firmicutes
Actinobacteria
NS
Sig.

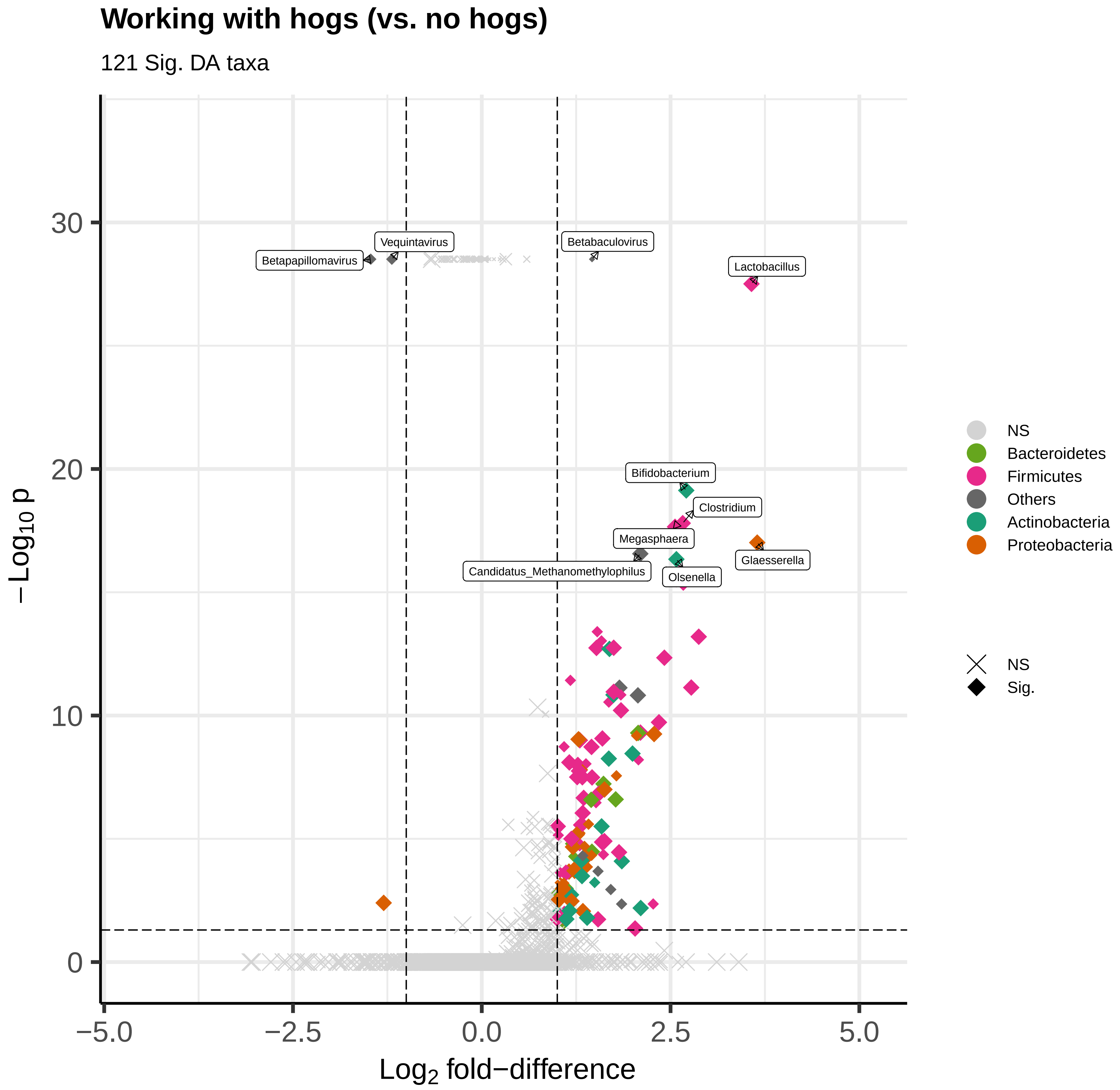
Working with hogs (vs. no hogs)
121 Sig. DA taxa
Betapapillomavirus
Vequintavirus
Betabaculovirus
Lactobacillus
Bifidobacterium
Clostridium
Megasphaera
Candidatus_Methanomethylophilus
Olsenella
Glaesserella
30
20
10
0
−5.0
−2.5
0.0
2.5
5.0
$-Log_{10}$ p
$Log_2$ fold−difference
NS
Bacteroidetes
Firmicutes
Others
Actinobacteria
Proteobacteria
NS
Sig.

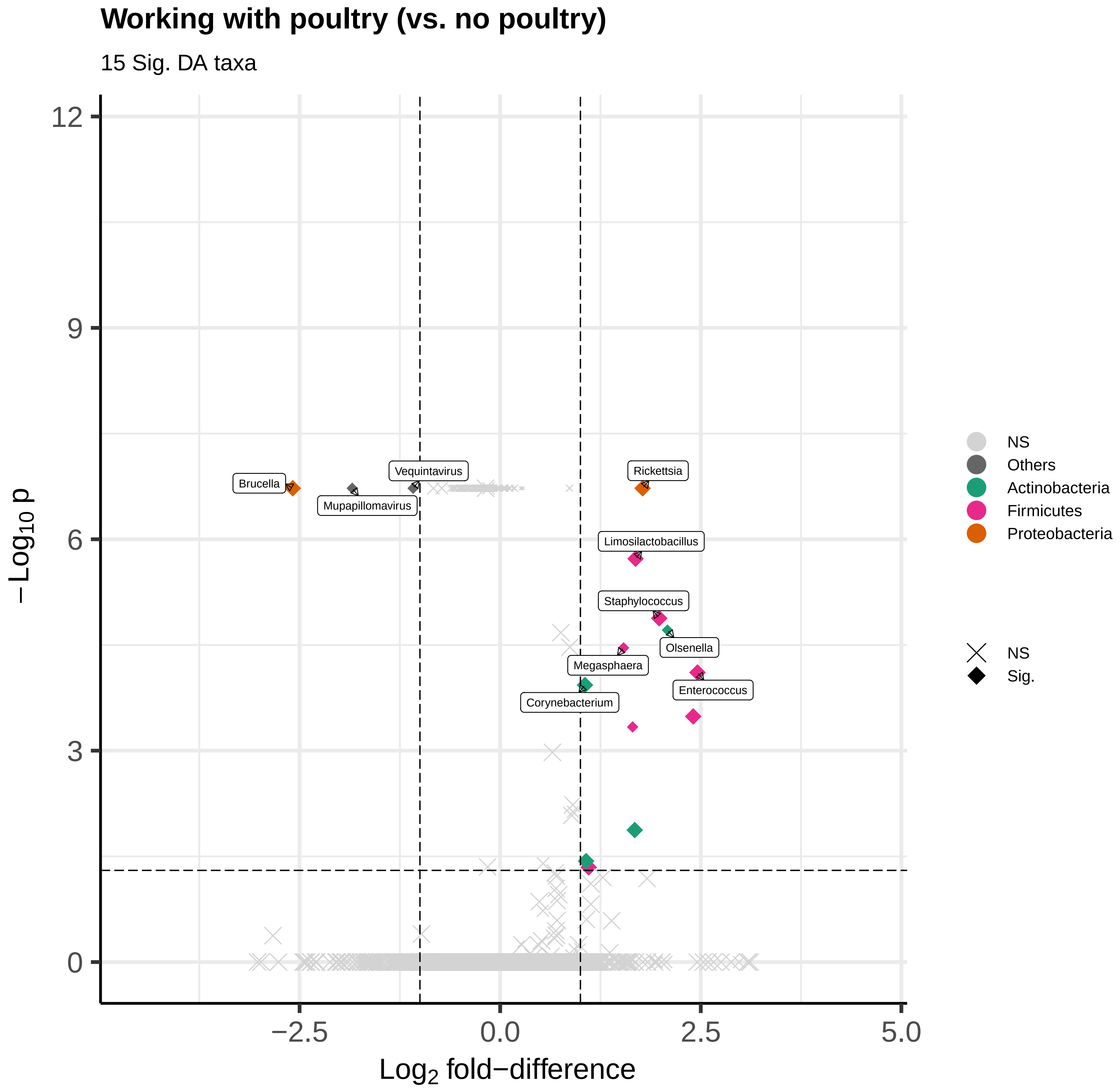
Working with poultry (vs. no poultry)
15 Sig. DA taxa
−Log10 p
Log2 fold−difference
12
9
6
3
0
−2.5
0.0
2.5
5.0
Brucella
Vequintavirus
Mupapillomavirus
Rickettsia
Limosilactobacillus
Staphylococcus
Olsenella
Megasphaera
Enterococcus
Corynebacterium
NS
Others
Actinobacteria
Firmicutes
Proteobacteria
NS
Sig.

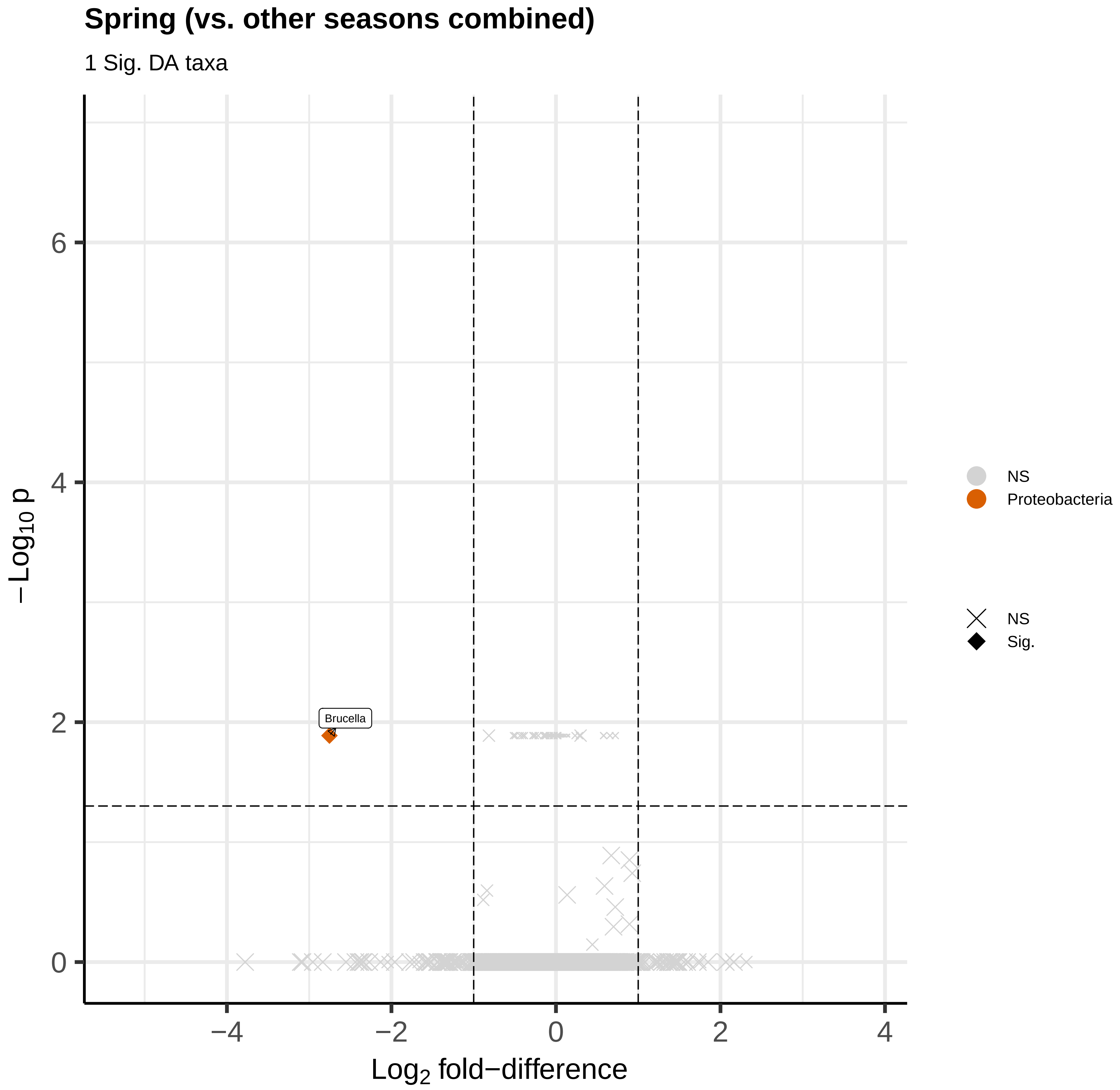

Spring (vs. other seasons combined)
1 Sig. DA taxa
Brucella
−Log10 p
Log2 fold−difference
0
2
4
6
−4
−2
0
2
4
NS
Proteobacteria
NS
Sig.

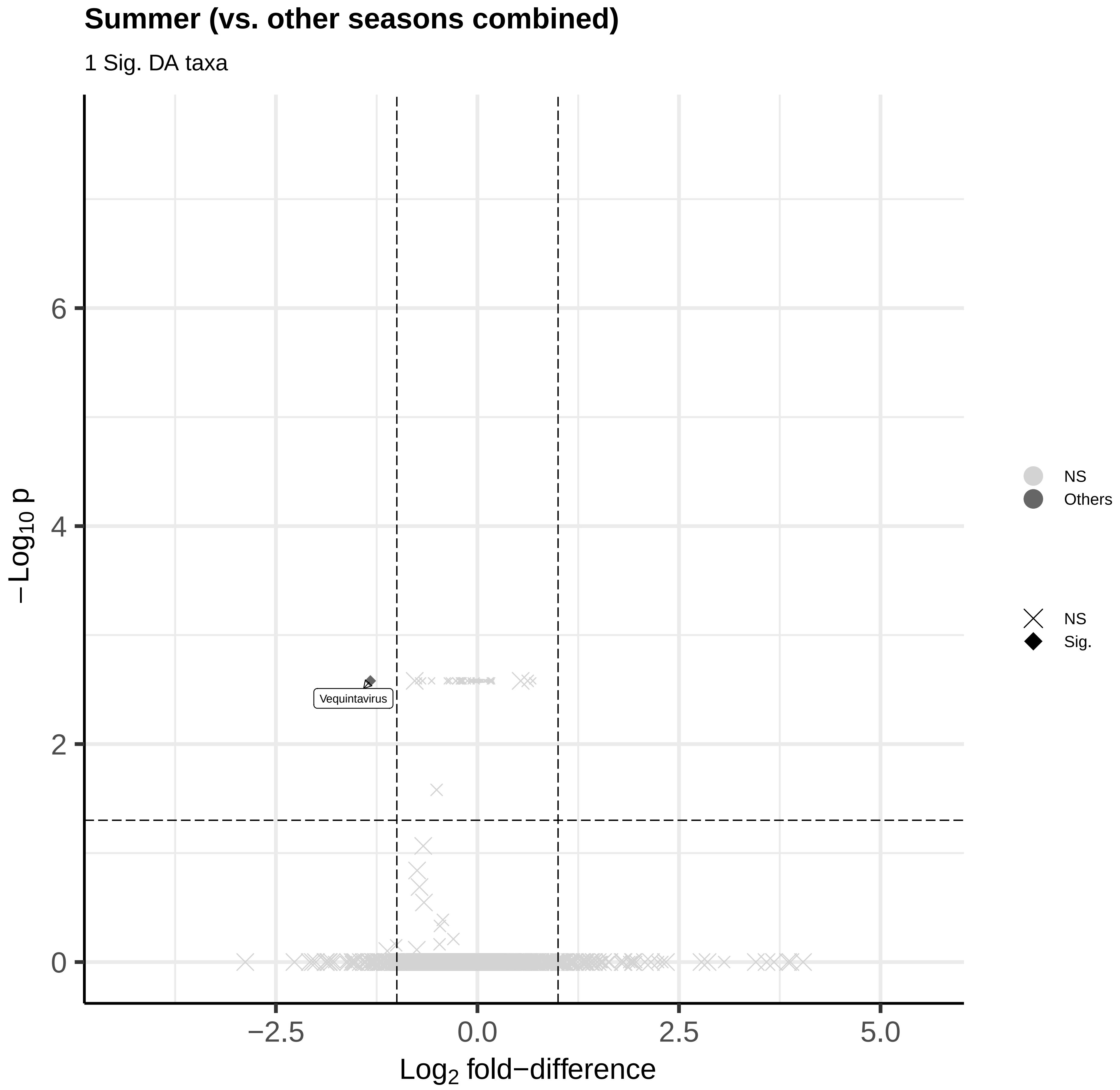

Summer (vs. other seasons combined)
1 Sig. DA taxa
−Log10 p
6
4
2
0
Vequintavirus
−2.5
0.0
2.5
5.0
Log2 fold−difference
NS
Others
NS
Sig.

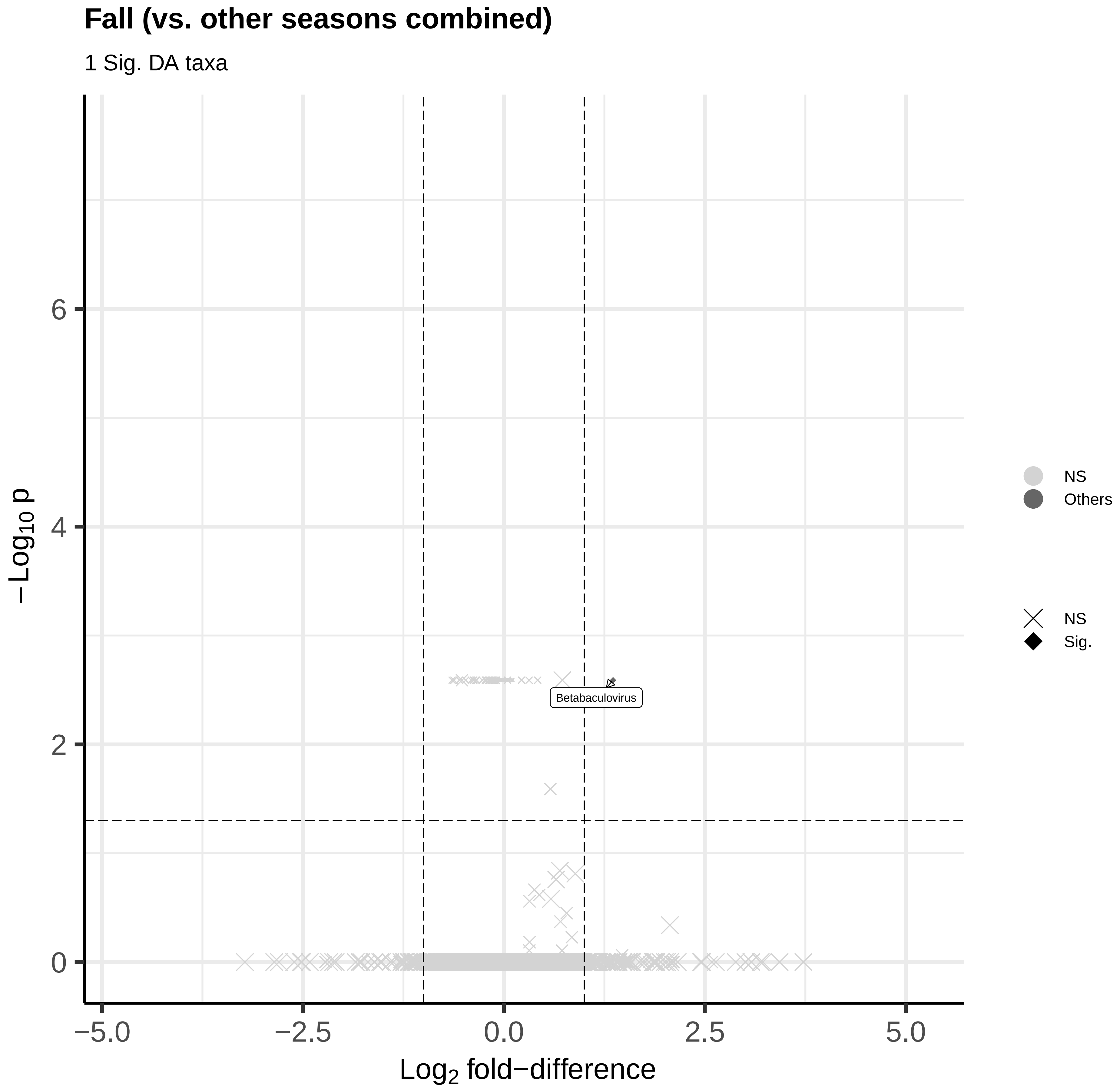

Fall (vs. other seasons combined)
1 Sig. DA taxa
−Log10 p
Log2 fold−difference
6
4
2
0
−5.0
−2.5
0.0
2.5
5.0
Betabaculovirus
NS
Others
NS
Sig.

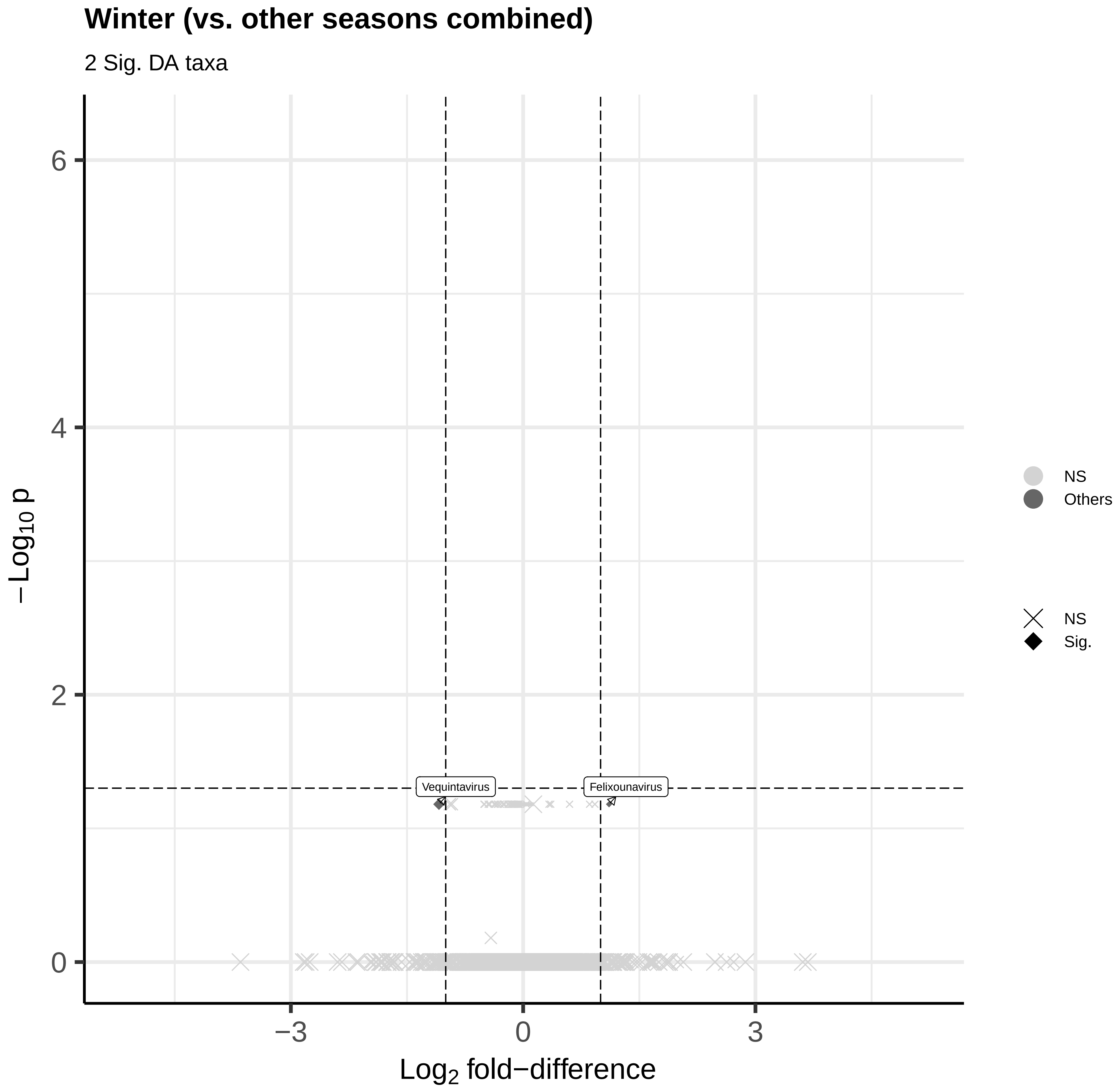

Winter (vs. other seasons combined)
2 Sig. DA taxa
Vequintavirus
Felixounavirus
6
4
2
0
−3
0
3
Log2 fold−difference
−Log10 p
NS
Others
NS
Sig.
